# Comparative Effectiveness of mRNA-1273 and BNT162b2 COVID-19 Vaccines Among Older Adults: Systematic Literature Review and Meta-Analysis Using the GRADE Framework

**DOI:** 10.1101/2023.11.21.23298832

**Authors:** Sushma Kavikondala, Katrin Haeussler, Xuan Wang, Mary T. Bausch-Jurken, Maria Nassim, Nitendra Kumar Mishra, Mia Malmenäs, Pawana Sharma, Nicolas Van de Velde, Nathan Green, Ekkehard Beck

## Abstract

**Background:** The mRNA vaccines mRNA-1273 and BNT162b2 demonstrated high efficacy against SARS-CoV-2 infection in phase 3 clinical trials, including among older adults. To inform COVID-19 vaccine selection, this systematic literature review (SLR) and meta-analysis assessed the comparative effectiveness of mRNA-1273 versus BNT162b2 in older adults.

**Methods:** We systematically searched for relevant studies reporting COVID-19 outcomes with mRNA vaccines in older adults aged ≥50 years by first cross-checking relevant published SLRs. Based on the cutoff date from a previous similar SLR, we then searched the WHO COVID-19 Research Database for relevant articles published between April 9, 2022 and June 2, 2023. Outcomes of interest were SARS-CoV-2 infection, symptomatic SARS-CoV-2 infection, severe SARS-CoV-2 infection, COVID-19‒related hospitalization, and COVID-19‒related death following ≥2 vaccine doses. Random-effects meta-analysis models were used to pool risk ratios (RRs) across studies. Heterogeneity was evaluated using chi-squared testing. Evidence certainty was assessed per GRADE framework.

**Results:** 24 non-randomized real-world studies reporting clinical outcomes with mRNA vaccines in individuals aged ≥50 years were included in the meta-analysis. Vaccination with mRNA-1273 was associated with significantly lower risk of SARS-CoV-2 infection (RR 0.72 [95% confidence interval (CI) 0.64‒0.80]), symptomatic SARS-CoV-2 infection (RR 0.72 [95% CI 0.62‒0.83]), severe SARS-CoV-2 infection (RR 0.67 [95% CI 0.57‒0.78]), COVID-19‒related hospitalization (RR 0.65 [95% CI 0.53‒0.79]) and COVID-19‒related death (RR 0.80 [95% CI 0.64‒0.99]) compared with BNT162b2. There was considerable heterogeneity between studies for all outcomes (I^2^>75%) except death (I^2^=0%). Multiple subgroup and sensitivity analyses excluding specific studies generally demonstrated consistent results. Certainty of evidence across outcomes was rated as low (type 3) or very low (type 4), reflecting the lack of randomized-controlled trial data.

**Conclusion:** Meta-analysis of 24 observational studies demonstrated significantly lower risk of asymptomatic, symptomatic, and severe infections; hospitalizations; and deaths with the mRNA-1273 versus BNT162b2 vaccine in older adults aged ≥50 years.

**SUMMARY POINTS:**

- The COVID-19 pandemic has disproportionately affected older adults, as this population is generally more susceptible to infection and severe outcomes due to immune senescence and underlying comorbidities.
- The 2 available mRNA vaccines mRNA-1273 and BNT162b2 demonstrated high efficacy against SARS-CoV-2 infection in phase 3 clinical trials, including among older adults.
- To inform COVID-19 vaccine selection, this systematic literature review and meta-analysis assessed the comparative effectiveness of mRNA-1273 versus BNT162b2 among older adults in real-world settings.
- Vaccination with homologous primary or booster mRNA-1273 was associated with significantly lower risk of infection (including asymptomatic, symptomatic, and severe infections), hospitalization, and death due to COVID-19 than vaccination with BNT162b2 in older adults aged ≥50 years.

## INTRODUCTION

As of October 2023, the global coronavirus disease 2019 (COVID-19) pandemic has resulted in more than 771.4 million reported infections and over 6.9 million deaths due to severe acute respiratory syndrome coronavirus 2 (SARS-CoV-2) (1). COVID-19 has disproportionately affected older adults (2, 3, 4, 5). Worldwide, older adults aged ≥60 years accounted for 80% of COVID-19-associated deaths reported to the World Health Organization (WHO) via detailed weekly surveillance from January 2020 to December 2021, and were estimated to account for 82% of deaths based on the WHO excess mortality model (4). Immune senescence and underlying comorbidities make older adults generally more susceptible to COVID-19 and associated severe outcomes. Several studies have identified older age as a primary risk factor for severe illness with COVID-19 (6, 7, 8), with one study demonstrating similar performance between a risk score that was based on age alone versus a validated risk score incorporating the effects of multiple underlying comorbidities (POINTED score) (9). Importantly, the WHO has identified older adults (commonly defined by age cutoffs of 50 to 60 years, depending on the country) as a high-priority group for COVID-19 vaccination (10), and many countries have prioritized vaccination of the older population (9).

A previous meta-analysis of 32 studies in older adults aged ≥55 years found that vaccination with either one of the 2 vaccines employing novel messenger ribonucleic acid (mRNA) technology provided the highest protection against COVID-19 compared with other vaccine types (11). The mRNA vaccines were developed and granted emergency use authorization in late 2020 to globally mitigate the spread of SARS-CoV-2: mRNA-1273 (Spikevax^®^; Moderna, Inc., Cambridge, MA, USA) (12) and BNT162b2 (Comirnaty®; Pfizer/BioNTech, New York, NY, USA/Mainz, Germany) (13). Phase 3 trials of these vaccines demonstrated high vaccine efficacy against SARS-CoV-2 infection when administered as 2-dose regimens (94.1% and 95.0% effectiveness with mRNA-1273 and BNT162b2, respectively) (14, 15), with subgroup analyses also confirming high vaccine efficacy in older participants (aged ≥65 years) (14, 15).

Although both mRNA-1273 and BNT162b2 are based on mRNA technology, their formulations differ. For example, the mRNA-1273 vaccine contains more active ingredient (100 µg of mRNA for primary; 50 µg for booster) than the BNT162b2 vaccine (30 µg of mRNA for both primary and booster) (12, 13, 16, 17) and uses a different lipid nanoparticle delivery system (18, 19, 20). As shown with other respiratory vaccines (21, 22), and as demonstrated in immunocompromised individuals (23), these differences may impact vaccine effectiveness in older adults. Thus, to inform COVID-19 vaccine selection and policy decision-making in the absence of head-to-head comparisons of the mRNA-1273 and BNT162b2 vaccines in randomized controlled trials (RCTs), there remains a need to synthesize evidence across real-world studies to provide robust information about the comparative effectiveness of the two mRNA vaccines among older adults.

We performed a systematic literature review and pairwise meta-analysis to compare the effectiveness of mRNA-1273 versus BNT162b2 in older adults. Our analysis followed the Grading of Recommendations, Assessment, Development and Evaluations (GRADE) framework (24) used by national immunization advisory groups when developing recommendations (25). The question addressed in the present research was ‘Is mRNA-1273 more effective than BNT162b2 at preventing SARS-CoV-2 infections and COVID-19-related hospitalizations and deaths in older adults aged ≥50 years?’.

## METHODS

### Search strategy and study selection

This systematic literature review and meta-analysis is registered in Prospero (CRD42023443149) and was conducted in accordance with the Preferred Reporting Items for Systematic Reviews and Meta-Analyses 2020 framework (26). Studies were identified using a 2-step search procedure. First, the WHO COVID-19 Research Database was searched to identify systematic literature reviews on COVID-19 vaccination in older adults aged ≥50 years published between March 2020 and 19 April 2023. Sixteen of 67 systematic reviews identified were relevant (Supplemental Table S1) and were cross-checked for articles to be included in abstract screening and full-text assessment. One prior systematic review identified had similar objectives to the current study (11) and all studies included in this prior review were included for full-text assessment. A total of 243 studies for full-text screening were identified from this first step. The main search was then conducted in the WHO COVID-19 Research Database to identify relevant studies published since the prior similar systematic review (11), which included studies from database inception through 9 April 2022 to 2 June 2023. Notably, although the WHO COVID-19 Research Database remains searchable, updates ceased in June 2023 (27), thus content spans March 2020 through June 2023. Databases searched include MEDLINE/PubMed, International Clinical Trials Registry Platform, Embase, EuropePMC, medRxiv, Web of Science, ProQuest Central, Academic Search Complete, Scopus, and COVIDWHO. The main database search identified an additional 1012 studies for full-text screening. The main search strategy is summarized in Supplemental Table S2.

RCTs, observational studies, or any real-world evidence published as full-text manuscripts, letters, commentaries, abstracts, or posters were included if they reported prespecified COVID-19 outcomes (described below) in older adults aged ≥50 years who received mRNA-1273 or BNT162b2 within the same study (studies with ≤10% of the study population aged ≤50 years included). Studies could include participants who had comorbidities, and those who were immunocompetent or defined as clinically extremely vulnerable (CEV) with conditions in CEV group 3, as categorized by Canadian Health Services (28) (studies with ≤10% of participants with CEV group 1 and 2 conditions were included). Diabetes was considered a CEV group 3 condition regardless of whether the patient was being treated with insulin. Only studies reporting the outcomes of interest for participants who received ≥2 vaccine doses were included and wherever available three-dose data was considered primarily. If a study did not report the outcomes for participants who received three doses, then two-dose or four-dose data was considered. Only homologous dose series (≥2 doses of mRNA-1273 or ≥2 doses BNT162b2) were included in analysis.

Outcomes of interest were vaccine efficacy or effectiveness against SARS-CoV-2 infection (defined as asymptomatic or symptomatic infection with positive test or a COVID-19 diagnosis code [U07.1]), laboratory-confirmed symptomatic SARS-CoV-2 infection (defined as positive test with symptoms including but not limited to fever, cough, shortness of breath and sudden onset of anosmia/ageusia; in some countries, runny nose was also included in the case definition), severe SARS-CoV-2 infection (defined specifically as severe infection or as hospitalization or death, as reported in the study; primarily defined by severe infection, followed by hospitalization and lastly by death if data on multiple endpoints were available), COVID-19‒related hospitalization (defined as intubation, hospitalization, or admission to intensive care unit with positive test for SARS-CoV-2 infection within 5 days before to 28 days from admission; cases with information on intubation but not hospitalization were assumed to be hospitalized), or COVID-19‒related death (defined as deaths occurring after a positive test for SARS-CoV-2 infection without previously declared recovery or another clear cause of death reported). A positive SARS-CoV-2 test could be based on any of the following methods, as reported by individual studies: reverse transcription polymerase chain reaction (PCR), rapid antigen test, or dried blood spot seropositivity for anti-nucleocapsid immunoglobulin G antibodies by validated enzyme-linked immunosorbent assay. Infections were considered if they occurred ≥7 days after the last vaccination. Only those studies that reported the following data were included in the meta-analysis: number of events and sample size per arm; or vaccine effectiveness (VE) per arm and subgroup derived as 1−risk ratio (RR), 1−odds ratio (OR), 1−hazard ratio (HR), or 1−incidence rate ratio (IRR). For the analyses of VE, if only VE data and total numbers of participants by vaccine arm were available, then the weighted average VE for all age groups among individuals aged ≥50 years was computed. Weighted average was calculated as the sum of the VE in all age groups in a vaccine arm divided by the total number of participants in that arm. If only VE data were available without participant numbers by vaccine arm, then VE in the age group that most closely matched the data within the studies in the meta-analysis was selected.

Studies in pregnant women, current or former smokers, and physically inactive participants; studies including only immunocompromised individuals with conditions within CEV groups 1 and 2; and studies with only safety and/or immunogenicity outcomes were excluded. The population, exposure, comparison, and outcomes used in the systematic literature review are summarized in **Supplemental Table S3**. Two independent reviewers selected studies using a 2-level approach; discrepancies were resolved by consensus or by a third reviewer. In level 1, titles and abstracts were screened against inclusion criteria; then in level 2, articles not excluded at level 1 underwent full-text screening against the selection criteria.

### Data extraction and quality assessment

Study design details, baseline characteristics of study participants, vaccine received and dosing details, and vaccine efficacy/effectiveness outcomes were extracted from the selected studies. Risk of bias was assessed using the Newcastle-Ottawa Scale (29) for observational studies. The certainty of evidence was evaluated based on GRADE criteria (24, 25).

### Statistical analysis

Random-effects meta-analysis models were used to pool RRs and to estimate absolute effects as risk difference (RD) per 100,000 individuals across the included studies, comparing mRNA-1273 to BNT162b2. The inverse variance method was applied for the random effects models (30). Details regarding methodology of the analyses are included in **Appendix 1**. Briefly, a standard pairwise meta-analysis was conducted using RRs instead of number of events and sample size per arm as the data input. However, due to differences in how outcomes were reported across studies, a conversion approach (31, 32, 33) was implemented. For studies that reported the number of events and sample size per arm, unadjusted RRs were estimated straightforwardly. For studies that exclusively reported VE, instead of number of events and sample size per arm, RR was either estimated as “1–VE” (for studies reporting VE as 1–RR) or estimated from VE through optimal approximate conversions of contrast-based data (**Supplemental Figure S1**). As a sensitivity analysis, a second-order meta-analysis approach was implemented to avoid the assumptions based on converting contrast-based data in the conversion approach. With this approach, data from studies reporting number of events and sample size were pooled in one meta-analysis, and data from studies reporting only VE were pooled in a second meta-analysis (i.e., without distinction as to how VE was estimated and without any conversion). In the second-order meta-analysis, the pooled results from these separate meta-analyses on RRs informed the analysis, resulting in the final RR estimate. Absolute effects (RD) cannot be reliably estimated using this second-order approach, so this method was used only for analysis of RR.

As additional sensitivity analyses, outcomes were assessed in the following subgroups: individuals aged ≥65 years; individuals aged ≥75 years; individuals who received exclusively three doses of the same vaccine; individuals aged ≥50 years, excluding those with disease conditions categorized in CEV groups 1 or 2; individuals infected with the SARS-CoV-2 Delta variant (ie, dominant variant during study time period); and excluding those studies that reported only VE.

Publication bias was assessed by visual examination of funnel plots and Egger’s regression test for asymmetry (34, 35). Heterogeneity across studies was evaluated using chi-squared testing (36), with the percentage of variation across studies estimated using the I^2^ statistic (scale of 0%–100%, with 0% meaning no evidence of heterogeneity; see **Appendix 1**). Results were summarized in forest plots to display the effect estimates with 95% CIs for the individual studies and the pooled estimate of the meta-analysis. The meta-analyses were conducted in R (v4.3.1), using the meta (37) and metafor (38) packages.

## RESULTS

### Search results and included studies

In total, 1255 abstracts identified from either the 16 relevant SLRs that were crosschecked (n=243) or the main search in the WHO COVID-19 database (n=1012) were screened for inclusion (**Figure 1**). Of these, 25 studies (all non-randomized) reported results for the clinical outcomes of interest in individuals aged ≥50 years, 24 of which were included in the meta-analysis (one study (39) was excluded because it reported only RR thus did not meet the prespecified criterion of reporting number of events and sample size or VE).

**Figure 1.**
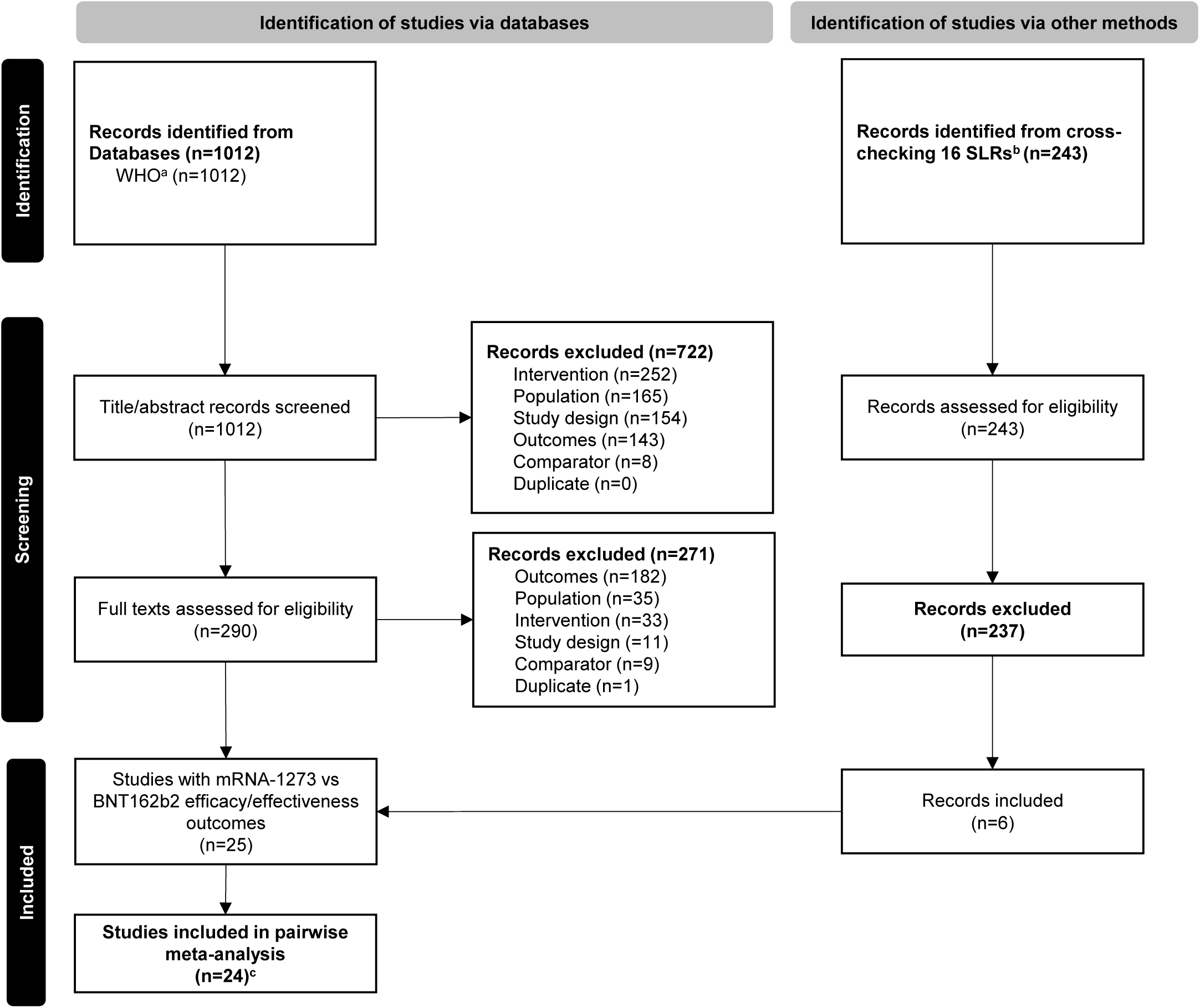
PRISMA flow diagram. ^a^Databases searched include ICTRP, EMBASE, EuropePMC, medRxiv, Web of Science, ProQuest Central, Academic Search Complete, Scopus, and COVIDWHO. ^b^16 recently published SLRs and internal documents from Moderna, Inc were cross-checked. ^c^One study (39) was excluded from the network meta-analysis because the presented data were not comparable to the data from other studies. SLR, systematic literature review.

Characteristics of each of the studies included in the meta-analysis are summarized in **Table 1**. Of the 24 studies, 1 was industry-sponsored. Overall, the studies included >3.9 million older adults (aged ≥50 years) vaccinated with mRNA-1273 and >5.2 million vaccinated with BNT162b2. Most studies involved North American (Canada, n=2 (40, 41); United States, n=11 (42, 43, 44, 45, 46, 47, 48, 49, 50, 51, 52)) or European (Belgium, n=1 (53); Greece, n=1 (54); Hungary, n=2 (55, 56); Norway, n=1 (57); Netherlands, n=1 (58); Spain, n=2 (59, 60); multiple countries, n=1 (61)) populations. Although most studies included general population samples, two were restricted to nursing home or retirement home residents (40, 43) and two were restricted to Veteran’s Affairs populations in the United States (42, 44). The majority of studies specified the Delta variant as the SARS-CoV-2 variant of concern (42, 43, 45, 46, 47, 48, 50, 51, 53, 54, 56, 57, 58, 60, 61); 5 studies specified the Alpha variant (46, 49, 53, 54, 55) and 4 specified the Omicron variant (40, 41, 50, 53). Some studies with longer follow-up periods collected data during multiple COVID-19 seasons, therefore reported data on multiple variants, either not further specified or in separate subgroups. We conducted a subgroup analysis in patients infected with the Delta variant due to the large number of available studies; subgroup analysis for other variants was deemed unfeasible due to sparse data. In the majority of studies, positivity for SARS-CoV-2 infection was determined using PCR or an antigen test; however, four studies did not specify the testing method (44, 45, 47, 51) and one study used a nasopharyngeal PCR test and/or circulating antinucleocapsid IgG antibodies (40).

**Table 1.** Characteristics of studies included in the meta-analysis.

| Author, year | Study characteristics |  |  |  |  |  |  |  |  | Outcomes reported |  |  |  |  |
| --- | --- | --- | --- | --- | --- | --- | --- | --- | --- | --- | --- | --- | --- | --- |
|  | Design | Country and data source | Age group(s) | CEV group 1/2 | CEV group 3 | SARS CoV-2 testing method | No. Vaccine doses | Study period | Vaccinated, n | Infection | Symptomatic infection | Severe infection<sup>a</sup> | Hospitalization | Death |
| Bello-Chavolla 2023 (68) | • Retrospective analysis | • Mexico<br>• SISVER database | • ≥60 y | ND | Y | RT-PCR and/or antigen test | 2 doses (MM vs PP) | Dec 2020–Sep 2021 | • BNT162b2: 47,694<br>• mRNA-1273: 1,155 | Y | N | Y | N | Y |
| Braeye 2023 (53) | • Retrospective cohort study | • Belgium<br>• Belgium data collected between Jan 2021 and Jan 2022 | • 65–85 y | ND | ND | RT-PCR test | 2 doses (MM vs PP) | Jan 2021–Jan 2022 | • BNT162b2: 13,613<br>• mRNA-1273: 1,155 | Y | N | N | N | N |
| Breznik 2023 (40) | • Retrospective cohort study | • Canada<br>• 17 nursing homes and 8 retirement homes in Ontario, Canada | • ≥50 y | ND | ND | Nasopharyngeal PCR and/or circulating antinucleo-capsid IgG antibodies | 3 doses (MMM vs PPP) | Dec 2021–May 2022 | • BNT162b2: 478<br>• mRNA-1273: 420 | Y | N | N | N | N |
| Butt 2022 (42) | • Retrospective cohort study | • USA<br>• VA Healthcare System COVID-19 Shared Data Resource | • ≥50 y | Y | Y | PCR test | 3 doses (MMM vs PPP) | Apr 2021–Sep 2021 | • BNT162b2: 236,693<br>• mRNA-1273: 158,993 | Y | Y | Y | Y | Y |
| Chemaitelly 2022 (69) | • Retrospective cohort study | • Qatar<br>• The national, federated databases of the Qatar | • ≥50 y | ND | Y | PCR and/or antigen test | 2 doses (MM vs PP) | Feb 2020–May 2022 | • BNT162b2: 180,790<br>• mRNA-1273: 79,456 | Y | N | N | N | N |

|  | Study characteristics |  |  |  |  |  |  |  |  | Outcomes reported |  |  |  |  |
| --- | --- | --- | --- | --- | --- | --- | --- | --- | --- | --- | --- | --- | --- | --- |
| Author, year | Design | Country and data source | Age group(s) | CEV group 1/2 | CEV group 3 | SARS CoV-2 testing method | No. Vaccine doses | Study period | Vaccinated, n | Infection | Symptomatic infection | Severe infection<sup>a</sup> | Hospitalization | Death |
|  |  | Ministry of Public Health |  |  |  |  |  |  |  |  |  |  |  |  |
| Chico-Sanchez 2022 (59) | • Test-negative case control | Spain<br>Health Information Systems Analysis Service of the Ministry of Universal Health and Public Health | • ≥60 y | Y | Y | PCR and/or antigen test | 2 doses (MM vs PP) | Jan 2021–Jul 2021 | BNT162b2: 264<br>mRNA-1273: 32 | Y | N | N | N | N |
| Grewal 2022 (41) | • Test-negative case control | Canada<br>Provincial databases | • ≥60 y | Y | Y | RT-PCR test | 3 doses (MMM vs PPP) | Dec 2021–Apr 2022 | BNT162b2: 48,706<sup>b</sup><br>mRNA-1273: 57,604<sup>b</sup> | Y | Y | Y | N | N |
| Hatfield 2022 (43) | • Retrospective cohort study | USA<br>Data from 105 nursing homes | • ≥50 y | Y | Y | RT-PCR and/or antigen test, or diagnostic code | 2 doses (MM vs PP) | Dec 2020–Nov 2021 | Pre-Delta period:<br>BNT162b2: 1196<br>mRNA-1273: 466<br>Delta period:<br>BNT162b2: 687<br>mRNA-1273: 409 | Y | N | N | N | N |
| Kelly 2022 (44) | • Retrospective cohort study | USA<br>Department of VA Corporate Data Warehouse and COVID19 Shared Data Resource | • ≥65 y | N | N | Laboratory confirmed test, method not specified | 3 doses (MMM vs PPP) | Jul 2021–May 2022 | BNT162b2: 83,998<br>mRNA-1273: 100,751 | Y | Y | Y | N | N |
| Kissling 2022 (61) | • Test negative design | Europe<br>Medical records care/community study | • ≥60 y | Y | Y | PCR or antigen test | 2 doses (MM vs PP) | Jul 2021–Aug 2021 | BNT162b2: 2949<br>mRNA-1273: 263 | Y | Y | N | N | N |
| Author, year | Design | Country and data source | Age group(s) | CEV group 1/2 | CEV group 3 | SARS CoV-2 testing method | No. Vaccine doses | Study period | Vaccinated, n | Infection | Symptomatic infection | Severe infection<sup>a</sup> | Hospitalization | Death |
|  |  | sites, questionnaire and vaccine registry linkage |  |  |  |  |  |  |  |  |  |  |  |  |
| Lin 2022 (51) | • Retrospective cohort study | • USA<br>• NC COVID | Overall:<br>• ≥50 y<br>Subgroup:<br>• ≥65 y | ND | ND | Laboratory confirmed test, method not specified | 2 doses (MM vs PP) | Dec 2020–Sep 2021 | ≥50 y:<br>• BNT162b2: 1,474,746<br>• mRNA-1273: 1,379,569<br>≥65 y:<br>• BNT162b2: 694,655<br>• mRNA-1273: 734,228 | Y | N | Y | Y | Y |
| Lytras 2022 (54) | • Retrospective cohort study | • Greece<br>• Active surveillance and vaccination registry | • 60–79 y | ND | ND | PCR or antigen test | 2 doses (MM vs PP) | Jan 2021–Dec 2021 | • ND | N | N | Y | N | Y |
| Martinez-Baz 2021 (60) | • Prospective dynamic cohort study | • Spain<br>• Regional vaccination register | • ≥60 y | ND | ND | RT-PCR and/or antigen test | 2 doses (MM vs PP) | Apr 2021–Aug 2021 | • BNT162b2: 2109<br>• mRNA-1273: 215 | Y | N | N | N | N |
| Moline 2021 (45) | • Retrospective cohort study | • USA<br>• COVID-NET | Overall:<br>• ≥65 y<br>Subgroup:<br>• ≥75 y | ND | ND | Laboratory confirmed test, method not specified | 2 doses (MM vs PP) | Feb 2021–Apr 2021 | ≥65 y:<br>• BNT162b2: 258<br>• mRNA-1273: 112<br>≥75 y:<br>• BNT162b2: 185<br>• mRNA-1273: 56 | N | N | Y | Y | N |
| Nguyen 2023 (52)<sup>c</sup> | • Retrospective cohort study | • USA<br>• Integrated real-world electronic health record data set (Veradigm Health Insights), | Overall:<br>• ≥65 y<br>Subgroup:<br>• ≥75 y | Y | Y | PCR or antigen test | 3 doses (MMM vs PPP) | Feb 2021–Jan 2022 | ≥65 y:<br>• BNT162b2: 45,285<br>• mRNA-1273: 45,285<br>≥75 y:<br>• BNT162b2: 11,404<br>• mRNA-1273: 11,404 | Y | N | N | Y | N |
| Author, year | Design | Country and data source | Age group(s) | CEV group 1/2 | CEV group 3 | SARS CoV-2 testing method | No. Vaccine doses | Study period | Vaccinated, n | Infection | Symptomatic infection | Severe infection<sup>a</sup> | Hospitalization | Death |
|  |  | pharmacy and medical claims data |  |  |  |  |  |  |  |  |  |  |  |  |
| Puranik 2022 (46) | • Retrospective cohort and test-negative case-control analysis | USA<br>Health records (Mayo Clinic Health System) | Overall:<br>≥50 y<br>Subgroups:<br>≥65 y<br>≥75 y | ND | ND | PCR test | 2 doses (MM vs PP) | Dec 2020–Sep 2021 | ≥50 y:<br>BNT162b2: 7119<br>mRNA-1273: 4105<br>≥65 y:<br>BNT162b2: 4,478<br>mRNA-1273: 2,878<br>≥75 y:<br>BNT162b2: 2,249<br>mRNA-1273: 1,148 | Y | Y | N | N | N |
| Robles-Fontan 2022 (47) | • Retrospective cohort study | USA<br>National level data from Department of Health databases (BioPortal and Electronic Immunization System) | Overall:<br>≥55 y<br>Subgroups:<br>≥65 y<br>≥ 75 y | ND | ND | Laboratory confirmed test, method not specified | 2 doses (MM vs PP) | Dec 2020–Oct 2021 | ≥55 y:<br>BNT162b2: 453,015<br>mRNA-1273: 402,102<br>≥65 y:<br>BNT162b2: 260,344<br>mRNA-1273: 262,626<br>≥75 y:<br>BNT162b2: 112,715<br>mRNA-1273: 116,566 | N | N | Y | Y | Y |
| Rosenberg 2022 (48) | • Surveillance-based cohort | USA<br>Data from databases linked to cohort (CIR, NYSIIS, ECLRS, HERDS) | Overall:<br>≥50 y<br>Subgroup:<br>≥65 y | ND | ND | PCR and/or antigen test | 2 doses (MM vs PP) | May 2021–Sep 2021 | ≥50 y:<br>BNT162b2: 1,793,698<br>mRNA-1273: 1,614,377<br>≥65 y:<br>BNT162b2: 968,198<br>mRNA-1273: 1,006,002 | Y | N | Y | Y | N |
| Author, year | Design | Country and data source | Age group(s) | CEV group 1/2 | CEV group 3 | SARS CoV-2 testing method | No. Vaccine doses | Study period | Vaccinated, n | Infection | Symptomatic infection | Severe infection<sup>a</sup> | Hospitalization | Death |
| Starrfelt 2022 (57) | • Retrospective cohort study | • Norway<br>• Linked data from Norwegian National Preparedness Register for COVID-19 + six different registries | • ≥65 y | Y | Y | PCR test | 3 doses (MMM vs PPP) | Jul 2021–Nov 2021 | • ND | Y | N | Y | Y | N |
| Thompson 2021 (49) | • Test-negative case-control study | • USA<br>• Data from the electronic records from the seven VISION US network | • ≥50 y | Y | Y | RT-PCR test | 2 doses (MM vs PP) | Jan 2021–Jun 2021 | • BNT162b2: 8,500<br>• mRNA-1273: 6,374 | Y | N | N | N | N |
| van Ewijk 2022 (58) | • Test-negative case-control study | • Netherlands<br>• Data from Public Health Service testing facilities | • ≥50 y | Y | Y | LFAT or RT-PCR or LAMP test | 2 doses (MM vs PP) | Jul 2021–Dec 2021 | • BNT162b2: 2,542<br>• mRNA-1273: 273 | Y | N | N | N | N |
| Vokó 2022 (56) | • Retrospective cohort study (nationwide cohort study) | • Hungary<br>• Data from National Public Health Centre | • ≥65 y | Y | Y | PCR and/or antigen test | 3 doses (MMM vs PPP) | Sep 2021–Dec 2021 | • ND | Y | N | Y | Y | Y |
| Vokó 2022a (55) | • Retrospective cohort study (nationwide cohort study) | • Hungary<br>• Data from National Public Health Centre | Overall:<br>• ≥55 y<br>Subgroups:<br>• ≥65 y<br>• ≥75 y | ND | ND | PCR and/or antigen test | 2 doses (MM vs PP) | Jan 2021–Jun 2021 | ≥55 y<br>• BNT162b2: 845,906<br>• mRNA-1273: 116,247<br>≥65 y:<br>• BNT162b2: 613,035<br>• mRNA-1273: 80,521<br>≥75 y: | Y | N | Y | N | Y |

| Study characteristics |  |  |  |  |  |  |  |  |  | Outcomes reported |  |  |  |  |
| --- | --- | --- | --- | --- | --- | --- | --- | --- | --- | --- | --- | --- | --- | --- |
| Author, year | Design | Country and data source | Age group(s) | CEV group 1/2 | CEV group 3 | SARS CoV-2 testing method | No. Vaccine doses | Study period | Vaccinated, n | Infection | Symptomatic infection | Severe infection<sup>a</sup> | Hospitalization | Death |
|  |  |  |  |  |  |  |  |  | • BNT162b2: 302,956<br>• mRNA-1273: 41,403 |  |  |  |  |  |
| Weng 2023 (50) | • Cohort study | • USA<br>• Data from a major FQHC in Rhode Island | • ≥55 y | ND | ND | RT-PCR test | 2 doses (MM vs PP) | Jan 2021–Dec 2021 | • ND | Y | N | N | N | N |

CEV, clinically extremely vulnerable; CIR, Citywide Immunization Registry; COVID-NET, COVID-19–Associated Hospitalization Surveillance Network; ECLRS, Electronic Clinical Laboratory Reporting System; FQHC, federally qualified health center; HERDS, Health Electronic Response Data System; LAMP, loop-mediated isothermal amplification; LFAT, lateral-flow antigen test; N, no; NC COVID, North Carolina COVID-19 Surveillance System; ND, not disclosed; NYSIIS, New York City, and the New York State Immunization Information System; PCR, polymerase chain reaction; RT, reverse transcription; SISVER, Sistema de Vigilancia Epidemiológica de Enfermedades Respiratorias (nationwide sentinel surveillance system); VA, Veterans Affairs; Y, yes.
Vaccine dosing abbreviated as MM or MMM for 2 or 3 doses of mRNA-1273, respectively, and PP or PPP for 2 or 3 doses of BNT162b2, respectively.
<sup>a</sup>Derived severe infections (based on either severe infection as defined in the study or hospitalization data or death data).
<sup>b</sup>Number of vaccinated participants included in the infection analysis. For the symptomatic infection analysis, n=2139 (BNT162b2) and n=1831 (mRNA-1273); for the severe infection analysis, n=1,638 (BNT162b2) and n=1,518 (mRNA-1273).
<sup>c</sup>Industry sponsored study (Moderna, Inc).

Based on the risk of bias assessment for nonrandomized studies, most of the studies included in the meta-analysis had no serious risk of bias; however there was serious risk of bias in four studies (40, 45, 54, 59), and risk of bias was not estimable for one study (50) (**Supplemental Table S4**).

### SARS-CoV-2 infection

In meta-analysis of 22 studies reporting the outcome of SARS-CoV-2 infection in older adults aged ≥50 years, vaccination with mRNA-1273 was associated with significantly lower risk of SARS-CoV-2 infection compared with vaccination with BNT162b2 (RR 0.72 [95% CI 0.64–0.80]; **Table 2** and **Figures 2 and 3**). The RD was estimated as 442 fewer (95% CI 570 fewer to 313 fewer) SARS-CoV-2 infections per 100,000 people vaccinated. There was considerable heterogeneity between the studies (RR I^2^=94.4%; RD, I^2^=98.4%). The certainty of evidence was graded as type 4 (very low) due to imprecision and indirectness resulting from the varying outcome definitions used for infection and inclusion of non-randomized studies (**Table 2**). In a sensitivity analysis using the second-order methodological approach, vaccination with mRNA-1273 was associated with significantly fewer SARS-CoV-2 infections compared with BNT162b2 (RR 0.72 [95% CI 0.62–0.85]; I^2^=0%), consistent with the base case analysis (**Figure 4A** and **Supplemental Figure S2A**).

**Figure 2.**
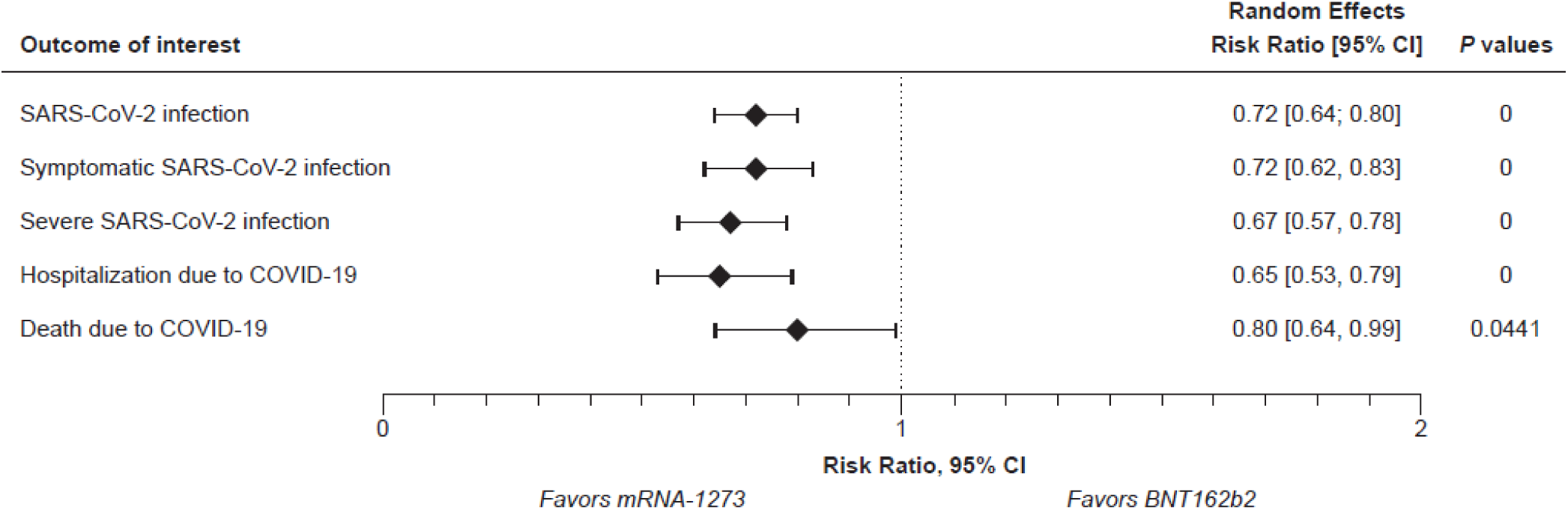
Summary of meta-analysis results on clinical effectiveness outcomes of the mRNA-1273 versus BNT162b2 COVID-19 vaccines in the overall population of older adults aged ≥50 years.

**Figure 3.**
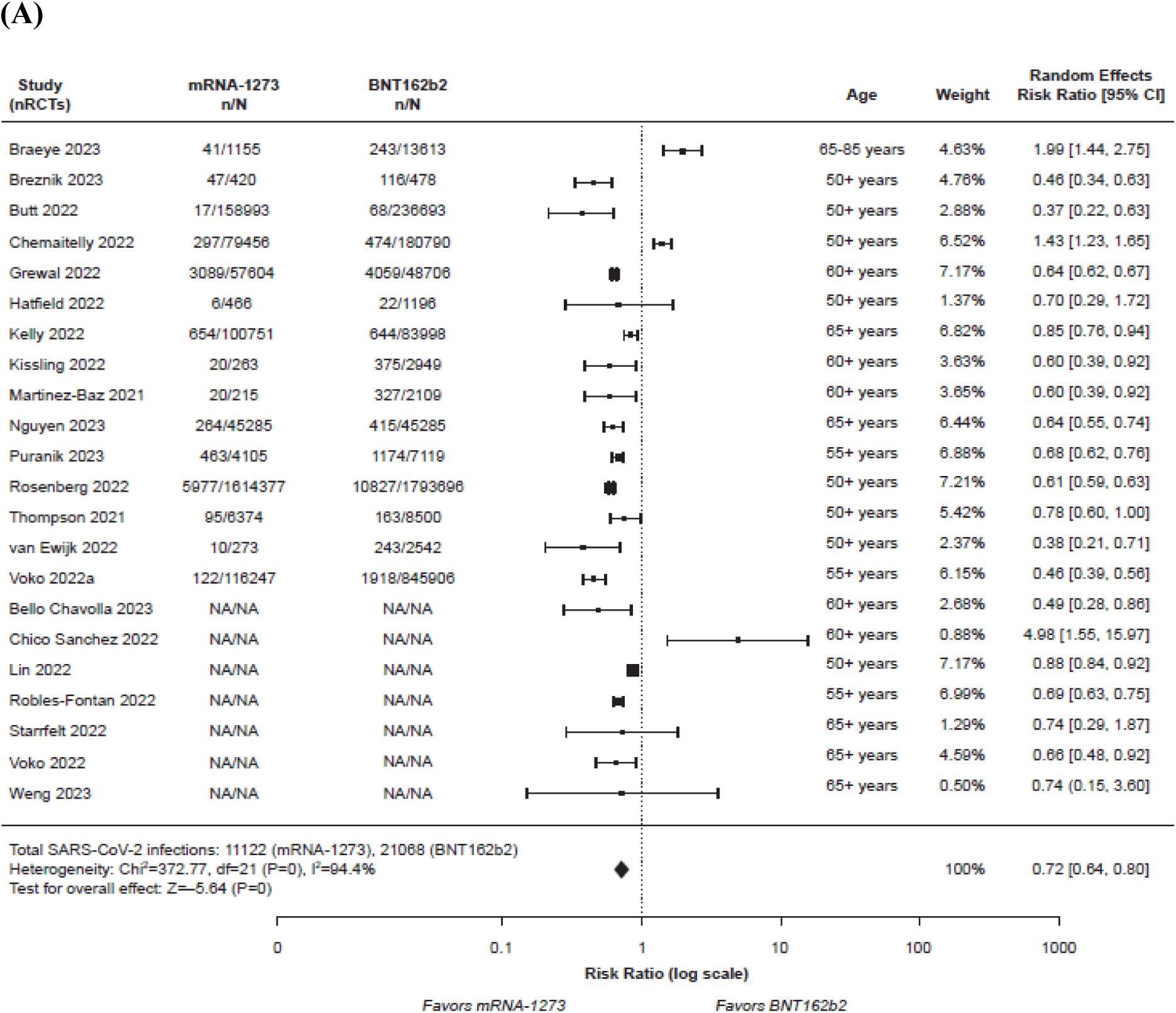
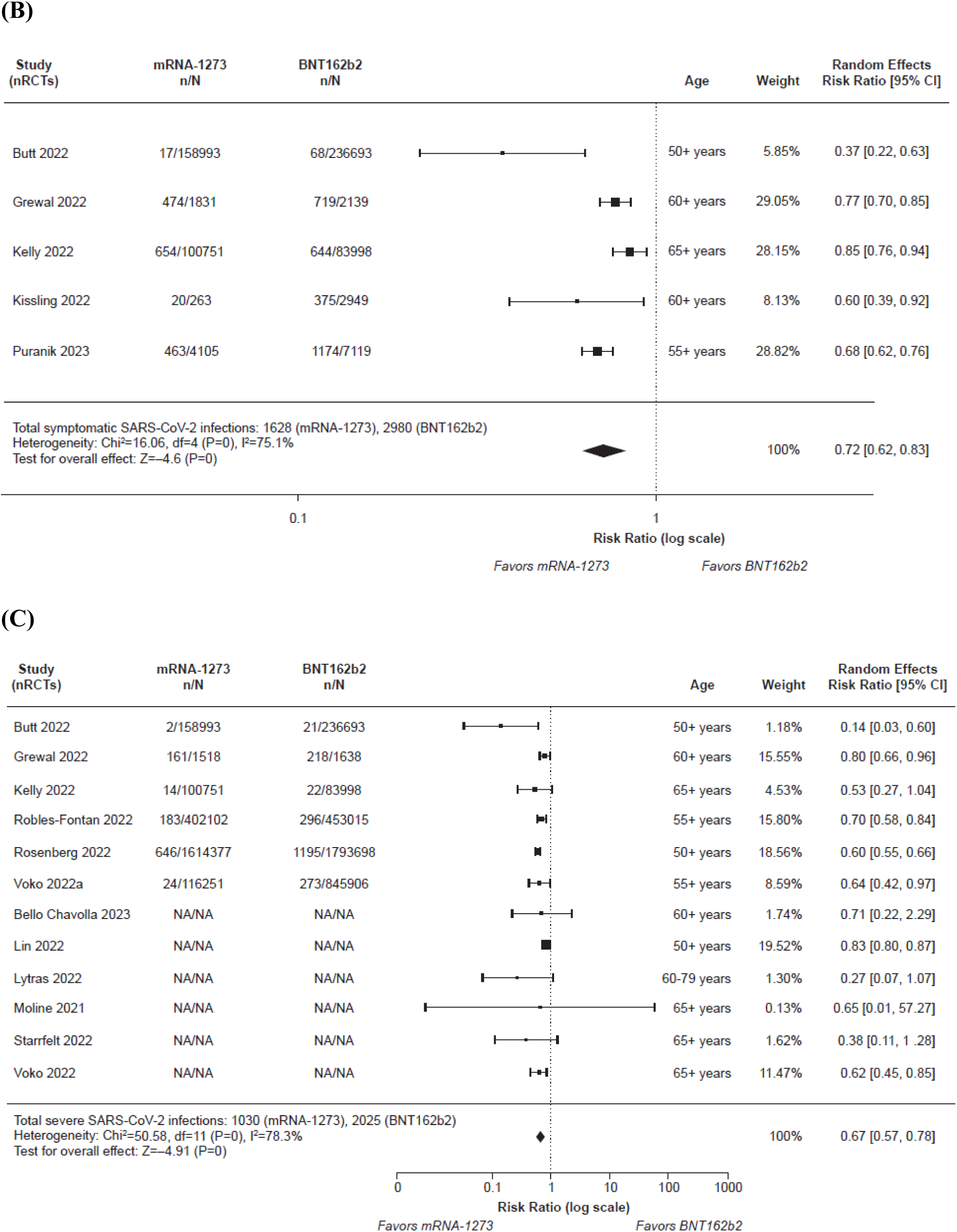
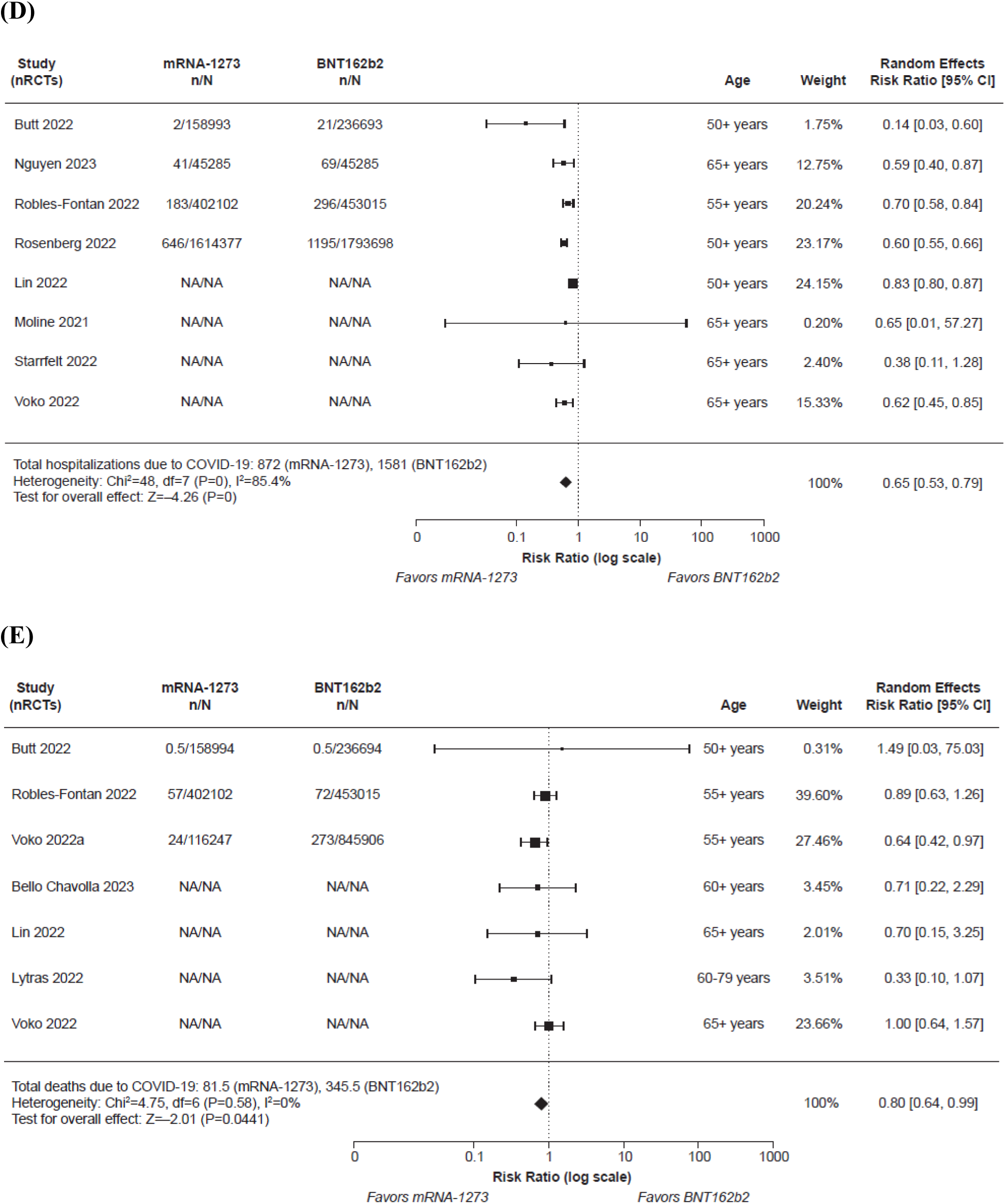
Meta-analysis results comparing the mRNA-1273 versus BNT162b2 COVID-19 vaccines in the overall population of older adults aged ≥50 years by study for (A) SARS-CoV-2 infection; (B) laboratory-confirmed symptomatic SARS-CoV-2 infection; (C) severe SARS-CoV-2 infection; (D) hospitalization due to COVID-19; and (E) death due to COVID-19.

**Figure 4.**
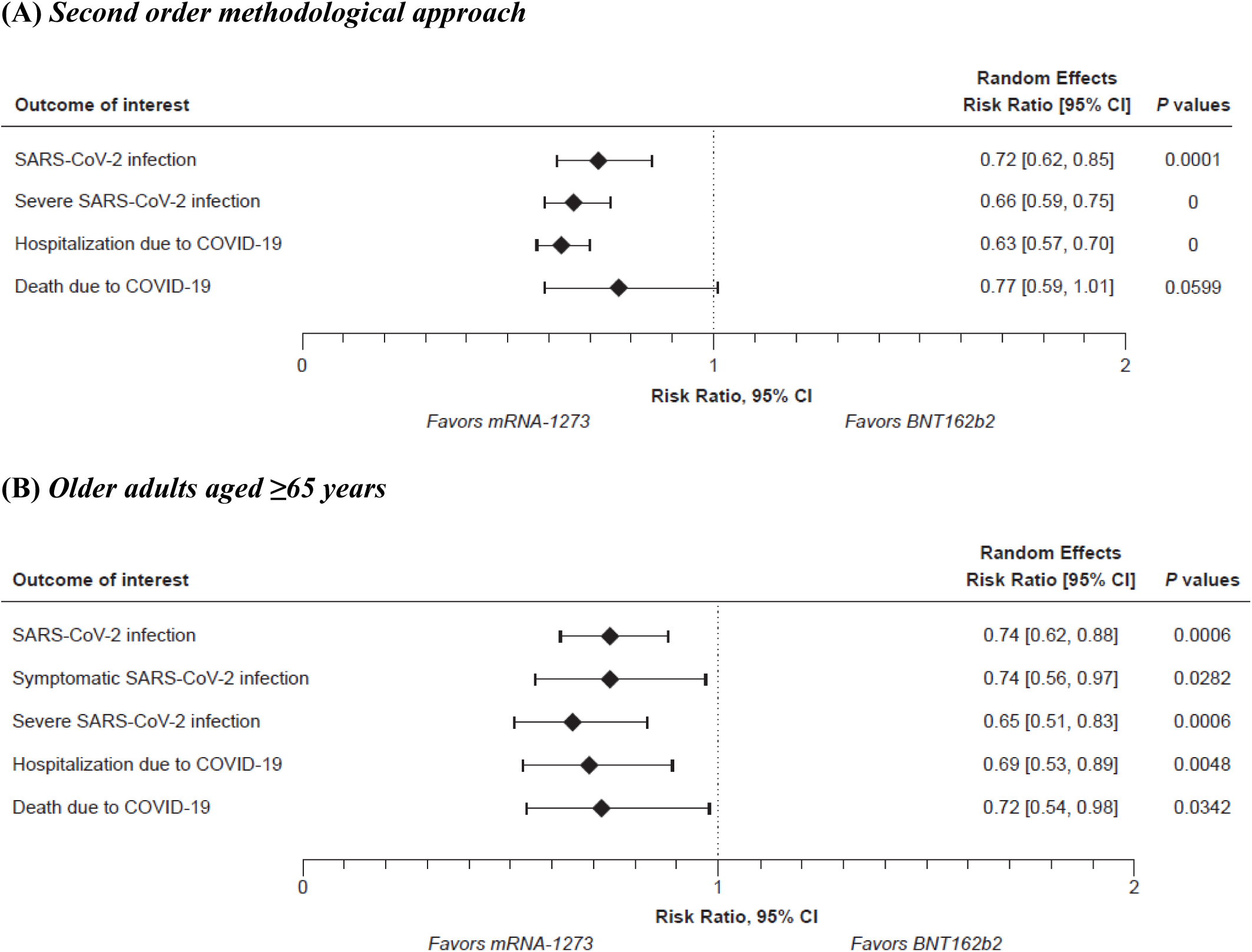
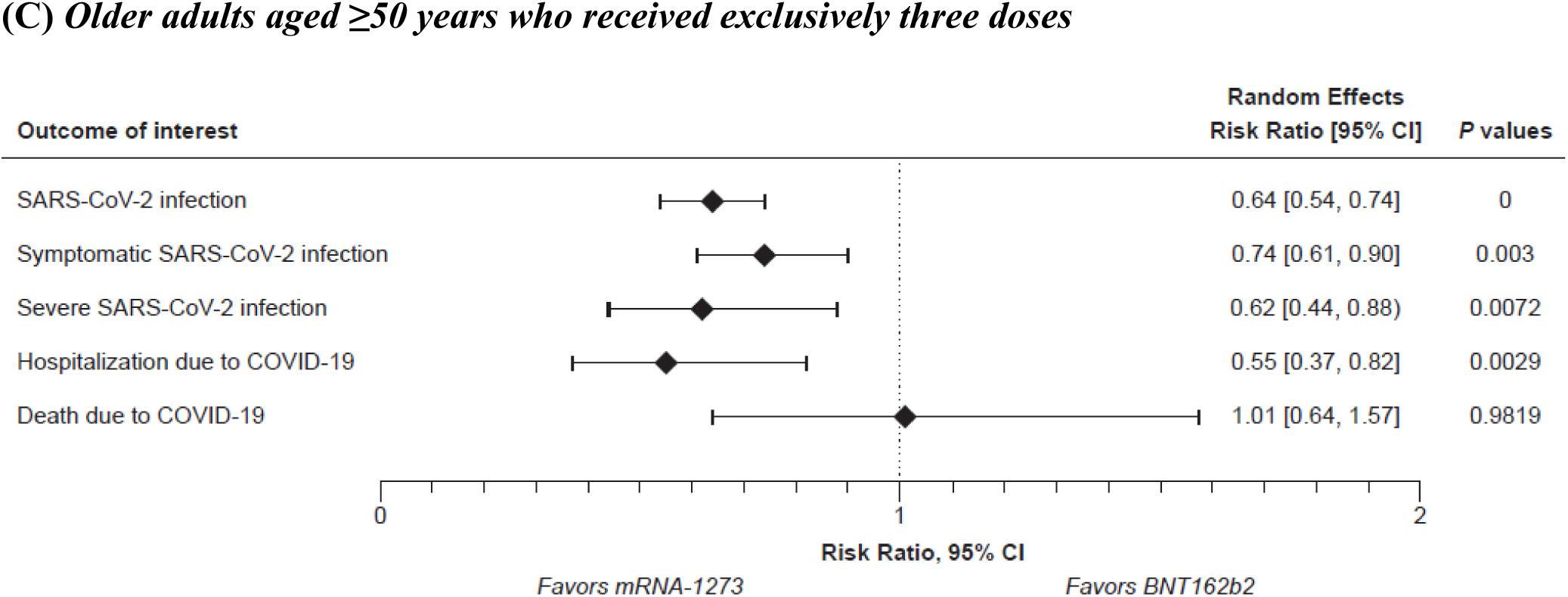
Summary of sensitivity meta-analyses on clinical effectiveness outcomes of the mRNA-1273 versus BNT162b2 COVID-19 vaccines (A) using the second order methodological approach^a^ and in subgroups of (B) older adults aged ≥65 years; and (C) older adults aged ≥50 years who received exclusively 3 doses. ^a^Results of the second-order methodological approach for the outcome of symptomatic infection are not presented because the results are identical to the results of the main analysis (no conversion was necessary for this outcome in the main analysis).

**Table 2.**
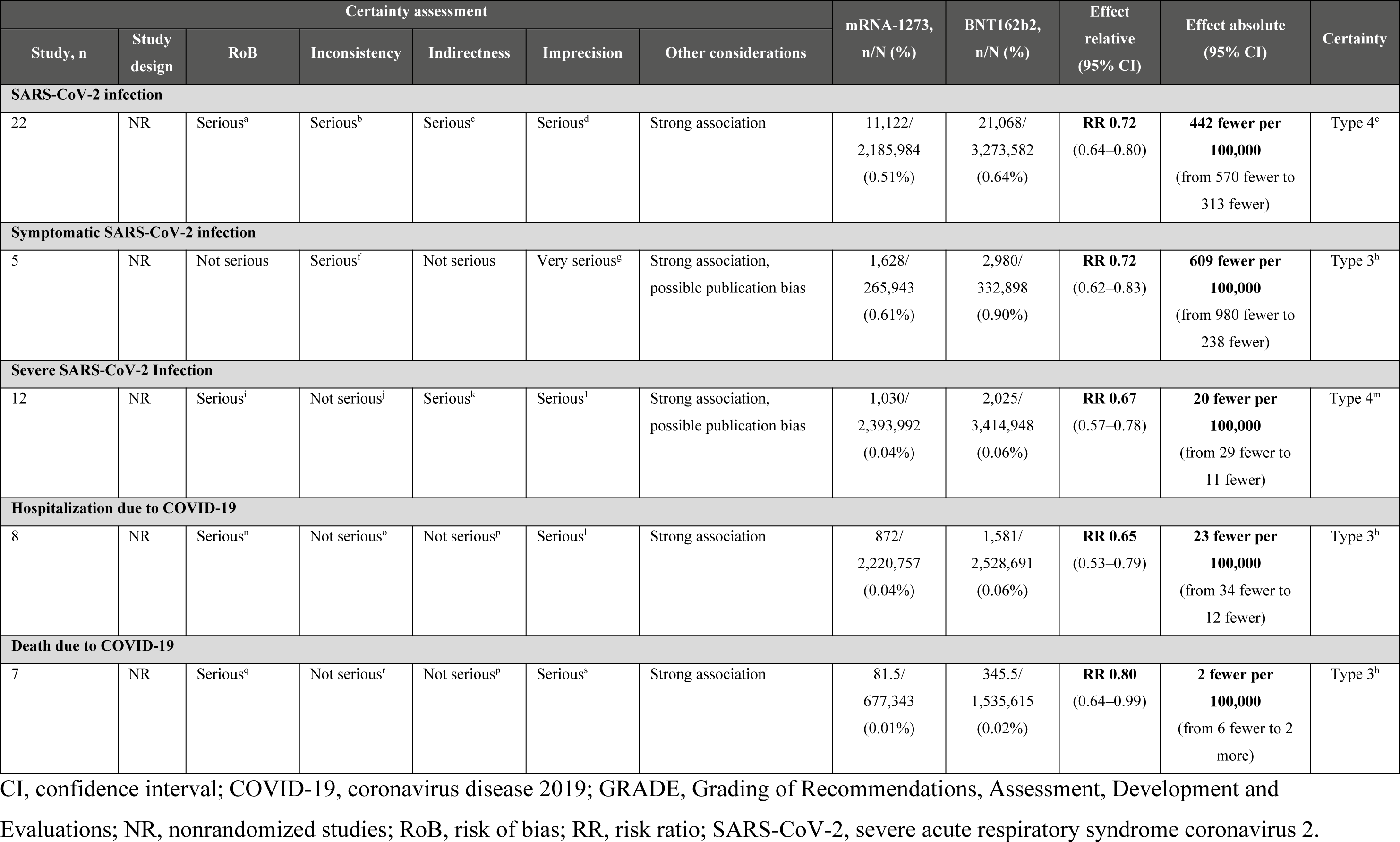
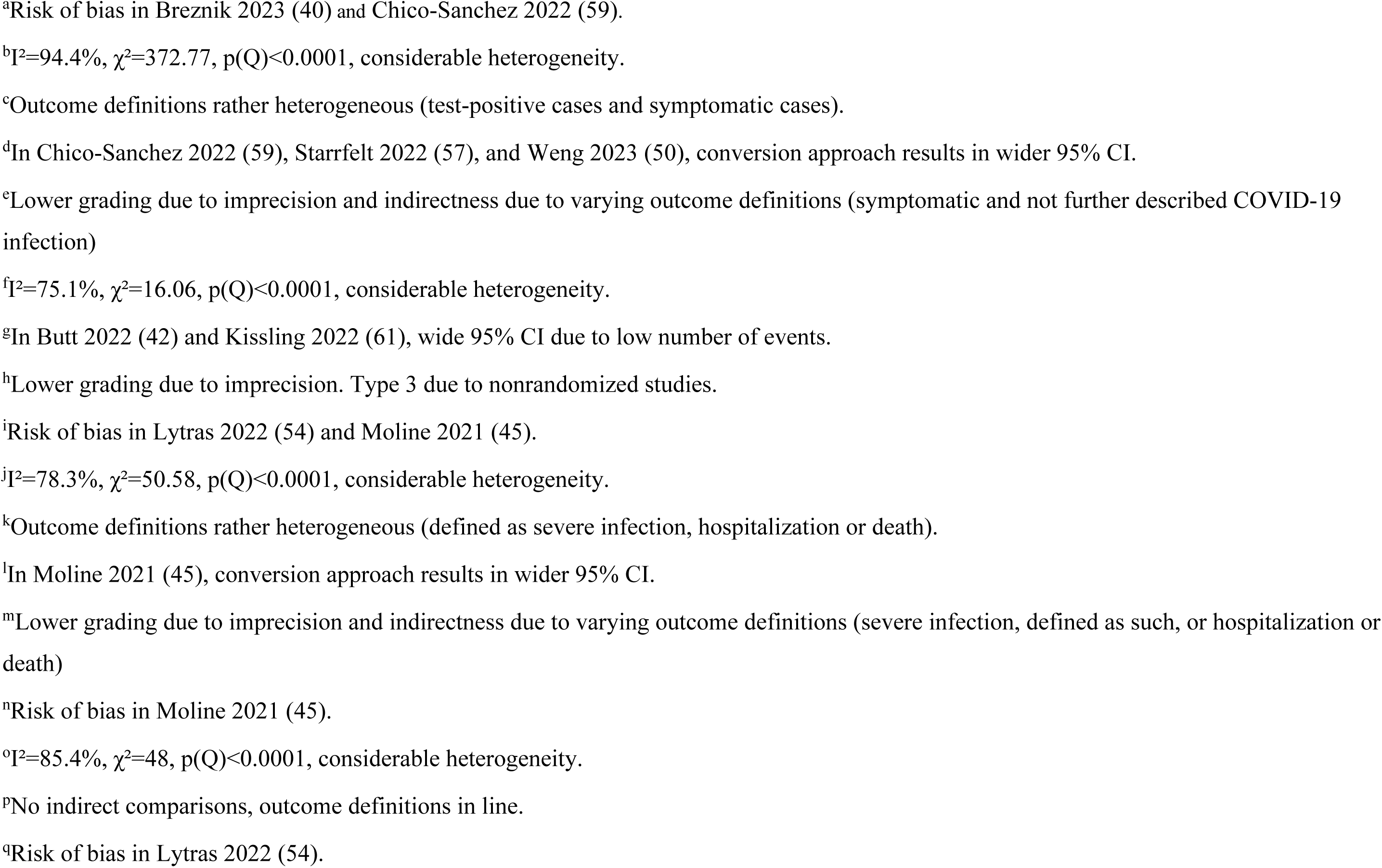
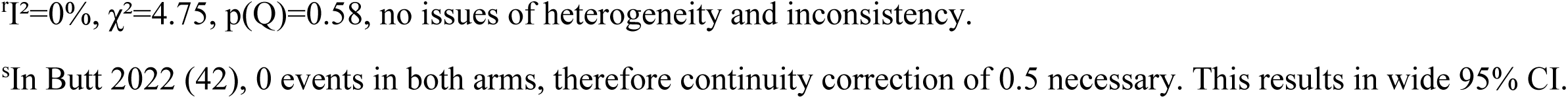
Summary of overall GRADE findings.

In a subgroup analysis of 10 studies reporting the outcome of SARS-CoV-2 infection in adults aged ≥65 years, mRNA-1273 vaccination was also associated with significantly fewer infections compared with BNT162b2 vaccination (RR 0.74 [95% CI 0.62–0.88]; RD 216 fewer cases per 100,000 vaccinated [95% CI 333 fewer to 100 fewer]; **Table 3**, **Figure 4B** and **Supplemental Figure S3A**). Subgroup analysis of seven studies reporting this outcome in individuals aged ≥50 years who received exclusively three vaccine doses also found that mRNA-1273 was associated with fewer infections versus BNT162b2 (RR 0.64 [95% CI 0.54–0.74]; RD 1098 fewer cases per 100,000 vaccinated [95% CI 1535 fewer to 661 fewer; **Table 3**, **Figure 4C** and **Supplemental Figure S4A**). As in the overall population analysis, the certainty of evidence in these two subgroups was graded as type 4 (very low) due to imprecision and varying outcome definitions (**Table 3**), and there was considerable heterogeneity between studies (RR I^2^=89.5% for the ≥65 years of age subgroup; I^2^=80.8% for the 3-dose subgroup). Additional subgroup analyses in older adults aged ≥75 years; in older adults aged ≥50 years, excluding individuals with CEV group 1 and 2 conditions; in older adults aged ≥50 years infected with the Delta variant; and excluding those studies that only included VE data were generally consistent with the findings from the overall meta-analysis (**Supplemental Figures S5A, S6A, S7A, and S8A**).

**Table 3.**
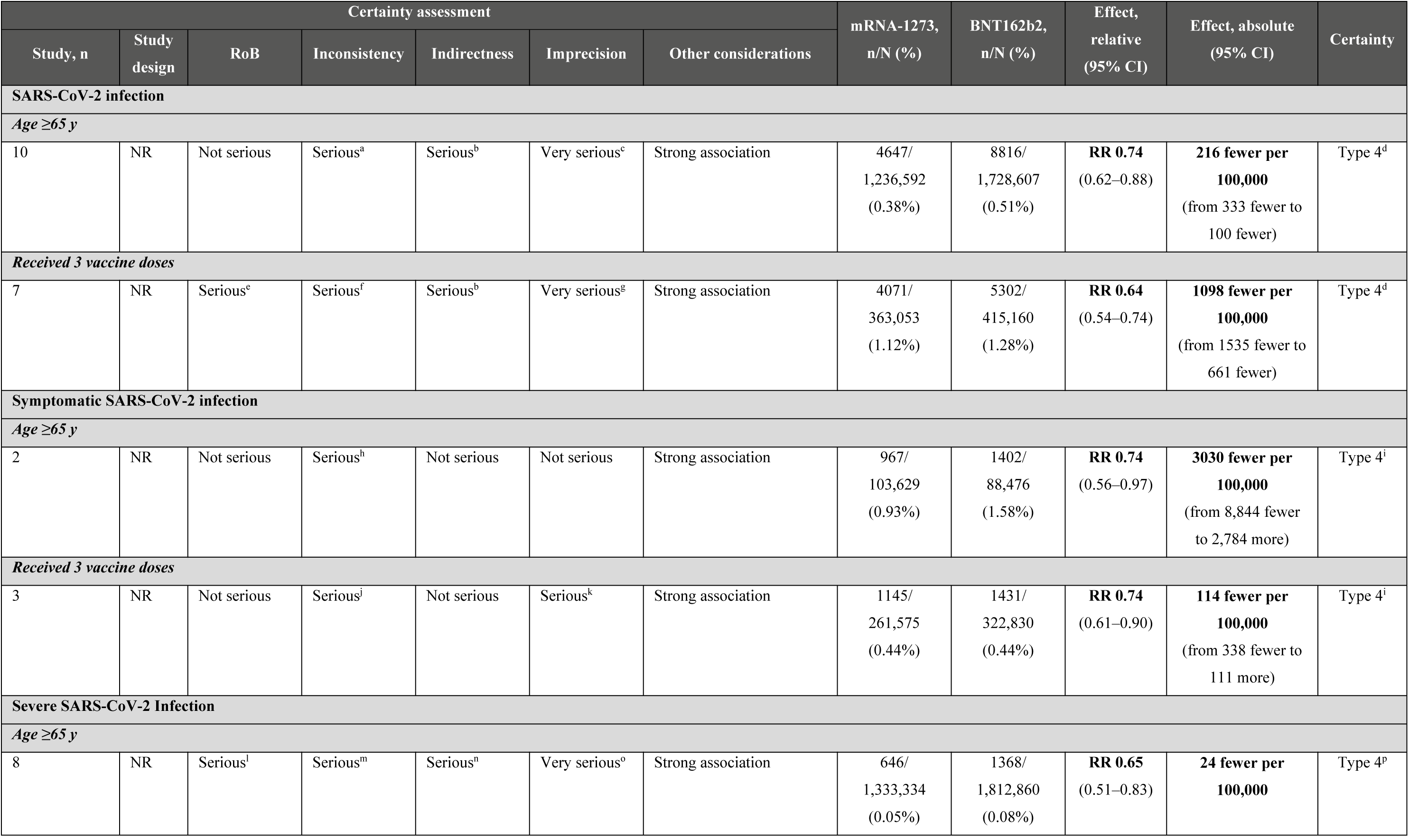
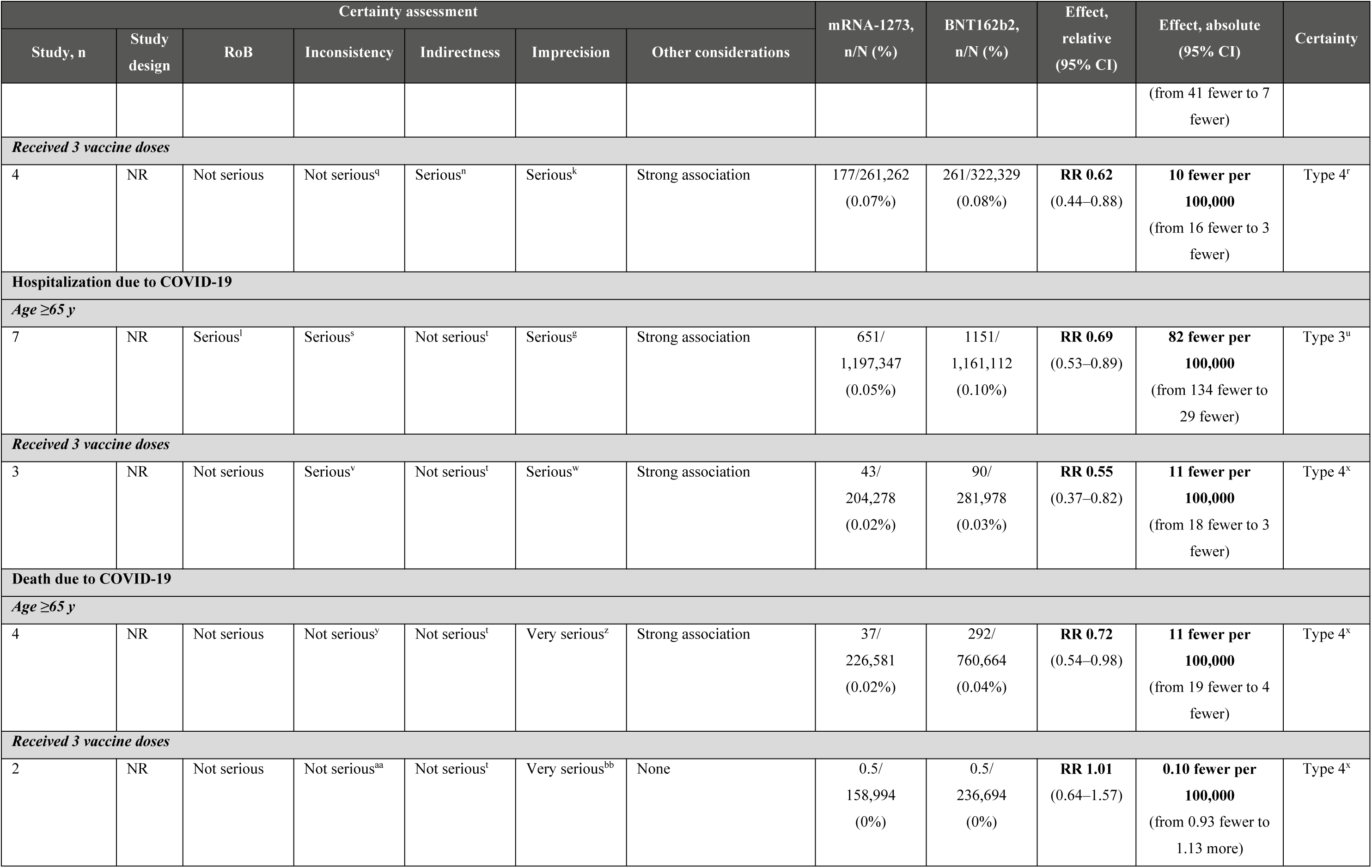
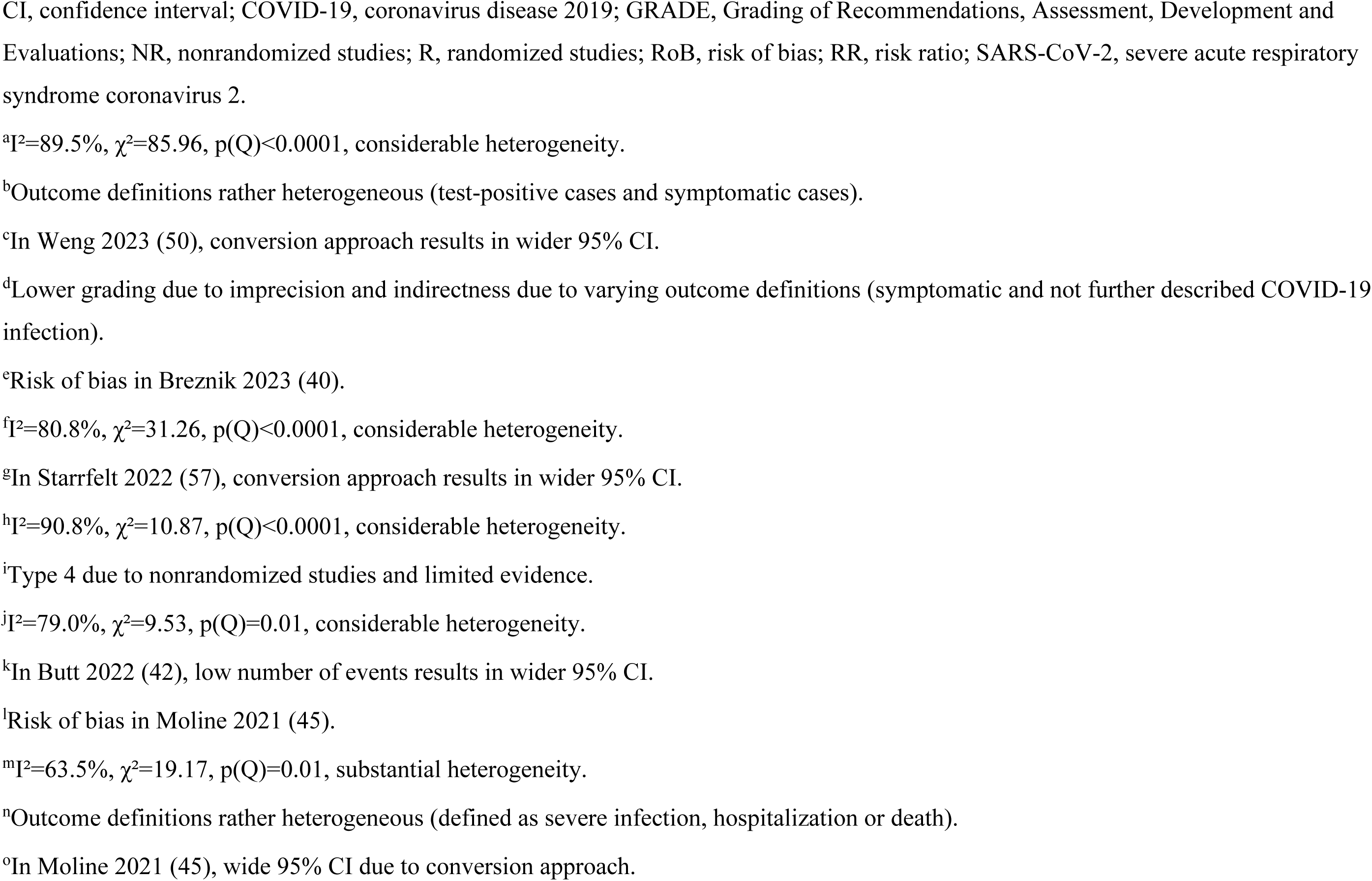
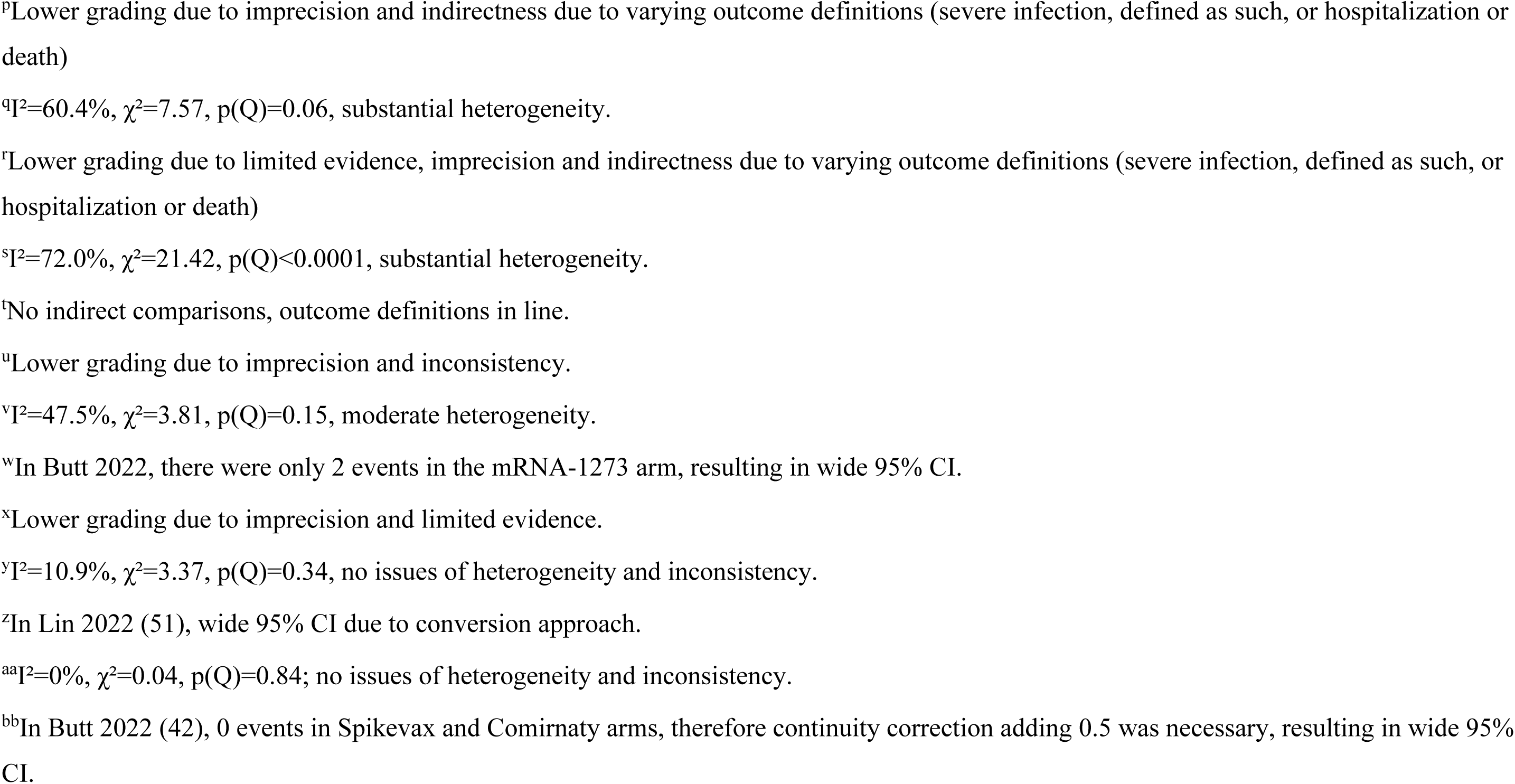
Summary of GRADE findings by population subgroup.

### Laboratory-confirmed symptomatic SARS-CoV-2 infection

Five studies were included in the meta-analysis of laboratory-confirmed symptomatic SARS-CoV-2 infection in individuals aged ≥50 years (**Table 2**). Vaccination with mRNA-1273 was associated with significantly fewer SARS-CoV-2 symptomatic infections versus vaccination with BNT162b2 (RR 0.72 [95% CI 0.62–0.83; **Figure 2 and 3**). The RD was estimated as 609 fewer symptomatic infections per 100,000 individuals vaccinated (95% CI 980 fewer to 238 fewer cases). Heterogeneity between studies was also considerable for this outcome (RR I^2^=75.1%; RD I^2^=96.2%). The certainty of evidence was graded as type 3 (low) due to imprecision, with a lower grading assigned due to inclusion of non-randomized studies (**Table 2**). Possible publication bias was noted for this outcome based on Egger’s regression test (*P* < 0.05) **(Supplemental Figure S9B)**. Because no VE data were used in the base case meta-analysis of symptomatic SARS-CoV-2 infections, no conversion was necessary. Therefore, results from the second-order methodological approach were identical to the base case results presented in **Figure 2 and 3**.

Subgroup analysis based on two studies in individuals aged ≥65 years also found significantly reduced risk of symptomatic SARS-CoV-2 infections with mRNA-1273 versus BNT162b2 vaccination (RR 0.74 [95% CI 0.56–0.97]; RD 3,030 fewer cases per 100,000 vaccinated [95% CI 8,844 fewer to 2,784 more cases]; **Table 3**, **Figure 4B** and **Supplemental Figure S3B**). Similarly, in meta-analysis of three studies that included individuals aged ≥50 years who received exclusively three doses of vaccine, mRNA-1273 was associated with lower risk of symptomatic infections compared with BNT162b2 (RR 0.74 [95% CI 0.61–0.90]; RD 114 fewer cases per 100,000 individuals vaccinated [95% CI 338 fewer to 111 more]; **Table 3**, **Figure 4C, Supplemental Figure S4B**). As in the overall meta-analysis, heterogeneity between studies was considerable for these subgroups (RR I^2^=90.8% and 79.0%, respectively). The certainty of evidence was graded as type 4 (very low) for both subgroups (**Table 3**). Results of additional subgroup analyses in adults aged ≥75 years; adults aged ≥50 years, excluding individuals with CEV group 1 and 2 conditions; and adults aged ≥50 years infected with the Delta variant were generally consistent with the findings from the overall meta-analysis (**Supplemental Figures S5B, S6B, and S7B**). There were no studies evaluating the outcome of lab-confirmed symptomatic SARS-CoV-2 infection that exclusively reported VE data.

### Severe SARS-CoV-2 infection

Based on meta-analysis of 12 studies, vaccination with mRNA-1273 was associated with significantly fewer severe SARS-CoV-2 infections compared with vaccination with BNT162b2 (RR 0.67 [95% CI 0.57–0.78]; **Table 2** and **Figure 2 and 3**). This result corresponds to an estimated RD of 20 fewer severe infections per 100,000 individuals vaccinated with mRNA-1273 versus BNT162b2 (95% CI 29 fewer to 11 fewer cases). There was considerable heterogeneity across studies for this outcome (RR I^2^=78.3%; RD I^2^=86.1%). Evidence certainty was graded as type 4 (very low) due to imprecision and varying definitions used for severe infection (defined as severe infection, or hospitalization, or death; **Table 2**). Possible publication bias was noted for this outcome based on Egger’s regression test (*P* < 0.05; **Supplemental Figure S9C**). Consistent with the findings from the base case analysis, sensitivity analysis using the second-order methodological approach also found that vaccination with mRNA-1273 was associated with significantly fewer severe SARS-CoV-2 infections compared with vaccination with BNT162b2 in older adults aged ≥50 years (RR 0.66 [95% CI 0.59–0.75]; **Figure 4A** and **Supplemental Figure S2B**).

In subgroup analyses, mRNA-1273 was associated with significantly fewer severe SARS-CoV-2 infections compared with vaccination with BNT162b2 in older adults aged ≥65 years (eight studies; RR 0.65 [95% CI: 0.51–0.83]; RD 24 fewer severe infections per 100,000 individuals vaccinated [95% CI 41 fewer to 7 fewer]) and in adults aged ≥50 years who received exclusively three vaccine doses (four studies; RR 0.62 [95% CI 0.44–0.88]; RD 10 fewer severe infections per 100,000 individuals vaccinated [95% CI 16 fewer to 3 fewer]; **Table 3**, **Figure 4B and 4C**, and **Supplemental Figure S3C and S4C**). There was substantial heterogeneity across studies for both subgroups (RR I^2^=63.5% and 60.4%, respectively). Evidence certainty was graded as type 4 (very low; **Table 3**). Similar to the findings from the overall meta-analysis, mRNA-1273 was associated with reduced risk of severe SARS-CoV-2 infection in additional subgroup analyses of individuals ≥75 years of age, individuals (aged ≥50 years) without CEV group 1 or 2 conditions, individuals (aged ≥50 years) infected with the Delta variant, and in the subgroup excluding those studies that only included VE data (**Supplemental Figures S5C, S6C, S7C, and S8B**).

### Hospitalization due to COVID-19

Based on a meta-analysis of eight studies, vaccination with mRNA-1273 was associated with significantly lower risk of hospitalization due to COVID-19 in individuals aged ≥50 years compared with vaccination with BNT162b2 (RR 0.65 [95% CI 0.53–0.79]; **Table 2** and **Figure 2 and 3**). The estimated RD was 23 fewer COVID-19 hospitalizations per 100,000 individuals vaccinated (95% CI 34 fewer to 12 fewer). Heterogeneity across studies was considerable (RR I^2^=85.4%; RD I^2^=90.3%). The certainty of evidence grade was type 3 (low) for this outcome, due to imprecision and inclusion of non-randomized studies (**Table 2**). The sensitivity analysis using the second-order methodological approach found that vaccination with mRNA-1273 was associated with significantly fewer COVID-19‒related hospitalizations compared with vaccination with BNT162b2 (RR 0.63 [95% CI 0.57–0.70]), consistent with the base case analysis (**Figure 4A** and **Supplemental Figure S2C**).

Based on seven studies of COVID-19‒related hospitalization in the subgroup of older adults aged ≥65 years, vaccination with mRNA-1273 was associated with significantly reduced risk of hospitalization compared with vaccination with BNT162b2 (RR 0.69 [95% CI 0.53–0.89]; RD 82 fewer hospitalizations per 100,000 individuals vaccinated [95% CI 134 fewer to 29 fewer]; **Table 3** and **Figure 4B** and **Supplemental Figure S3D**). As in the overall meta-analysis, there was considerable heterogeneity across studies (RR I^2^=72.0%), and the evidence certainty was graded as type 3 (low; **Table 3**). Vaccination with mRNA-1273 was also associated with significantly reduced risk of hospitalization compared with vaccination with BNT162b2 among individuals aged ≥50 years who received three vaccine doses based on meta-analysis of three studies (RR 0.55 [95% CI 0.37–0.82]; RD 11 fewer hospitalizations per 100,000 individuals vaccinated [95% CI 18 fewer to 3 fewer]; **Table 3** and **Figure 4C** and **Supplemental Figure S4D**). There was moderate heterogeneity across studies (RR I^2^=47.5%), and the evidence certainty was graded as type 4 (very low; **Table 3**). Additional subgroup analyses in adults aged ≥75 years; adults aged ≥50 years, excluding individuals with CEV group 1 and 2 conditions; adults aged ≥50 years infected with the Delta variant; and excluding those studies that only included VE data were generally consistent with the findings from the overall meta-analysis (**Supplemental Figures S5D, S6D, S7D, and S8C**).

### Death due to COVID-19

In meta-analysis of seven studies reporting mortality in individuals aged ≥50 years, vaccination with mRNA-1273 was associated with significantly lower risk of COVID-19‒ related death compared with vaccination with BNT162b2 (RR 0.80 [95% CI 0.64–0.99]). The estimated RD was 2 fewer deaths per 100,000 people vaccinated (95% CI 6 fewer to 2 more) (**Table 2** and **Figure 2** and **3**). No evidence of heterogeneity between the studies was observed in the RR analysis (I^2^=0%), although heterogeneity was moderate for the estimation of RD (I^2^=47.8%). The certainty of evidence was graded as type 3 (low) for this outcome, due to imprecision and inclusion of non-randomized studies (**Table 2**). In the sensitivity analysis using the second-order approach, mRNA-1273 vaccination was associated with numerically reduced risk of death due to COVID-19 compared with BNT162b2 vaccination, but this was not statistically significant (RR 0.77 [95% CI 0.59–1.01]; **Figure 4A** and **Supplemental Figure S2D**).

In subgroup analysis of four studies reporting this outcome in older adults aged ≥65 years, vaccination with mRNA-1273 was associated with fewer COVID-19 deaths versus vaccination with BNT162b2 (RR 0.72 [95% CI 0.54–0.98]; RD 11 fewer deaths per 100,000 individuals vaccinated [95% CI 19 fewer to 4 fewer) (**Table 3**, **Figure 4B**, and **Supplemental Figure S3E**). The evidence suggested that the heterogeneity across studies might not be important (RR I^2^=10.9%). The evidence certainty was graded as type 4 (very low) in this analysis due to imprecision and limited evidence (**Table 3**). There was no statistically significant difference between the mRNA vaccines against the outcome of COVID-19–related deaths in the subgroup of individuals aged ≥50 years who received exclusively three vaccine doses, based on analysis of two studies (RR 1.01 [95% CI 0.64– 1.57]; **Table 3**, **Figure 4C**, and **Supplemental Figure S4E**). The mRNA-1273 vaccine was associated with reduced risk of COVID-19–related death compared with BNT162b2 when individuals with CEV1/2 group conditions were excluded (**Supplementary Figure S6E**). There was no statistically significant difference in mortality risk between mRNA vaccines in subgroup analyses of individuals aged ≥75 years, individuals aged ≥50 years exposed to Delta variant, or in the subgroup excluding those studies with only VE data (**Supplemental Figures S5E, S7E, and S8D**).

## DISCUSSION

This meta-analysis of 24 studies in older adults aged ≥50 years found that vaccination with mRNA-1273 was statistically significantly associated with lower risk of SARS-CoV-2 infections, including asymptomatic, symptomatic, and severe infections, as well as hospitalizations and deaths due to COVID-19 compared with vaccination with BNT162b2. To our knowledge, this is the first such analysis of pairwise real-world evidence in adults aged 50 years or older. This evidence helps inform considerations about which vaccine to choose for older adults.

Older age has consistently been identified as a primary risk factor for worse outcomes with COVID-19 (6, 7, 8), with older adults accounting for the majority of COVID-19–related deaths (2, 3, 5, 62). This meta-analysis provides evidence for improved outcomes with the mRNA-1273 vaccine compared with the BNT162b2 vaccine in older adults. Similarly, high-dose and adjuvanted influenza vaccines have demonstrated improved outcomes over standard dose influenza vaccines in older adults (21, 22), and, as a result, these vaccines are preferentially recommended for the elderly population in many countries (63, 64). Immunology studies have also reported higher antibody production with the mRNA-1273 vaccine compared with the BNT162b2 vaccine (19).

Findings from the sensitivity analysis using the second order meta-analysis approach were consistent with the overall results among older adults aged ≥50 years, except that there was no significant difference between mRNA vaccines for the outcome of COVID-19–related deaths. Consistent findings were also observed in subgroup analyses among adults aged ≥65 years or ≥75 years, in adults aged ≥50 years who received exclusively three doses, in those who did not have CEV groups 1 and 2 conditions, and those infected by the Delta variant, and in the subgroup excluding studies that only reported VE. Across these subgroups, vaccination with mRNA-1273 was associated with significantly fewer infections, symptomatic infections, and severe infections compared with vaccination with BNT162b2. Vaccination with mRNA-1273 was also associated with significantly fewer hospitalizations compared with vaccination with BNT162b2 in each subgroup. Similar results were observed for COVID-19–related death, except that there was no significant difference observed for this outcome between the vaccines in the subgroup of adults aged ≥75 years and in those infected with the Delta variant. Overall, the findings from the base-case analysis were confirmed by a broad range of sensitivity analyses considering different subgroups as well as different methodologies (i.e., second order approach and exclusion of VE studies), suggesting that the findings of the meta-analysis are robust.

Limitations of this systematic review and meta-analysis should be considered. Because all the studies included in the analysis were observational in nature, the certainty of evidence was graded as low or very low (type 3 or below). Furthermore, four of the 24 studies included in the meta-analysis had a serious risk of bias, and risk of bias was not estimable for one additional study due to lack of sufficient information. Importantly, the tight timelines for developing variant-adapted vaccines for COVID-19 limits the feasibility of large RCTs. In this context, estimates of the comparative effectiveness of COVID-19 vaccines based on real-world evidence provides crucial information to address important clinical questions and inform policy decisions regarding vaccination (65). Nevertheless, higher quality real-world evidence studies on vaccine effectiveness are needed, particularly for the outcomes of COVID-19–related hospitalizations and deaths. Possible publication bias was noted based on Egger’s regression test (*P*<0.05) for the outcomes of symptomatic infections and severe infections. However, the number of studies reporting symptomatic infections was small (n=5), limiting the power of Egger’s regression test to accurately distinguish chance from true asymmetry. A combination of various endpoints, including severe infection (as defined by the study), hospitalization, and death, was used to define a composite severe infections outcome in this meta-analysis, introducing additional heterogeneity. This may have contributed to the significant asymmetry observed for this outcome. Publication bias was not detected based on Egger’s test for outcomes with sufficient numbers of studies in the evidence base (ie, for SARS-2-CoV-2 infections, hospitalizations, and deaths). The studies included in our meta-analysis showed a large amount of heterogeneity. This finding is possibly a reflection of the complex interactions between vaccination and contextual factors as they operate in the real world. However, such heterogeneity does introduce challenges in predicting true vaccine effectiveness under a given regimen, or for a given population. Various factors could have driven the observed heterogeneity, including differences in study populations, statistical approaches employed, definitions of outcomes (e.g., for severe COVID-19), analyzed time points after vaccination, and vaccination schedules and regimens. Such high heterogeneity may also be expected in older populations, in part due to the large heterogeneity in health status associated with underlying comorbidities, for example. Meta-regression accounting for some of the factors plausibly driving heterogeneity (such as varying time points of analysis after vaccination and vaccination schedules and regimes) could not be conducted due to sparse data. However, we performed multiple subgroup analyses to account for age differences (i.e., restricted to individuals aged ≥65 years and ≥75 years), differences in number of vaccine doses (i.e., restricted to individuals who received three doses), underlying medical conditions (i.e., excluding those with CEV group 1 and 2 conditions), and SARS-CoV-2 variant (i.e., restricted to a single variant of concern [Delta]) to better understand the source of heterogeneity. Heterogeneity continued to be observed across these sub-analyses. Notably, high heterogeneity has also been noted in meta-analyses of influenza vaccine effectiveness in older adults (21, 64, 66). Future studies and reviews examining which factors predict when, where, and for whom the vaccines show differential effectiveness would be beneficial to address possible disparities in protection. Despite the high heterogeneity we observed, comprehensive sensitivity analyses considering only subsets of studies (i.e., excluding studies or groups of studies) were conducted, results of which highlight the robustness of effect sizes and the conclusion of the overall meta-analysis (67).

Our evidence synthesis has several considerable strengths. First, we used broad search terms and high-quality systematic literature review methodology, which included training reviewers and validating the included studies and extracted data. Second, we used advanced meta-analytical methods to include both studies reporting event and participant numbers by vaccine arm as well as studies reporting only VE. This approach allowed for inclusion of all available data, taking into account both within-study and between-study variability, resulting in more robust and reliable conclusions than would be possible if either only binary data or only VE data were included. We also carried out a sensitivity analysis using a second-order meta-analytical model which demonstrated similar results to the main analysis, corroborating the robustness of the data and the analytical methods employed. Finally, this evidence synthesis and meta-analysis provides important updates compared with previous analyses, notably providing results on the comparative effectiveness of the two available mRNA vaccines in preventing SARS-CoV-2 infections and associated severe outcomes.

### Conclusions

Vaccination with mRNA-1273 was associated with significantly fewer asymptomatic, symptomatic, and severe infections; hospitalizations; and deaths due to COVID-19 than vaccination with BNT162b2 in older adults aged ≥50 years, and these differences generally persisted among subgroups of patients, including among older adults aged ≥65 years and adults aged ≥50 years who received three doses of the same vaccine. These results can inform policy makers who wish to optimize vaccination programs at the population level, as well as health care professionals making individual-based recommendations to their patients.

## Data Availability

All data produced in the present work are contained in the manuscript.

## DATA SHARING STATEMENT

The original contributions presented in the study are included in the article/Supplementary Material. Further inquiries can be directed to the corresponding author.

## AUTHOR CONTRIBUTIONS

EB, KH, XW, and NG designed and performed the systematic literature review and meta-analysis and critically evaluated the manuscript. SK, MN, NKM, MM, and PS performed the systematic literature review and meta-analysis and critically evaluated the manuscript. EB, MTB-J, and NV conceptualized the article and provided oversight and critical evaluation of the manuscript. All authors contributed to the article and approved the submitted version.

## ACKNOWLEDGMENTS

Writing assistance was provided by Erin Bekes, PhD, and Sheri Arndt, PharmD, of ICON (Blue Bell, PA, USA) in accordance with Good Publication Practice (GPP3) guidelines, funded by Moderna, Inc., and under the direction of the authors.

