## Supplemental Material for "Comparative Effectiveness of mRNA-1273 and BNT162b2 COVID-19 Vaccines Among Older Adults: Systematic Literature Review and Meta-Analysis Using the GRADE Framework"

### Appendix 1. Statistical Analysis Methods

#### *Random Effects Meta-Analysis Model*

To account for heterogeneity, exclusively random effects meta-analysis models were used. The random effects model allows the study outcomes to vary among studies; there is a normal distribution of true effect sizes and not one true effect that the fixed effect model assumes. Hence, rather than a common mean as in the fixed effect model, every study has a specific mean.

#### *Inverse Variance Method*

The inverse variance method was applied for the random effects models. This method assigns a weight to each study, which is the inverse of the within-studies variance plus between-studies variance for the random effects model. Thus, larger studies, which have smaller standard errors, are given more weight than smaller ones. This choice of weight minimizes the imprecision of the pooled effect estimate.

#### *Studies with zero cell counts*

Having no events observed in one or both arms of a study included in the meta-analysis (e.g., for death due to COVID-19) causes computational problems using the inverse variance method. Therefore, for studies in which no events were observed in one or both arms, a fixed value of 0.5 was added to the number of events and sample size in both arms (continuity correction).

#### *Conversion approach*

Studies exclusively reporting on vaccine efficacy

A subset of studies exclusively reported vaccine efficacy (VE) instead of number of events and sample size per arm. All outcomes of interest were binary, meaning that, as an input to the meta-analysis, either number of events and sample size, or estimates on the effect size could be used. The meta-analysis results were obtained in terms of risk ratio ($RR_{SC}$), comparing the mRNA-1273 (Spikevax) to the BNT162b2 (Comirnaty) vaccine. Vaccine efficacy was estimated as $1 - RR_{VU}$, $1 - OR_{VU}$ (odds ratio), $1 - HR_{VU}$ (hazard ratio), or $1 - IRR_{VU}$ (incidence rate ratio), comparing the active vaccine arm $V$ (either mRNA-1273 or BNT162b2) to the unvaccinated arm $U$. If VE was estimated from $RR_{VU}$, no conversion was necessary ($RR_{VU} = 1 - VE$). If estimated from another effect size, conversion to $RR_{VU}$ was necessary. Conversion was then performed as described previously (1):

If $OR_{VU} > 1$ with $w \leq p_0 \leq p_1 \leq u$,

$$RR_{VU} = \left\{\frac{OR_{VU}(u + OR_{VU} - OR_{VU}u)}{1 - w + OR_{VU}w}\right\}^{1/2},$$

where $w$ was the lowest outcome probability, $p_0$ was the risk in the comparator group, $p_1$ was the risk in the treatment group, and $u$ was the highest outcome probability.

If $OR_{VU} < 1$ with $w \leq p_0 \leq p_1 \leq u$,

$$RR_{VU} = \left\{\frac{OR_{VU}(w + OR_{VU} - OR_{VU}w)}{1 - u + OR_{VU}u}\right\}^{1/2}.$$

If $HR_{VU} > 1$ with $w \leq p_0 \leq p_1 \leq u$,

$$RR_{VU} = \left\{\frac{1 - (1 - w)^{HR_{VU}}}{1 - (1 - u)^{1/HR_{VU}}} \frac{u}{w}\right\}^{1/2}.$$

If $HR_{VU} < 1$ with $w \leq p_0 \leq p_1 \leq u$,

$$RR_{VU} = \left\{\frac{1 - (1 - u)^{HR_{VU}}}{1 - (1 - w)^{1/HR_{VU}}} \frac{w}{u}\right\}^{1/2}.$$

Conversion was conducted accordingly to estimate the confidence interval (CI) bounds for $OR_{VU}$. However, additionally, the lower bound of the 95% CI was divided by 1.25, whereas the upper bound of the 95% CI was multiplied by 1.25, given that $p_0$ and $p_1$ were between the outcome probabilities $w \geq 0.2$ and $u \leq 0.8$. For values of $p_0$ and $p_1$ outside this interval, the factor 1.25 increased to 1.45 for $HR_{VU}$ (1.67 for $OR_{VU}$).

In several studies, VE was reported as 1-$IRR_{VU}$. The method proposed by VanderWeele (1) did not provide a conversion approach for $IRR_{VU}$ to $RR_{VU}$; therefore, the VE data were not used from these studies. Instead, the rates in the treatment and comparator arms were used directly by transforming rates $\rho_0$ and $\rho_1$ to probabilities $\pi_0$ and $\pi_1$ as

$$\pi_0 = 1 - \exp(-\rho_0),$$

assuming an exponential random variable with rate $\rho_0$, using the cumulative distribution function for a time interval of 1 unit (same approach for $\pi_1$). As a next step, $RR_{VU}$ was then estimated as

$$RR_{VU} = \frac{\pi_1}{\pi_0}.$$

Having estimated $RR_{VU}$ from VE or rates for all studies in which number of events and sample size were not reported directly, $RR_{VU}$ comparing mRNA-1273 vaccination to unvaccinated as well as comparing BNT162b2 vaccination to unvaccinated informed an indirect comparison via common comparator (unvaccinated arm) using the Bucher method (2, 3) to result in $RR_{SC}$, comparing the two active vaccines mRNA-1273 ($S$) and BNT162b2 ($C$) on the log scale, as

$$\ln(RR_{SC}) = \ln(RR_{SU}) - \ln(RR_{CU})$$

$$RR_{SC} = \exp(RR_{SC}) \quad .$$

As for the corresponding standard $SE_{SC}(\ln(RR_{SC}))$, this was estimated from the 95% CIs of $RR_{SU}$ and $RR_{CU}$ as

$$SE_{SU}(\ln(RR_{SU})) = (UCL(\ln(RR_{SU})) - LCL(\ln(RR_{SU})))/3.92;$$

the estimation for $SE_{CU}(\ln(RR_{CU}))$ was conducted accordingly. Finally, the Bucher method was used to estimate

$$SE_{SC}(\ln(RR_{SC})) = \sqrt{(SE_{SU}(\ln(RR_{SU})))^2 + (SE_{CU}(\ln(RR_{CU})))^2}.$$

***Estimation of absolute effects (risk difference) through relative effects***

To estimate absolute effects in terms of risk difference (RD) through relative effects $RR_{SC}$, the probability in the comparator group $p_c$ had to be estimated from the available study data. This corresponded to the probability of having an event (e.g., an infection or death due to COVID-19) in the unvaccinated arm.

Absolute effects (RD) were estimated through relative effects as

$$RD = p_c RR_{SC} - p_c,$$

where $p_c$ represented the probability in the unvaccinated arm.

***Imputation of standard errors***

In some studies VE was reported without any measure of spread, such as 95% CI, standard error (SE), or standard deviation (SD). SE was imputed using the measures of spread of other trials in the evidence base following the steps below (4):

1. The SD of the $\ln(RR_{SC})$ was calculated for every study where information on SE was available (either directly or estimable via RD or 95%CI) as

$$SD(\ln(RR_{SC})) = \frac{SE(\ln(RR_{SC}))}{\frac{1}{N_S} + \frac{1}{N_C}},$$

where $N_S$ and $N_C$ represent the sample size in the mRNA-1273 and BNT162b2 arms, respectively.

2. The average SD of the trials in the network was calculated as

$$\overline{SD(\ln(RR_{SC}))} = \frac{1}{T}\sum_{t=1}^{T} SD\left(\ln(RR_{SC,t})\right),$$

where $T$ represented the overall number of trials, and t represented the respective trial.

3. $\overline{SD(\ln(RR_{SC}))}$ was imputed for the trials that did not report a measure of spread, estimating

$$SE(\ln(RR_{SC})) = \left(\frac{1}{N_S} + \frac{1}{N_C}\right)\overline{SD(\ln(RR_{SC}))}.$$

***Second-order meta-analysis approach***

Different studies reported VE using different definitions, thus, a second-order meta-analysis approach (5) was conducted as a sensitivity analysis.  This method avoids assumptions based on converting odds ratios (ORs), hazard ratios (HRs), or incidence rate ratios (IRRs) to RR as described above, and instead assumes that they were all on the same scale. In a first step, data from studies reporting number of events and sample size were pooled in one meta-analysis, and data from studies reporting VE were pooled in a second meta-analysis. Thereby, no distinction was made regarding estimation of VE from RR, OR, HR, or IRR, and conversion of any kind was avoided. VE was assumed to originate from RR throughout. Of note, uncertainty still exists in these data due to the heterogeneous nature (originating from various effect sizes). RRs estimated from the two meta-analyses in the first step were plotted separately in a forest plot. The pooled RR results informed the second-order meta-analysis, resulting in a final estimate of RR, which was included in the same forest plot. The second step causes an increase in variance, resulting in slightly wider 95% CIs compared with the conversion approach.

Results in terms of absolute effects (RD) could not be estimated using the second-order approach avoiding conversion, since the estimation should be conducted through RR exclusively, and some of the VE data were obtained through alternative effect sizes. Therefore, results are shown in terms of RR only (relative effects).

***Heterogeneity***

Cochran's Q-statistic and $I^2$ are commonly used to quantify heterogeneity in meta-analyses. The Cochran's Q - statistic is calculated as the weighted sum of squared differences between individual study effects and the pooled effects across studies, with the weights (i.e., the inverse variance) being those used in the pooling method. $I^2$ describes the percentage of variation across studies that is attributed to heterogeneity rather than chance.

$I^2 = 100\% \times (Q\text{-}df)/Q$,

where df are the degrees of freedom.

$I^2$ has an intuitive interpretation (6):

- 0% to 40%: might not be important;
- 30% to 60%: may represent moderate heterogeneity*;
- 50% to 90%: may represent substantial heterogeneity*;
- 75% to 100%: considerable heterogeneity*.

*The importance of the observed value of $I^2$ depends on (i) magnitude and direction of effects and (ii) strength of evidence for heterogeneity (e.g., P value of the chi-squared test) (6).

Sensitivity and subgroup analyses were conducted to evaluate whether the sources of clinical heterogeneity could be identified, such as:

- older age (≥65 years or ≥75 years)
- the Delta variant
- severe comorbidities (clinically extremely vulnerable [CEV] groups 1 and 2)
- a higher number of COVID-19 vaccinations (3 doses exclusively)

**Table S1. Published systematic literature reviews cross-checked to identify studies for screening.**

| Author, year | Number of studies included | Study types | Outcomes | Age group(s) | Vaccine |
|---|---|---|---|---|---|
| Li 2022 (7) | 32<br>• mRNA-1273, n=3<br>• BNT162b2, n=3 | • RCTs<br>• Observational studies | • Efficacy<br>• Immunogenicity<br>• Safety | • ≥55 y | Any |
| Graña 2022 (8) | 41<br>• mRNA-1273, n=2<br>• BNT162b2, n=3 | • RCTs | • Efficacy<br>• Immunogenicity<br>• Safety | • Any | Any |
| Sadeghi 2022 (9) | 22<br>• mRNA-1273, n=3<br>• BNT162b2, n=12 | • RCTs<br>• Observational studies | • Efficacy<br>• Immunogenicity<br>• Safety | • <21 y | • Pfizer/<br>• AstraZeneca Moderna/ Cansino |
| Feikin 2022 (10) | 18<br>• mRNA-1273, n=23<br>• BNT162b2, n=38 | • RCTs<br>• Observational studies | • Efficacy<br>• Effectiveness | • Any | Any |
| Asghar 2022 (11) | 29<br>• mRNA-1273, n=3<br>• BNT162b2, n=3 | • RCTs<br>• Non-randomized controlled trials | • Efficacy<br>• Immunogenicity<br>• Safety | • Any | • Pfizer/ AstraZenecaAZ/ Moderna/ Cansino |
| Rahmani 2022 (12) | 54<br>• mRNA-1273, n=8<br>• BNT162b2, n=37 | • Observational studies | • Effectiveness | • Any | Any |
| Korang 2022 (13) | 46<br>• mRNA-1273, n=1 | • RCTs | • Effectiveness<br>• Safety | • Adults | Any |

| Author, year | Number of studies included | Study types | Outcomes | Age group(s) | Vaccine |
|---|---|---|---|---|---|
| | • BNT162b2, n=4 | | | | |
| Mohammed 2022 (14) | 42<br>• mRNA-1273, n=3<br>• BNT162b2, n=16 | • RCTs<br>• Observational studies | • Effectiveness | • Adults | Any |
| Zheng 2022 (15) | 51<br>• mRNA-1273, n=3<br>• BNT162b2, many | • Observational studies | • Effectiveness | • Adults | Any |
| Au 2022 (16) | 63 (44 mRNA) | • Observational studies<br>• RCTs<br>• Non-randomized controlled trials | • Efficacy | • Adults | Any |
| Lv 2022 (17) | 13<br>• BNT162b2, n=12<br>• mRNA-1273, n= 2<br>ChAdOx, n=13 | • Observational studies<br>• RCTs | • Efficacy<br>• Immunogenicity<br>• Safety | • Adult | • BNT162b2<br>• mRNA-1273<br>• ChAdOx1 COVID-19 |
| Iheanacho 2021 (18) | 11<br>• BNT162b2; n=9<br>ChAdOx1, n=5 | • Observational studies | • Efficacy | • Any | • BNT162b2<br>• ChAdOx1 COVID-19 |
| Sharif 2021 (19) | 25<br>• mRNA-1273, n=8<br>• BNT162b2, n=2 | • Observational studies<br>• RCTs<br>• Non-randomized controlled trials | • Efficacy<br>• Safety | • Adults | Any |

| Author, year | Number of studies included | Study types | Outcomes | Age group(s) | Vaccine |
|---|---|---|---|---|---|
| Rotshild 2021 (20) | 8<br>• mRNA-1273, n=1<br>• BNT162b2, n=1 | • RCTs | • Efficacy | • Adults | Any |
| Chen 2021 (21) | 14 (3 mRNA) | • RCTs | • Safety | • Adults | Any |
| Kow 2021 (22) | 19 (BNT162b2) | • Observational studies | • Effectiveness | • Any | BNT162b2 |

RCT, randomized controlled trial.

**Table S2. Database and strategies used for the systematic literature review.**

| **Database** | WHO COVID-19 Global literature on coronavirus disease | | |
|---|---|---|---|
| **URL** | https://search.bvsalud.org/global-literature-on-novel-coronavirus-2019-ncov/ | | |
| **No** | **Query Scope** | **Search Field** | **Query Script** |
| 1 | All COVID-19 | — | Not applicable |
| 2 | Major focus on COVID-19 vaccines | — | (mj: covid-19 vacc*) |
| 3 | Intervention - Moderna | Title, abstract, subject | (mRNA) OR (mRNA-*) OR (Moderna) OR (Spikevax) OR (elasomeran) OR (mRNA-1273) OR (mRNA1273) |
| 4 | Intervention - Pfizer | Title, abstract, subject | (Pfizer) OR (*BioNTech*) OR (Pfizer*) OR (COMIRNATY) OR (Tozinameran) OR (*BNT162b2*) |
| 5 | Intervention - [3] AND [4] Moderna AND Pfizer | Title, abstract, subject | ((mrna) OR (mrna-*) OR (moderna) OR (spikevax) OR (elasomeran) OR (mrna-1273) OR (mrna1273) ) AND ((pfizer) OR (*biontech*) OR (pfizer*) OR (comirnaty) OR (tozinameran) OR (*bnt162b2*)) |
| 6 | Outcomes (Vaccine efficacy, effectiveness) | Title, abstract, subject | (vaccine efficacy) OR (vaccine effectiveness) OR (hospitali*) OR (mortality) OR (death) |
| 7 | Outcomes (Vaccine efficacy, effectiveness) | Title, abstract, subject | (efficacy) OR (effectiveness) OR (hospitali*) OR (mortality) OR (death) |
| 8 | Outcomes (Vaccine efficacy, effectiveness) | Title, abstract, subject | (infection) |

| | | | |
|---|---|---|---|
| 9 | [6] OR [7] OR [8] without filter | — | ((vaccine efficacy) OR (vaccine effectiveness) OR (hospitali*) OR (mortality) OR (death)) OR ((efficacy) OR (effectiveness) OR (hospitali*) OR (mortality) OR (death)) OR ((infection)) |
| 10 | [2] AND [5] AND [9] without filter | — | ((mj: covid-19 vacc*)) AND (((mrna) OR (mrna-*) OR (moderna) OR (spikevax) OR (elasomeran) OR (mrna-1273) OR (mrna1273) ) AND ((pfizer) OR (*biontech*) OR (pfizer*) OR (comirnaty) OR (tozinameran) OR (*bnt162b2*))) AND (((vaccine efficacy) OR (vaccine effectiveness) OR (hospitali*) OR (mortality) OR (death)) OR ((efficacy) OR (effectiveness) OR (hospitali*) OR (mortality) OR (death)) OR ((infection))) |
| 11 | [10] with MEDLINE | — | ((mj: covid-19 vacc*)) AND (((mrna) OR (mrna-*) OR (moderna) OR (spikevax) OR (elasomeran) OR (mrna-1273) OR (mrna1273) ) AND ((pfizer) OR (*biontech*) OR (pfizer*) OR (comirnaty) OR (tozinameran) OR (*bnt162b2*))) AND (((vaccine efficacy) OR (vaccine effectiveness) OR (hospitali*) OR (mortality) OR (death)) OR ((efficacy) OR (effectiveness) OR (hospitali*) OR (mortality) OR (death)) OR ((infection))) AND db:("MEDLINE") |
| 12 | [11] with following filters on<br>- Observational study | — | ((mj: covid-19 vacc*)) AND (((mrna) OR (mrna-*) OR (moderna) OR (spikevax) OR (elasomeran) OR (mrna-1273) OR (mrna1273) ) AND ((pfizer) OR (*biontech*) OR (pfizer*) OR (comirnaty) OR (tozinameran) OR (*bnt162b2*))) AND |

| | | | |
|---|---|---|---|
| | - Cohort study<br>- Randomized controlled trials<br>- Systematic review/Meta Analysis | | (((vaccine efficacy) OR (vaccine effectiveness) OR (hospitali*) OR (mortality) OR (death)) OR ((efficacy) OR (effectiveness) OR (hospitali*) OR (mortality) OR (death)) OR ((infection))) AND db:("MEDLINE") AND type_of_study:("observational_studies" OR "cohort_studies" OR "rct" OR "systematic_reviews") |
| 13 | [12] with Language: English | — | ((mj: covid-19 vacc*)) AND (((mrna) OR (mrna-*) OR (moderna) OR (spikevax) OR (elasomeran) OR (mrna-1273) OR (mrna1273) ) AND ((pfizer) OR (biontech) OR (pfizer*) OR (comirnaty) OR (tozinameran) OR (bnt162b2))) AND (((vaccine efficacy) OR (vaccine effectiveness) OR (hospitali*) OR (mortality) OR (death)) OR ((efficacy) OR (effectiveness) OR (hospitali*) OR (mortality) OR (death)) OR ((infection))) AND db:("MEDLINE") AND type_of_study:("observational_studies" OR "cohort_studies" OR "rct" OR "review" OR "systematic_reviews") AND la:("en") |
| 14 | [13] with Year: 2022 onwards | — | ((mj: covid-19 vacc*)) AND (((mrna) OR (mrna-*) OR (moderna) OR (spikevax) OR (elasomeran) OR (mrna-1273) OR (mrna1273) ) AND ((pfizer) OR (biontech) OR (pfizer*) OR (comirnaty) OR (tozinameran) OR (bnt162b2))) AND (((vaccine efficacy) OR (vaccine effectiveness) OR (hospitali*) OR (mortality) OR (death)) OR ((efficacy) OR (effectiveness) OR (hospitali*) OR (mortality) OR (death)) OR ((infection))) AND db:("MEDLINE") |

| | | | |
|---|---|---|---|
| | | | AND type_of_study:("observational_studies" OR "cohort_studies" OR "rct" OR "review" OR "systematic_reviews") AND la:("en") AND year_cluster:("2022" OR "2023") |
| **URL** | https://search.bvsalud.org/global-literature-on-novel-coronavirus-2019-ncov/ | | |
| **No** | **Query Scope** | **Search Field** | **Query Script** |
| 1 | All COVID-19 | — | Not applicable |
| 2 | COVID-19 vaccines | — | ("covid19 vaccine" ~2 OR "covid-19 vaccine"~2 OR "covid-19 vaccines"~2 OR "covid19 vaccines" ~2 OR "covid-19 vaccination"~2 OR "Covid19 vaccination"~2) |
| 3 | Intervention - Moderna | Title, abstract, subject | (mRNA) OR (mRNA-*) OR (Moderna) OR (Spikevax) OR (elasomeran) OR (mRNA-1273) OR (mRNA1273) |
| 4 | Intervention - Pfizer | Title, abstract, subject | (Pfizer) OR (*BioNTech*) OR (Pfizer*) OR (COMIRNATY) OR (Tozinameran) OR (*BNT162b2*) |
| 5 | Intervention - [3] AND [4] Moderna AND Pfizer | Title, abstract, subject | ((mrna) OR (mrna-*) OR (moderna) OR (spikevax) OR (elasomeran) OR (mrna-1273) OR (mrna1273) ) AND ((pfizer) OR (*biontech*) OR (pfizer*) OR (comirnaty) OR (tozinameran) OR (*bnt162b2*)) |
| 6 | Outcomes (Vaccine efficacy, effectiveness) | Title, abstract, subject | (vaccine efficacy) OR (vaccine effectiveness) OR (hospitali*) OR (mortality) OR (death) |
| 7 | Outcomes (Vaccine efficacy, effectiveness) | Title, abstract, subject | (efficacy) OR (effectiveness) OR (hospitali*) OR (mortality) OR (death) |

| | | | |
|---|---|---|---|
| 8 | Outcomes (Vaccine efficacy, effectiveness) | Title, abstract, subject | (infection) |
| 9 | [6] OR [7] OR [8] without filter | — | ((vaccine efficacy) OR (vaccine effectiveness) OR (hospitali*) OR (mortality) OR (death)) OR ((efficacy) OR (effectiveness) OR (hospitali*) OR (mortality) OR (death)) OR ((infection)) |
| 10 | [2] AND [5] AND [9] without filter | — | (("covid19 vaccine" ~2 OR "covid-19 vaccine"~2 OR "covid-19 vaccines"~2 OR "covid19 vaccines" ~2 OR "covid-19 vaccination"~2 OR "Covid19 vaccination"~2)) AND (((mrna) OR (mrna-*) OR (moderna) OR (spikevax) OR (elasomeran) OR (mrna-1273) OR (mrna1273) ) AND ((pfizer) OR (*biontech*) OR (pfizer*) OR (comirnaty) OR (tozinameran) OR (*bnt162b2*))) AND (((vaccine efficacy) OR (vaccine effectiveness) OR (hospitali*) OR (mortality) OR (death)) OR ((efficacy) OR (effectiveness) OR (hospitali*) OR (mortality) OR (death)) OR ((infection))) |
| 11 | [10] with other databases (except MEDLINE) | — | (("covid19 vaccine" ~2 OR "covid-19 vaccine"~2 OR "covid-19 vaccines"~2 OR "covid19 vaccines" ~2 OR "covid-19 vaccination"~2 OR "Covid19 vaccination"~2)) AND (((mrna) OR (mrna-*) OR (moderna) OR (spikevax) OR (elasomeran) OR (mrna-1273) OR (mrna1273) ) AND ((pfizer) OR (*biontech*) OR (pfizer*) OR (comirnaty) OR |

| | | | |
|---|---|---|---|
| | | | (tozinameran) OR (*bnt162b2*))) AND (((vaccine efficacy) OR (vaccine effectiveness) OR (hospitali*) OR (mortality) OR (death)) OR ((efficacy) OR (effectiveness) OR (hospitali*) OR (mortality) OR (death)) OR ((infection))) AND db:("PREPRINT-MEDRXIV" OR "PREPRINT-RESEARCHSQUARE" OR "PREPRINT-SSRN" OR "PREPRINT-BIORXIV" OR "EMBASE" OR "PREPRINT-AUTHOREA PREPRINTS" OR "PREPRINT-PREPRINTS.ORG" OR "PREPRINT-ARXIV" OR "ICTRP") |
| 12 | [11] with Language: English | — | (("covid19 vaccine" ~2 OR "covid-19 vaccine"~2 OR "covid-19 vaccines"~2 OR "covid19 vaccines" ~2 OR "covid-19 vaccination"~2 OR "Covid19 vaccination"~2)) AND (((mrna) OR (mrna-*) OR (moderna) OR (spikevax) OR (elasomeran) OR (mrna-1273) OR (mrna1273) ) AND ((pfizer) OR (*biontech*) OR (pfizer*) OR (comirnaty) OR (tozinameran) OR (*bnt162b2*))) AND (((vaccine efficacy) OR (vaccine effectiveness) OR (hospitali*) OR (mortality) OR (death)) OR ((efficacy) OR (effectiveness) OR (hospitali*) OR (mortality) OR (death)) OR ((infection))) AND db:("PREPRINT-MEDRXIV" OR "PREPRINT-RESEARCHSQUARE" OR "PREPRINT-BIORXIV" OR "EMBASE" OR "PREPRINT-PREPRINTS.ORG" OR "PREPRINT-AUTHOREA PREPRINTS" OR |

| | | | |
|---|---|---|---|
| | | | "PREPRINT-SSRN" OR "PREPRINT-ARXIV" OR "ICTRP") AND la:("en") |
| 13 | [12] with Year: 2022 onwards | — | (("covid19 vaccine" ~2 OR "covid-19 vaccine"~2 OR "covid-19 vaccines"~2 OR "covid19 vaccines" ~2 OR "covid-19 vaccination"~2 OR "Covid19 vaccination"~2)) AND (((mrna) OR (mrna-*) OR (moderna) OR (spikevax) OR (elasomeran) OR (mrna-1273) OR (mrna1273) ) AND ((pfizer) OR (*biontech*) OR (pfizer*) OR (comirnaty) OR (tozinameran) OR (*bnt162b2*))) AND (((vaccine efficacy) OR (vaccine effectiveness) OR (hospitali*) OR (mortality) OR (death)) OR ((efficacy) OR (effectiveness) OR (hospitali*) OR (mortality) OR (death)) OR ((infection))) AND db:("PREPRINT-MEDRXIV" OR "PREPRINT-RESEARCHSQUARE" OR "PREPRINT-SSRN" OR "PREPRINT-BIORXIV" OR "EMBASE" OR "PREPRINT-AUTHOREA PREPRINTS" OR "PREPRINT-PREPRINTS.ORG" OR "PREPRINT-ARXIV" OR "ICTRP") AND la:("en") AND year_cluster:("2022" OR "2023") |

**Table S3. Summary of population, exposure, comparison, and outcomes used in systematic literature review.**

| | | |
|---|---|---|
| **Research question** | Is mRNA-1273 more effective than BNT162b2 at preventing SARS-CoV-2 infections and COVID-19-related hospitalizations and deaths in older adults aged ≥50 years? | |
| | **Include** | **Exclude** |
| **Population** | Healthy adults, aged ≥50 years (healthy as defined by the paper; some individuals with comorbidities may be present, including some individuals with conditions within CEV group 1 or 2 (23) | • Pregnant women, current/former smokers, physically inactive<br>• Studies on only immunocompromised individuals within CEV groups 1 and 2 (23) |
| **Exposure** | mRNA-1273 | No data specific to mRNA-1273- or BNT162b2 (i.e., data on mixed mRNA-1273, BNT162b2, or other vaccines) |
| **Comparison** | BNT162b2 | |
| **Outcomes** | • Vaccine efficacy/effectiveness against COVID-19 infection<br>• Vaccine efficacy against symptomatic COVID-19 infection<br>• Vaccine efficacy against severe COVID-19 infection<br>• Vaccine efficacy against hospitalization<br>• Vaccine efficacy against death<br>• SARS-CoV2 infection (test positive with/without symptoms)<br>• Symptomatic laboratory-confirmed COVID-19 infection<br>• Severe COVID-19 infection | Studies with only safety and immunogenicity outcomes |

| | | |
|---|---|---|
| | • Breakthrough infection<br>• COVID-19 re-infection<br>• Hospitalization due to COVID-19 (ICU, ER, or ventilation, etc.)<br>• Death due to COVID-19 | |
| **Study design** | • Clinical trials<br>• Observational studies<br>• Any kind of real-world evidence | • Study protocol (no results)<br>• Economic models<br>• Single-arm studies |
| **Other limits** | • Any publication type (including letters, commentary, abstract, full text, poster)<br>• Publication in English | |

CEV, clinically extremely vulnerable; COVID-19, coronavirus disease 2019; ER, emergency room; ICU, intensive care unit; RCT, randomized controlled trial.

**Table S4. Results from risk of bias assessment based on the Newcastle-Ottawa Scale.[a]**

| Cohort studies | | | | | | | | | |
|---|---|---|---|---|---|---|---|---|---|
| **Author, year** | **Total score** | **Representativeness of exposed cohort[b]** | **Selection of nonexposed cohort[c]** | **Ascertain-ment of exposure[d]** | **Outcome not present at baseline[e]** | **Comparability of cohorts[f]** | **Assessment of outcome[g]** | **Sufficient follow-up duration[h]** | **Adequate follow-up[i]** |
| Bello-Chavolla 2023 (24) | 8 stars | 1 star | 1 star | 1 star | 1 star | 1 star | 1 star | 1 star | 1 star |
| Braeye 2023 (25) | 7 stars | 1 star | 1 star | 1 star | 1 star | 1 star | 1 star | 1 star | 0 star |
| Breznik 2023 (26) | 6 stars | 1 star | 1 star | 1 star | 1 star | 1 star | 1 star | 0 star | 0 star |
| Butt 2022 (27) | 7 stars | 1 star | 1 star | 1 star | 1 star | 1 star | 1 star | 1 star | 0 star |
| Chemaitelly 2022 (28) | 8 stars | 1 star | 1 star | 1 star | 1 star | 1 star | 1 star | 1 star | 1 star |

| | | | | | | | | | |
|---|---|---|---|---|---|---|---|---|---|
| Hatfield 2022 (29) | 8 stars | 1 star | 1 star | 1 star | 1 star | 1 star | 1 star | 1 star | 1 star |
| Kelly 2022 (30) | 8 stars | 1 star | 1 star | 1 star | 1 star | 1 star | 1 star | 1 star | 1 star |
| Lin 2022 (31) | 8 stars | 1 star | 1 star | 1 star | 1 star | 1 star | 1 star | 1 star | 1 star |
| Lytras 2022 (32) | 6 stars | 1 star | 0 star | 1 star | 1 star | 0 star | 1 star | 1 star | 1 star |
| Martinez-Baz 2021 (33) | 7 stars | 1 star | 1 star | 1 star | 1 star | 0 star | 1 star | 1 star | 1 star |
| Moline 2021 (34) | 6 stars | 1 star | 1 star | 1 star | 1 star | 1 star | 1 star | 0 star | 0 star |
| Nguyen 2023 (35) | 8 stars | 1 star | 1 star | 1 star | 1 star | 1 star | 1 star | 1 star | 1 star |
| Puranik 2022 (36) | 8 stars | 1 star | 1 star | 1 star | 1 star | 1 star | 1 star | 1 star | 1 star |

| | | | | | | | | | |
|---|---|---|---|---|---|---|---|---|---|
| Robles-Fontan 2022 (37) | 8 stars | 1 star | 1 star | 1 star | 1 star | 1 star | 1 star | 1 star | 1 star |
| Rosenberg 2022 (38) | 8 stars | 1 star | 1 star | 1 star | 1 star | 1 star | 1 star | 1 star | 1 star |
| Starrfelt 2022 (39) | 8 stars | 1 star | 1 star | 1 star | 1 star | 1 star | 1 star | 1 star | 1 star |
| Voko 2022 (40) | 8 stars | 1 star | 1 star | 1 star | 1 star | 1 star | 1 star | 1 star | 1 star |
| Voko 2022a (41) | 8 stars | 1 star | 1 star | 1 star | 1 star | 1 star | 1 star | 1 star | 1 star |
| **Case-control studies** | | | | | | | | | |
| **Author, year** | **Total score** | **Adequate case definition**[j] | **Representative-ness of the cases**[k] | **Selection of controls**[l] | **Definition of controls**[m] | **Comparability**[n] | **Ascertain-ment of exposure**[o] | **Same method of ascertain-ment for cases and controls**[p] | **Non-response rate**[q] |

| | | | | | | | | | |
|---|---|---|---|---|---|---|---|---|---|
| Chico-Sanchez 2022 (42) | 5 stars | 1 star | 0 star | 1 star | 0 star | 1 star | 1 star | 1 star | 0 stars |
| Grewal 2022 (43) | 7 stars | 1 star | 1 star | 1 star | 0 star | 1 star | 1 star | 1 star | 1 star |
| Kissling 2022 (44) | 7 stars | 1 star | 1 star | 1 star | 0 star | 1 star | 1 star | 1 star | 1 star |
| Thompson 2021 (45) | 8 stars | 1 star | 1 star | 1 star | 1 star | 1 star | 1 star | 1 star | 1 star |
| Van Ewijk 2022 (46) | 7 stars | 1 star | 1 star | 0 star | 1 star | 1 star | 1 star | 1 star | 1 star |

[a]Risk of bias was not estimable for one study.

[b]1 star was given if the study population was truly or somewhat representative of a community or population.

[c]1 star was given if the nonexposed cohort was drawn from the same community as the exposed cohort. If the nonexposed cohort was drawn from a different source, no star was awarded.

[d]1 star was given if a secure record (e.g., medical record) or a structured interview was used to ascertain exposure.

[e]1 star was given if the outcomes were assessed at the beginning of the study.

[f]1 star was given if the study was adjusted for age, sex, and marital status or other important factors. If the cohorts were not comparable on the basis of the design or the analysis did not control for confounders, no star was awarded.

[g]1 star was given if the outcome was assessed by independent blind assessment or record linkage.

[h]1 star was given if the duration of follow-up was sufficient.

[i]1 star was given if there was complete follow-up or the lost to follow-up rate was ≤20%. No star was awarded if the follow-up rate was <80% or if the follow-up rate was not reported.

[j]1 star was given if the case definition was adequate and independently validated. No star was given if the case definition was based on record linkage, or self-reported, or not described.

[k]1 star was given if the cases were consecutive or a representative series of cases. No star was given if there was a potential for selection biases or the representativeness of cases was not stated.

[l]1 star was given if community controls were used. No star was awarded if hospital controls were used or controls were not described.

[m]1 star was given if the control had no history of disease. No star was given if no description was provided.

[n]1 star was given if the study controlled for the most important factor; 1 star was also given if the study controlled for any additional factors (up to 2 stars).

[o]1 star was given if exposure was ascertained from a secure record or structured interview with blinding to the case/control status. No stars were given if this was ascertained by non-blinded interviews or written self-report or if no description was provided.

[p]1 star was awarded if the method was the same. If a different method was used to ascertain cases and controls, no star was given.

[q]1 star was given if the nonresponse rate was the same for both cases and controls. No star was given if the nonresponse rate was different or not described.

**Figure S1. Summary of pairwise meta-analysis conversion approach.**

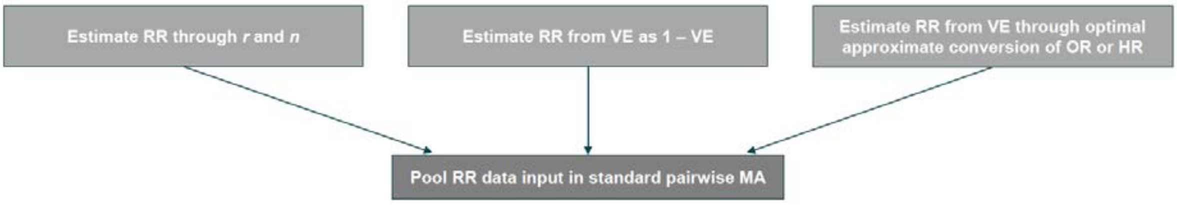


HR, hazard ratio; *n*, sample size per arm; MA, meta-analysis; OR, odds ratio; *r*, number of events per arm; RR, risk ratio; MA, meta-analysis; VE, vaccine effectiveness

**Figure S2. Meta-analysis results on clinical effectiveness outcomes of the mRNA-1273 versus BNT162b2 COVID-19 vaccines using second order methodological approach.**

**(A)** ***SARS-CoV-2 Infection***

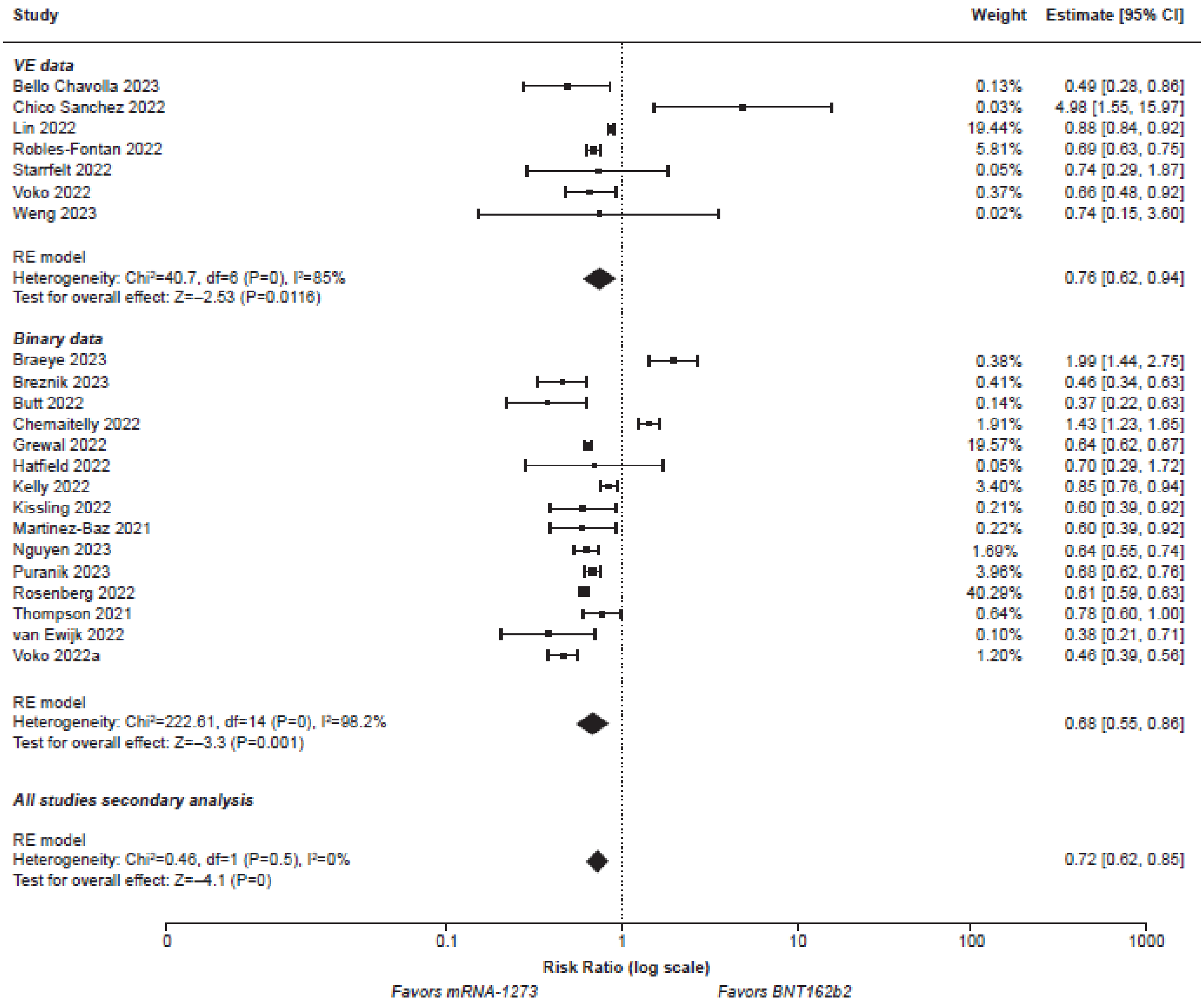

### (B) *Severe SARS-CoV-2 Infection*

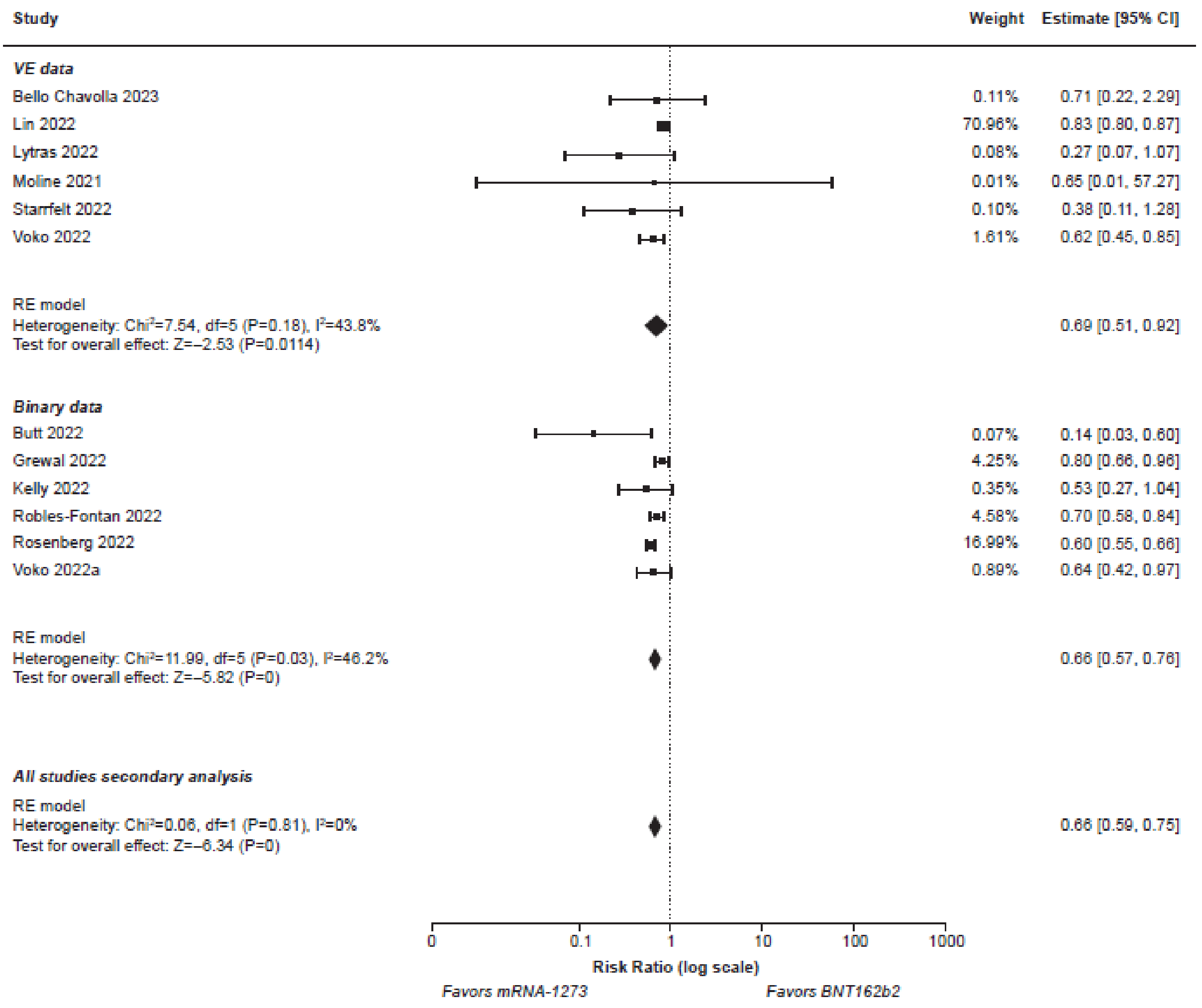

### (C) *Hospitalization*

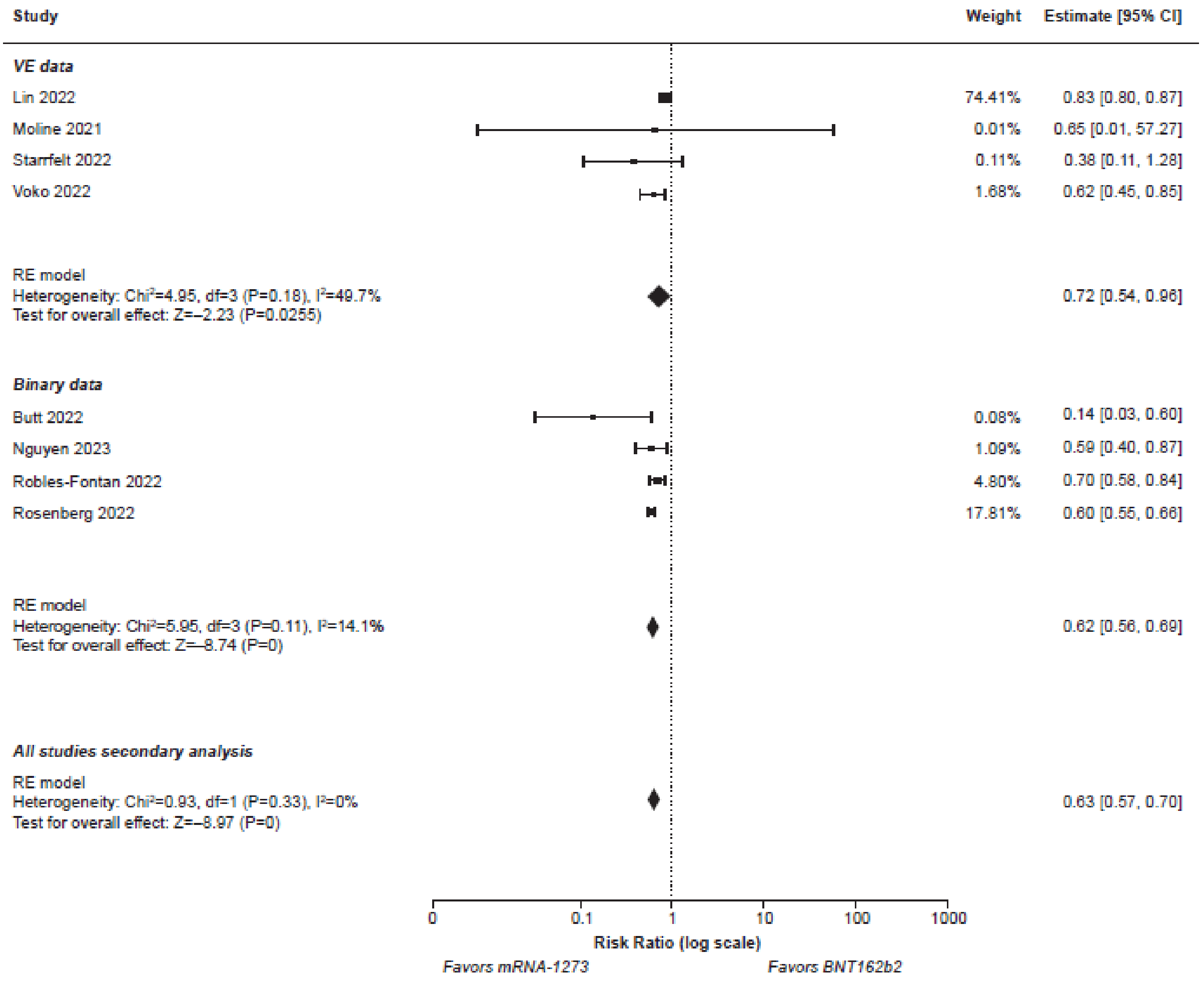

**(D)** ***Death***

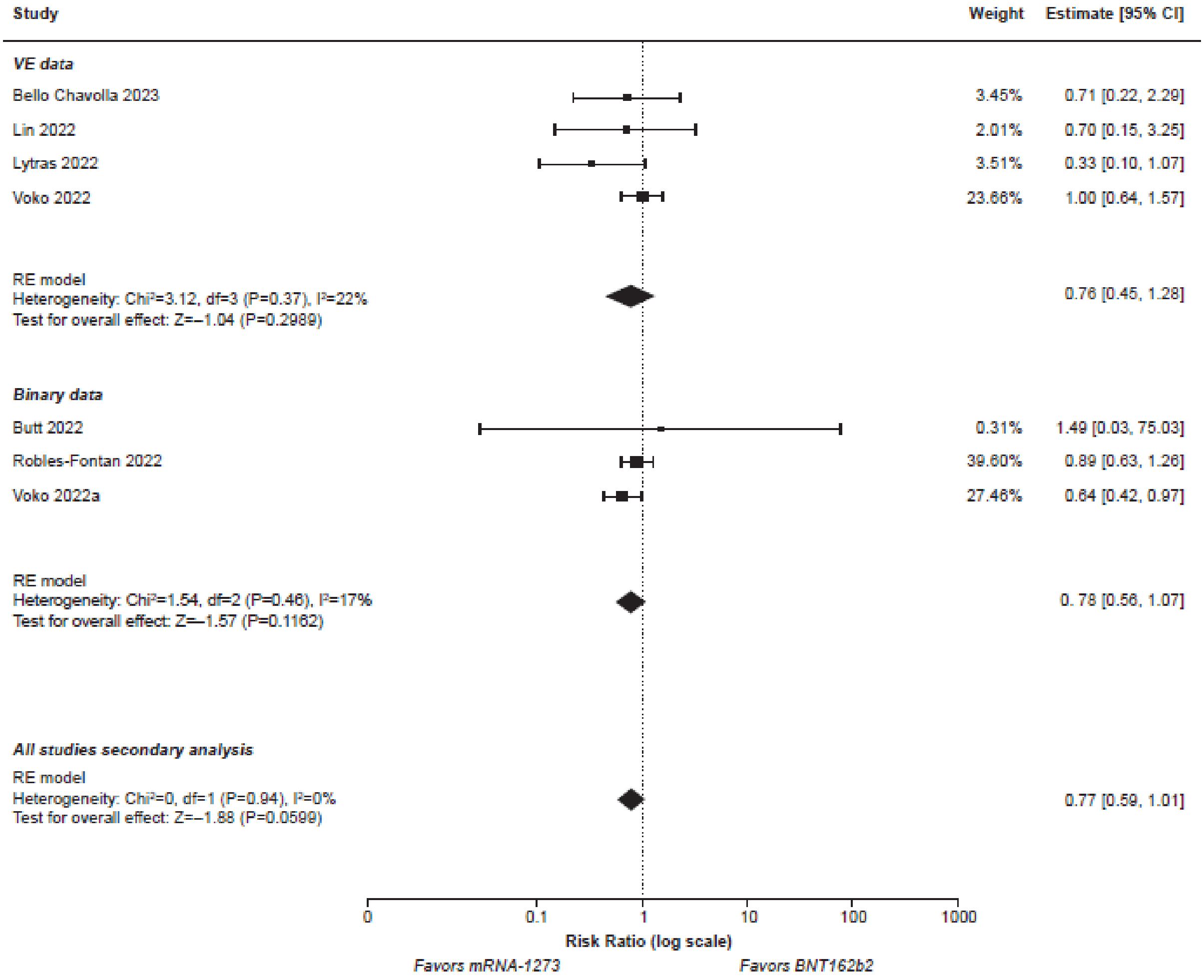

Study
Weight
Estimate [95% CI]
VE data
Bello Chavolla 2023
3.45%
0.71 [0.22, 2.29]
Lin 2022
2.01%
0.70 [0.15, 3.25]
Lytras 2022
3.51%
0.33 [0.10, 1.07]
Voko 2022
23.66%
1.00 [0.64, 1.57]
RE model
Heterogeneity: Chi²=3.12, df=3 (P=0.37), I²=22%
Test for overall effect: Z=–1.04 (P=0.2989)
0.76 [0.45, 1.28]
Binary data
Butt 2022
0.31%
1.49 [0.03, 75.03]
Robles-Fontan 2022
39.60%
0.89 [0.63, 1.26]
Voko 2022a
27.46%
0.64 [0.42, 0.97]
RE model
Heterogeneity: Chi²=1.54, df=2 (P=0.46), I²=17%
Test for overall effect: Z=–1.57 (P=0.1162)
0. 78 [0.56, 1.07]
All studies secondary analysis
RE model
Heterogeneity: Chi²=0, df=1 (P=0.94), I²=0%
Test for overall effect: Z=–1.88 (P=0.0599)
0.77 [0.59, 1.01]
0
0.1
1
10
100
1000
Risk Ratio (log scale)
Favors mRNA-1273
Favors BNT162b2

**Figure S3. Meta-analysis results on clinical effectiveness outcomes of the mRNA-1273 versus BNT162b2 COVID-19 vaccines in the subgroup of older adults aged ≥65 years.**

**(A)** ***SARS-CoV-2 Infection***

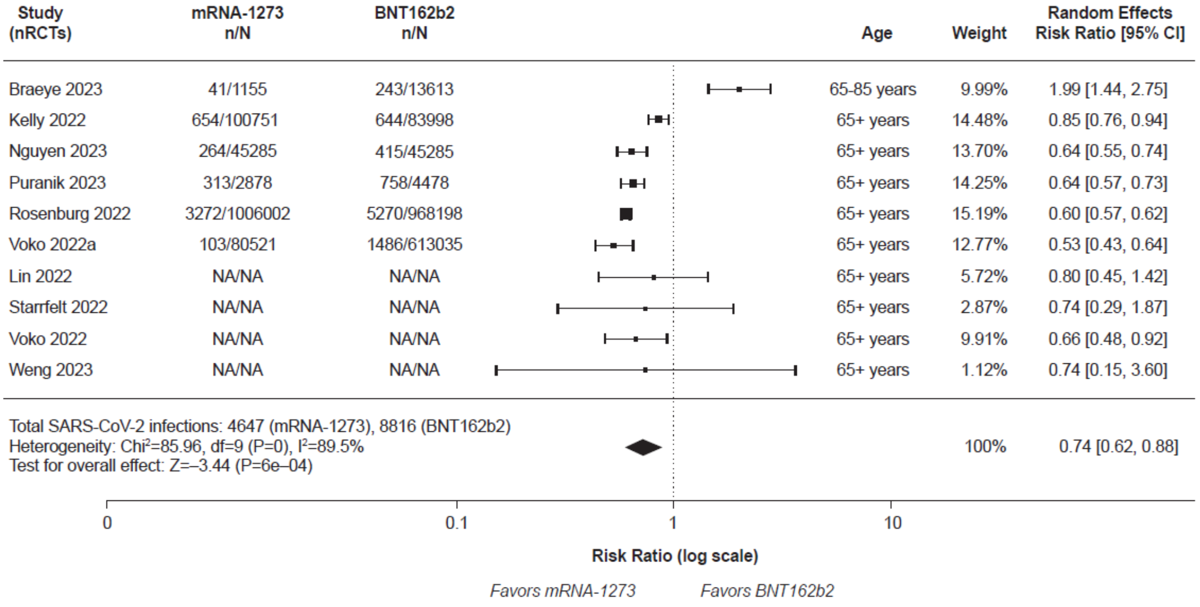

**(B)** ***Laboratory-Confirmed Symptomatic SARS-CoV-2 Infection***

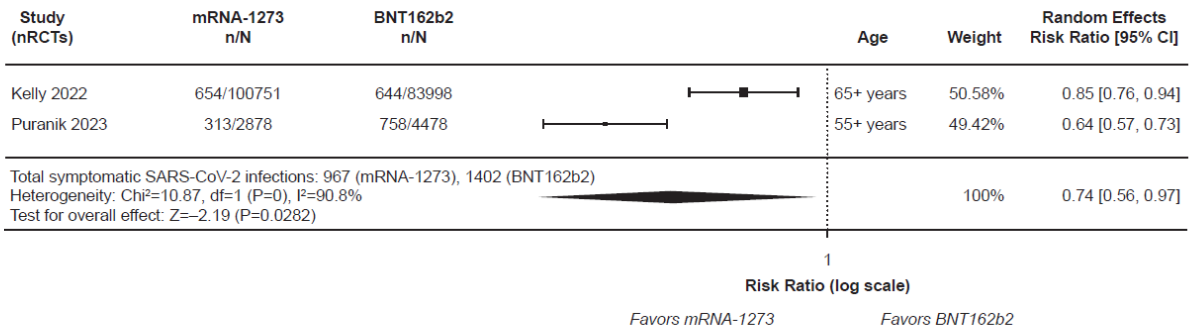


**(C)** ***Severe SARS-CoV-2 Infection***

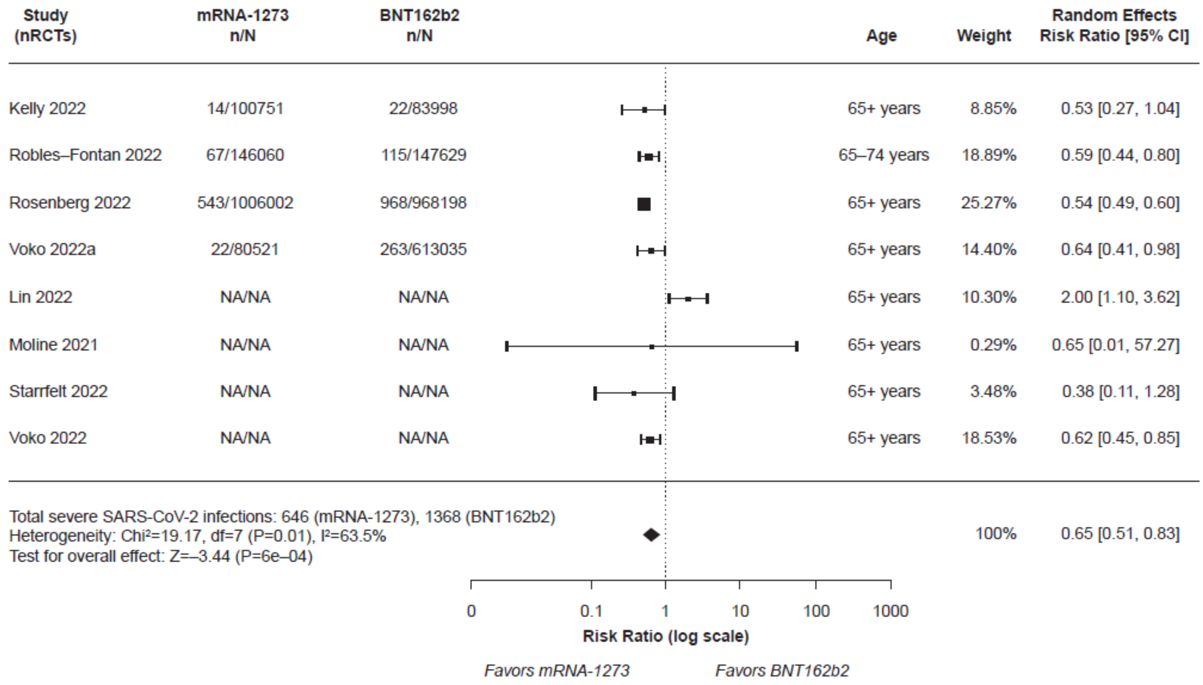

**(D)** ***Hospitalization***

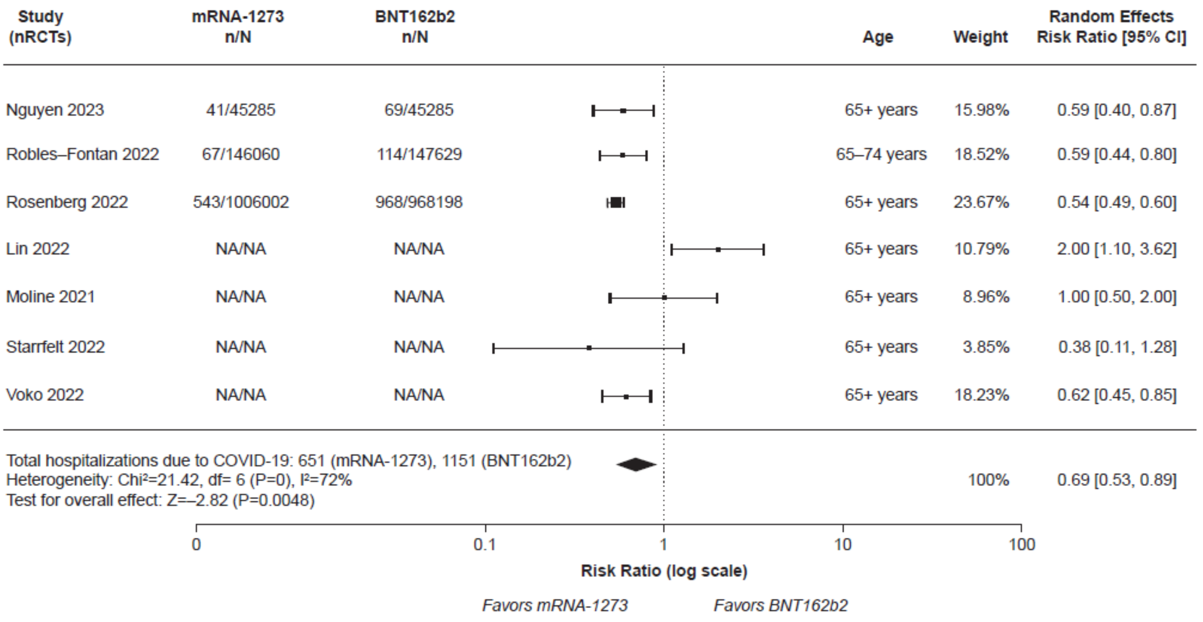


**(E)** ***Death***

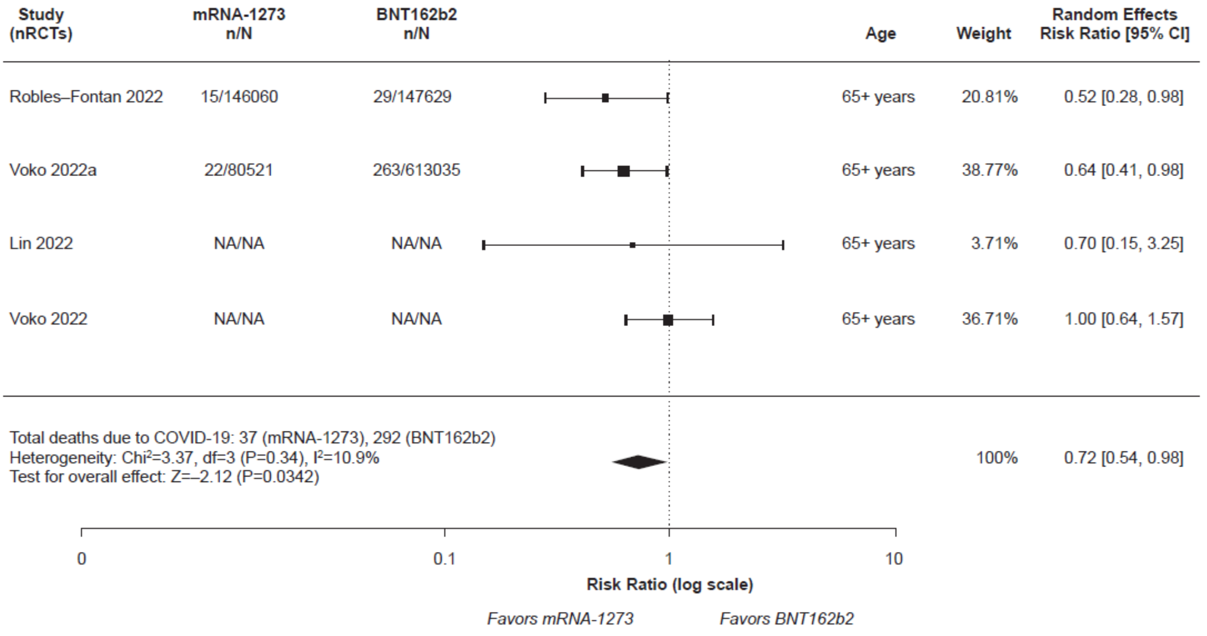

**Figure S4. Meta-analysis results on clinical effectiveness outcomes of the mRNA-1273 versus BNT162b2 COVID-19 vaccines in the subgroup of older adults aged ≥50 years who received exclusively 3 doses.**

**(A) *SARS-CoV-2 Infection***

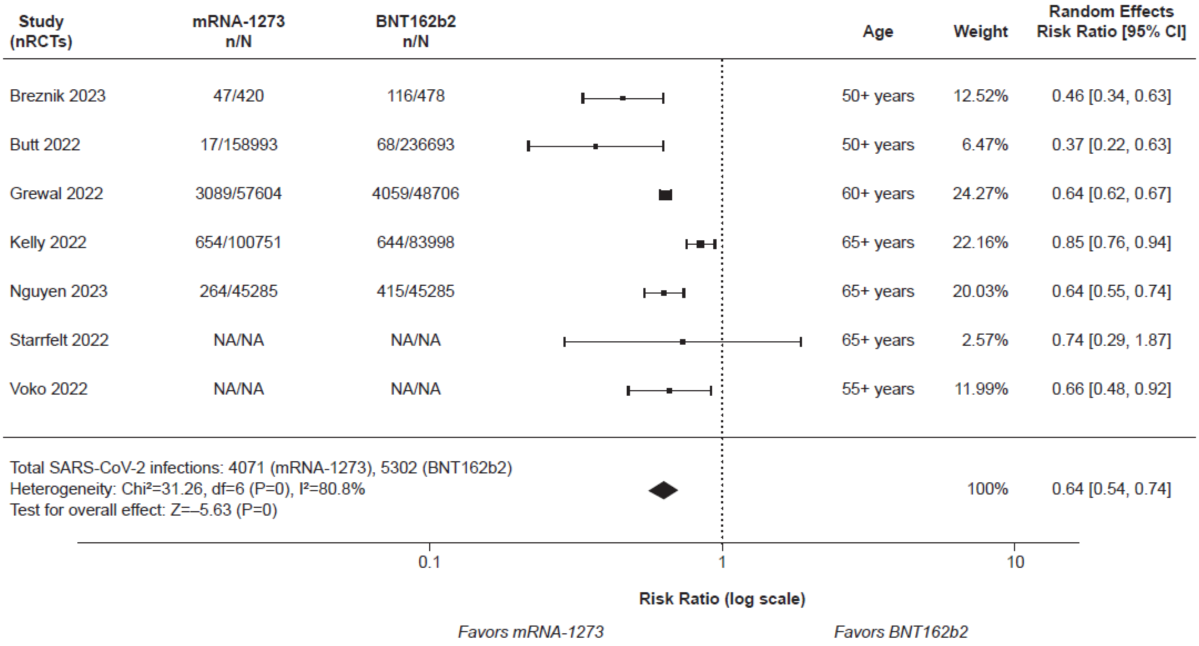

### (B) *Laboratory-Confirmed Symptomatic SARS-CoV-2 Infection*

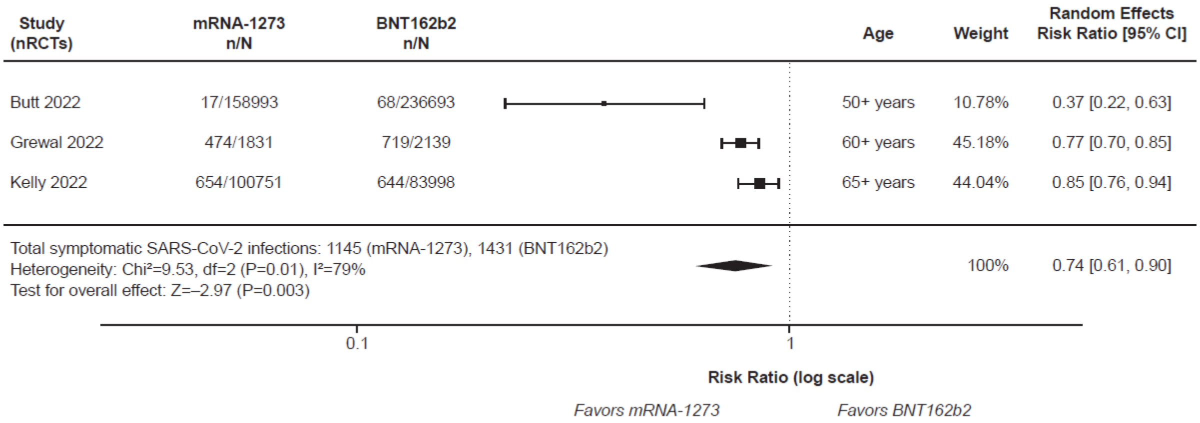


### (C) *Severe SARS-CoV-2 Infection*

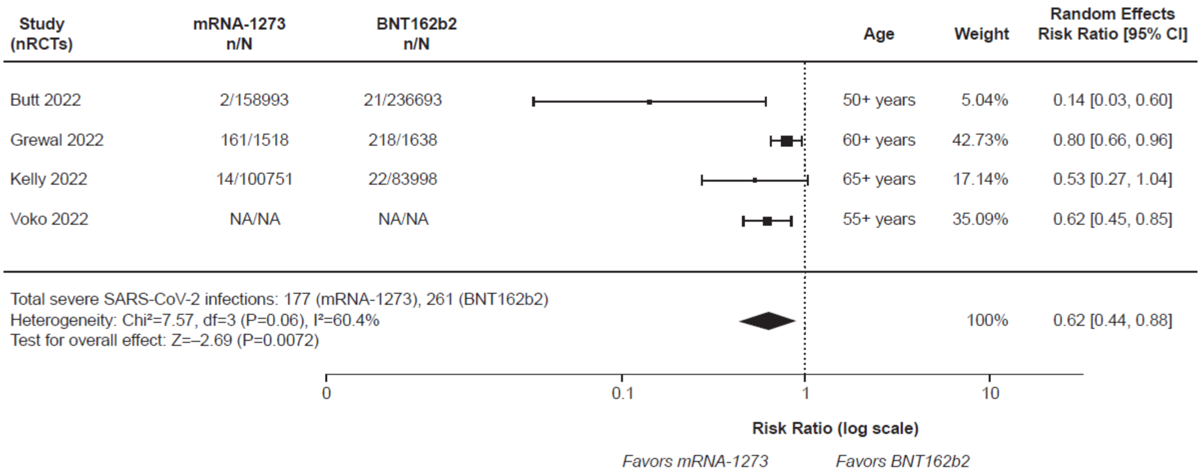

**(D) *Hospitalization***

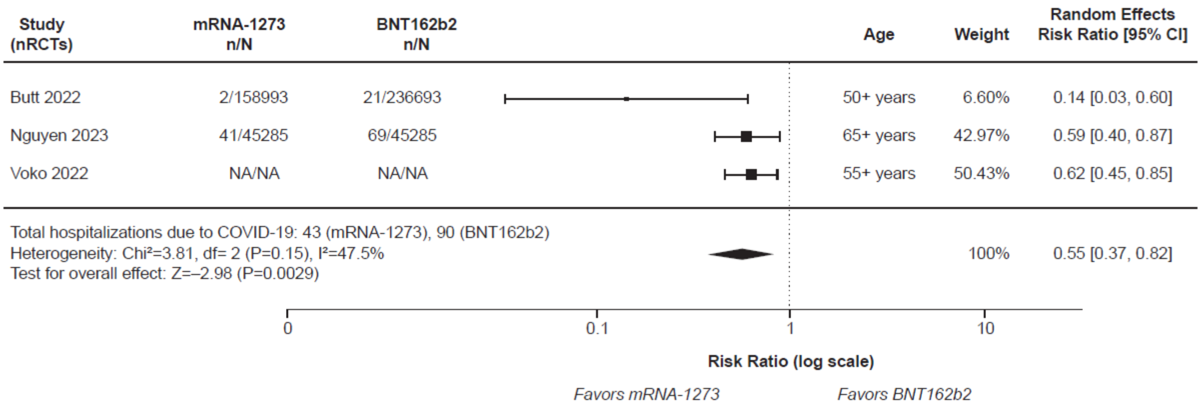


**(E) *Death***

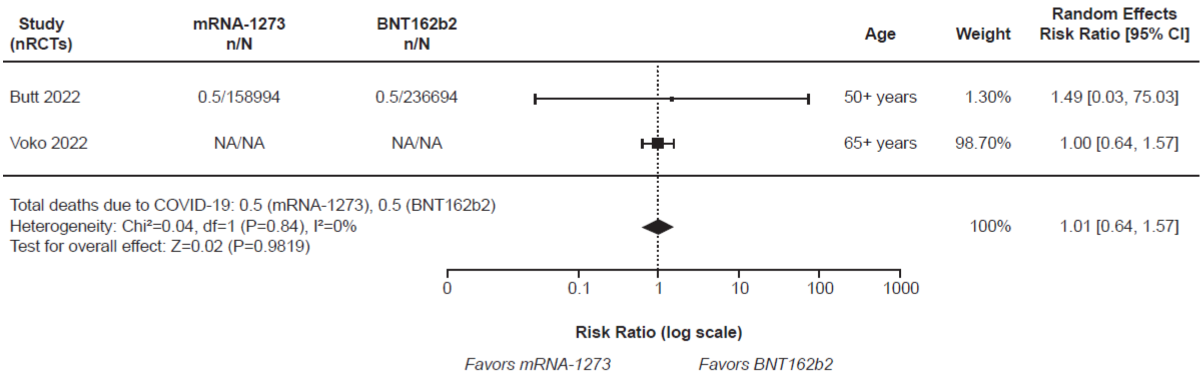

**Figure S5. Meta-analysis results on clinical effectiveness outcomes of the mRNA-1273 versus BNT162b2 COVID-19 vaccines in the subgroup of older adults aged ≥75 years**

**(A)** ***SARS-CoV-2 Infection***

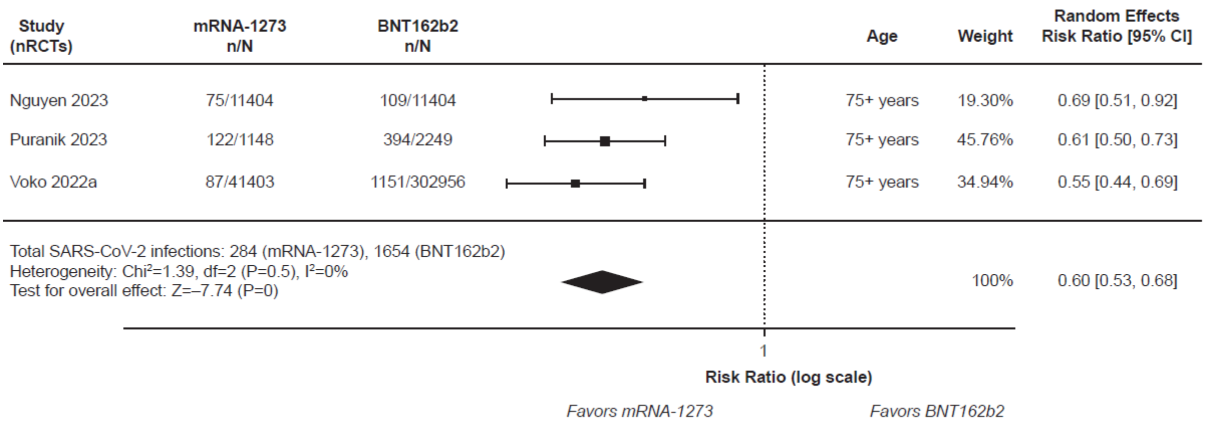

**(B)** ***Laboratory-Confirmed Symptomatic SARS-CoV-2 Infection***

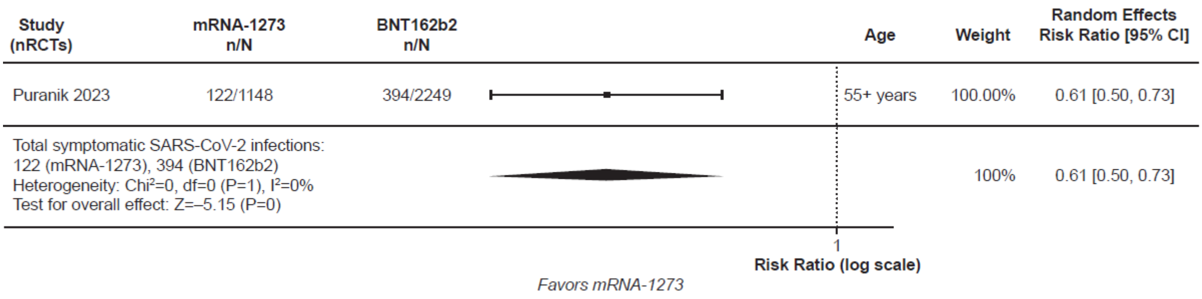


**(C)** ***Severe SARS-CoV-2 Infection***

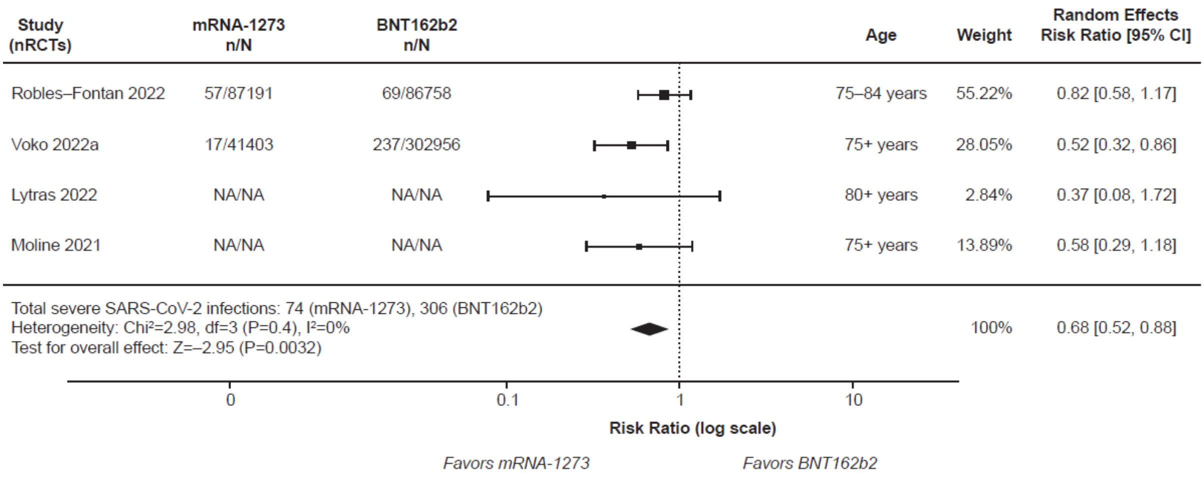

**(D)** ***Hospitalization***

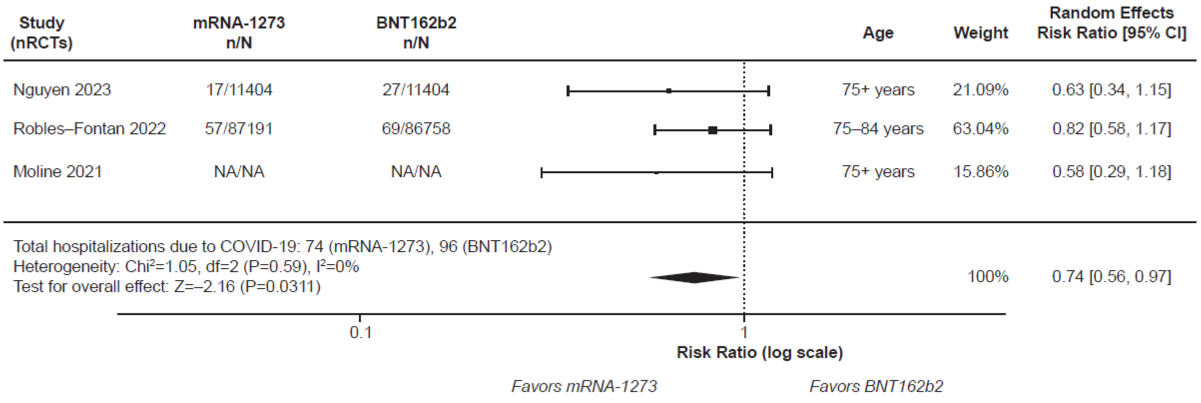


**(E)** ***Death***

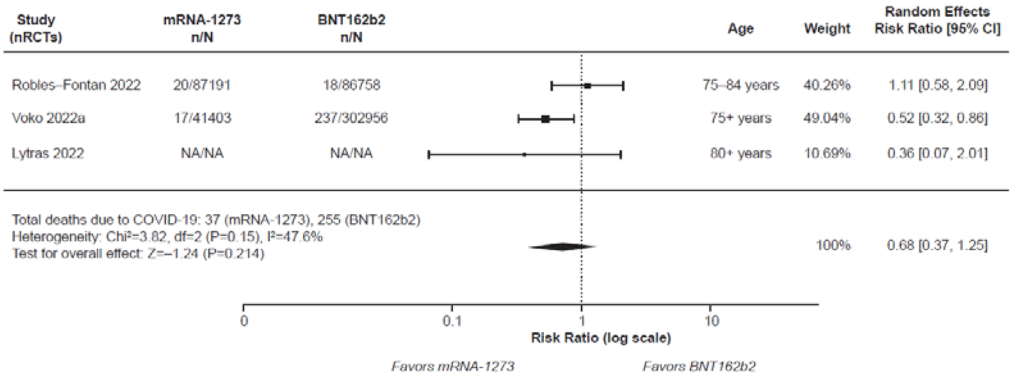

**Figure S6. Meta-analysis results on clinical effectiveness outcomes of the mRNA-1273 versus BNT162b2 COVID-19 vaccines in the subgroup of older adults aged ≥50 years, excluding patients in clinically extremely vulnerable groups 1 and 2. CEV, clinically extremely vulnerable.**

(A) ***SARS-CoV-2 Infection***

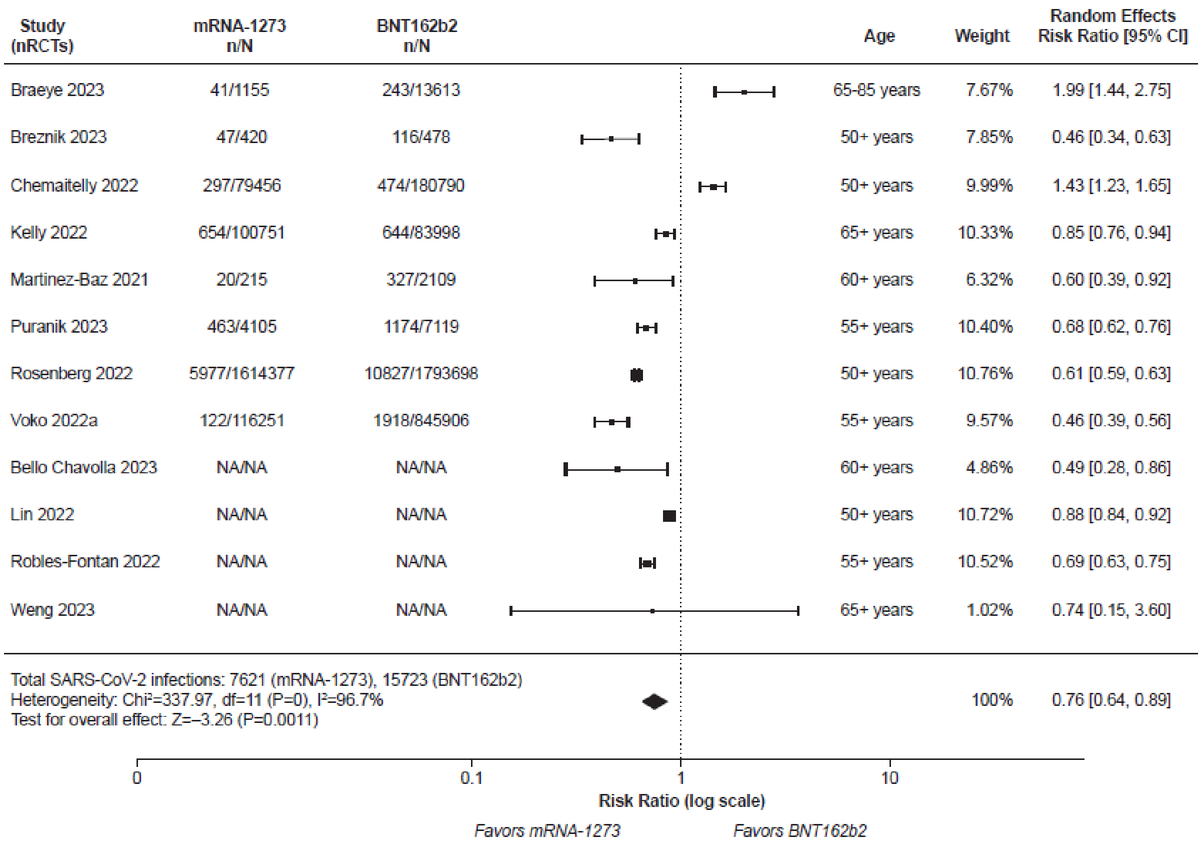


(B) ***Laboratory-Confirmed Symptomatic SARS-CoV-2 Infection***

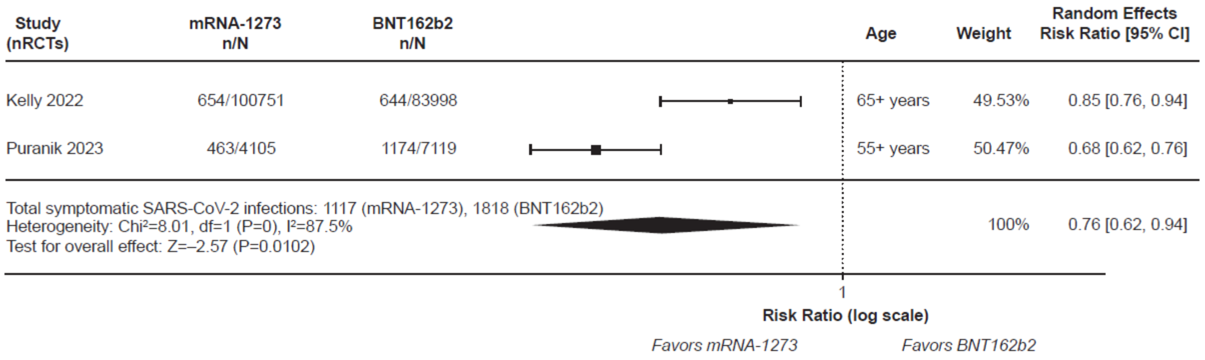

### (C) *Severe SARS-CoV-2 Infection*

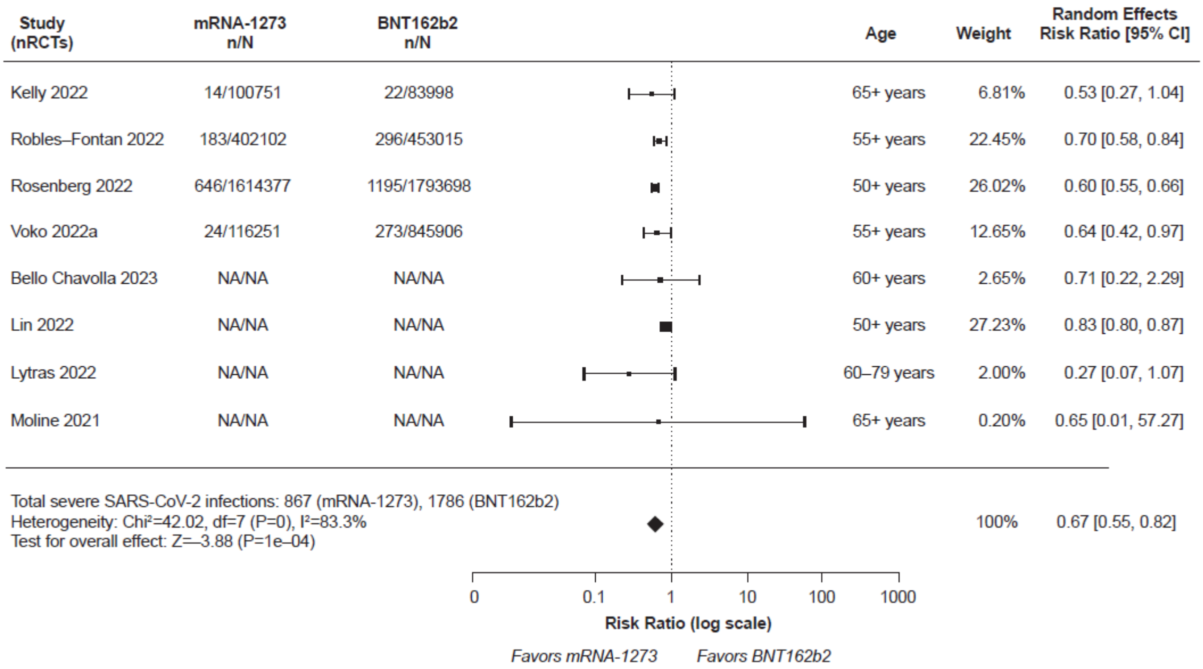

### (D) *Hospitalization*

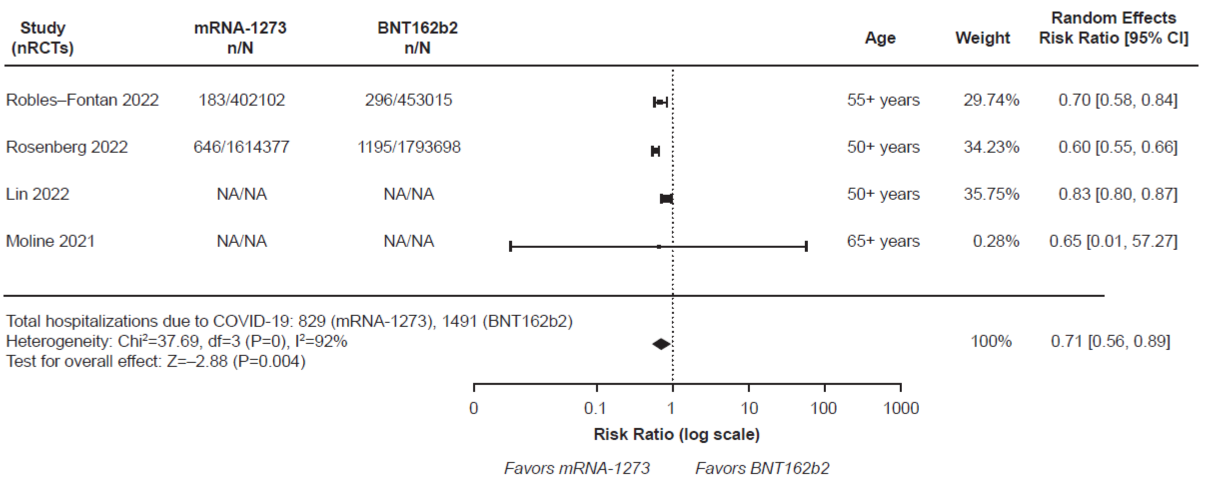


### (E) *Death*

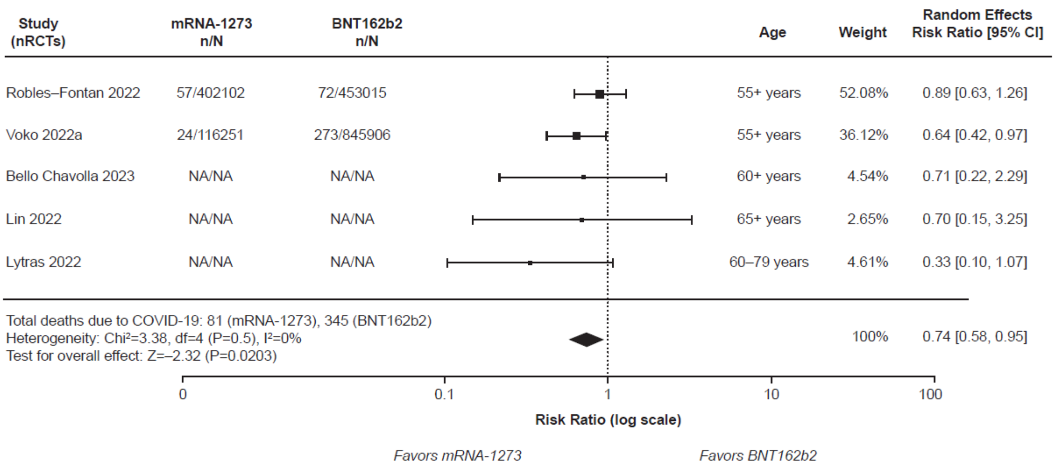

**Figure S7. Meta-analysis results on clinical effectiveness outcomes of the mRNA-1273 versus BNT162b2 COVID-19 vaccines in the subgroup of older adults aged ≥50 years in studies assessing the Delta variant.**

**(A) *SARS-CoV-2 Infection***

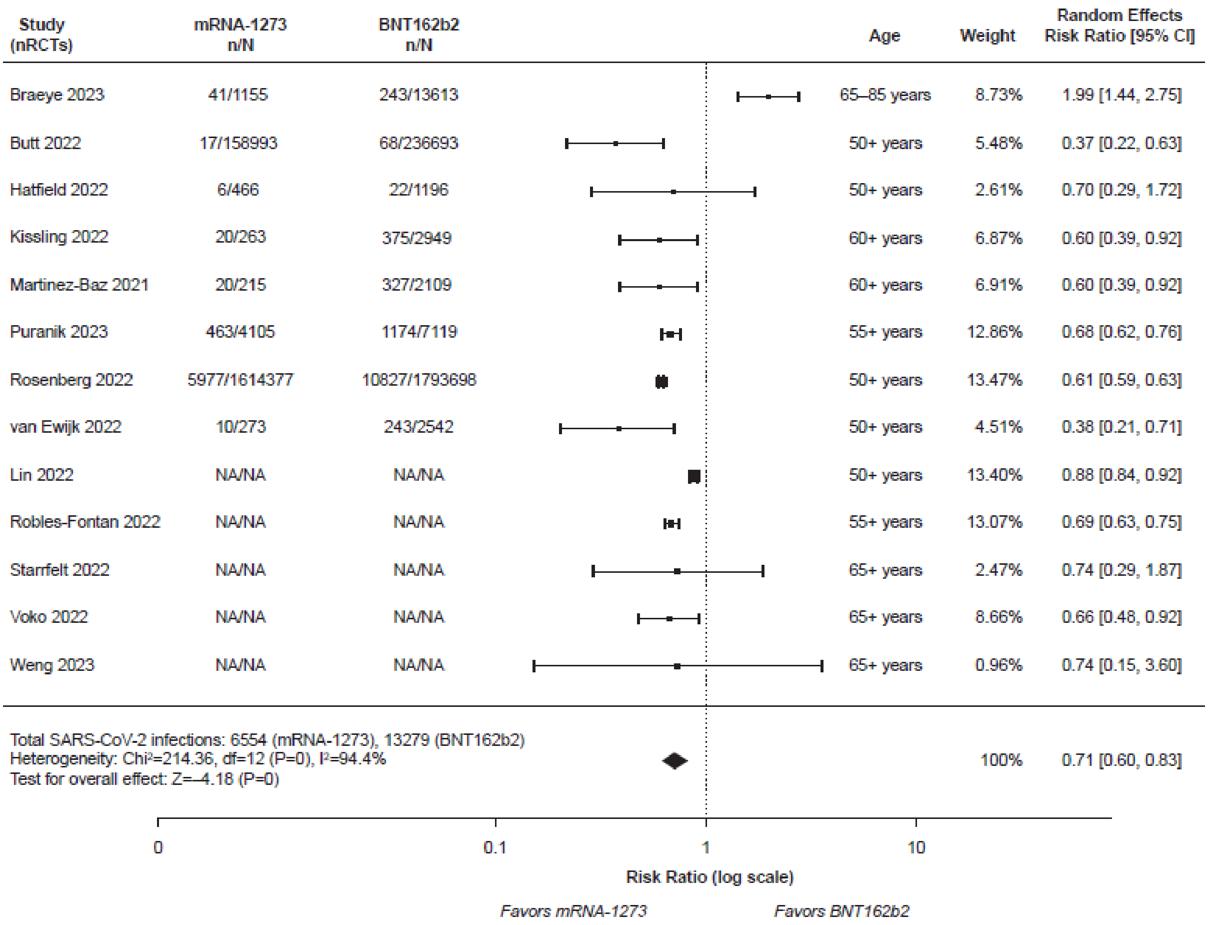

**(B)** ***Laboratory-Confirmed Symptomatic SARS-CoV-2 Infection***

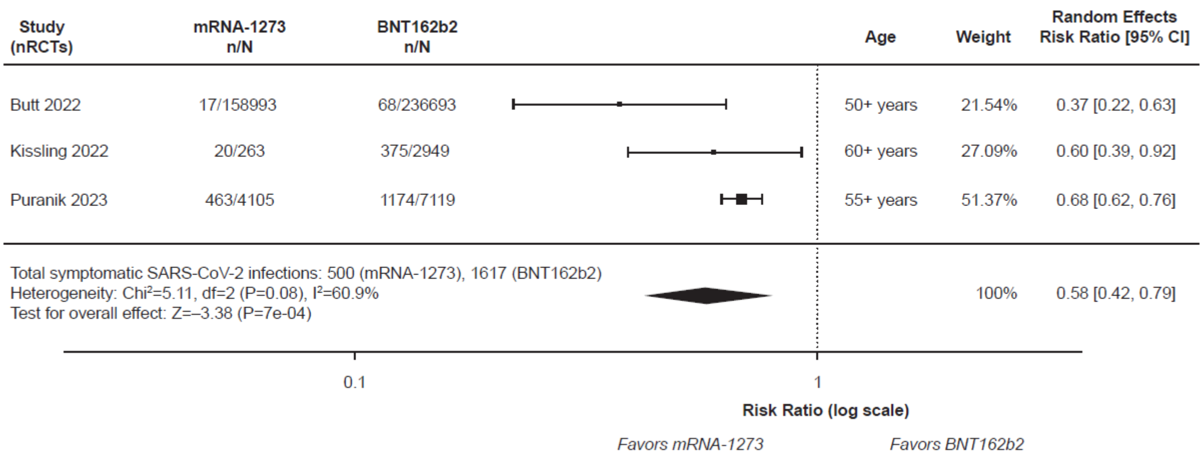


**(C)** ***Severe SARS-CoV-2 Infection***

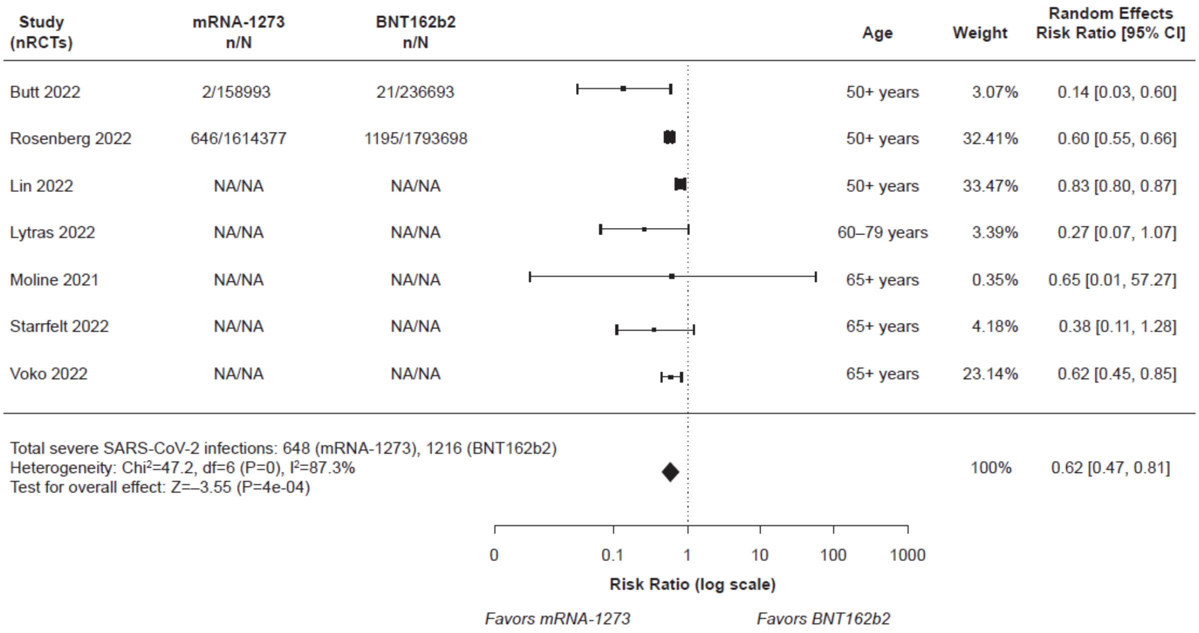

### (D) *Hospitalization*

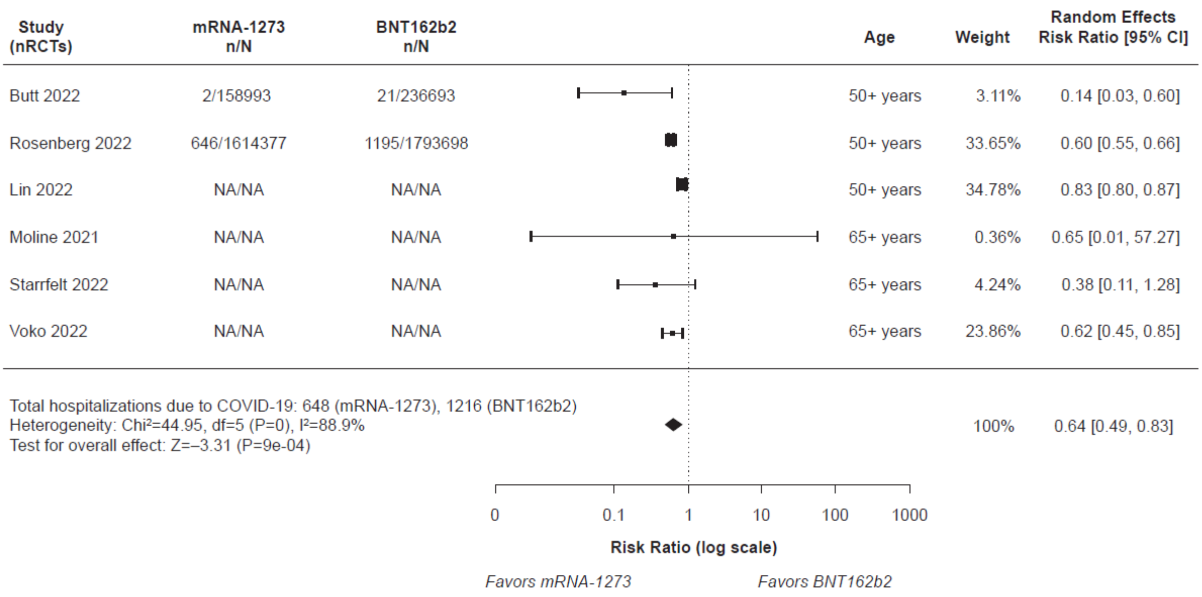


### (E) *Death*

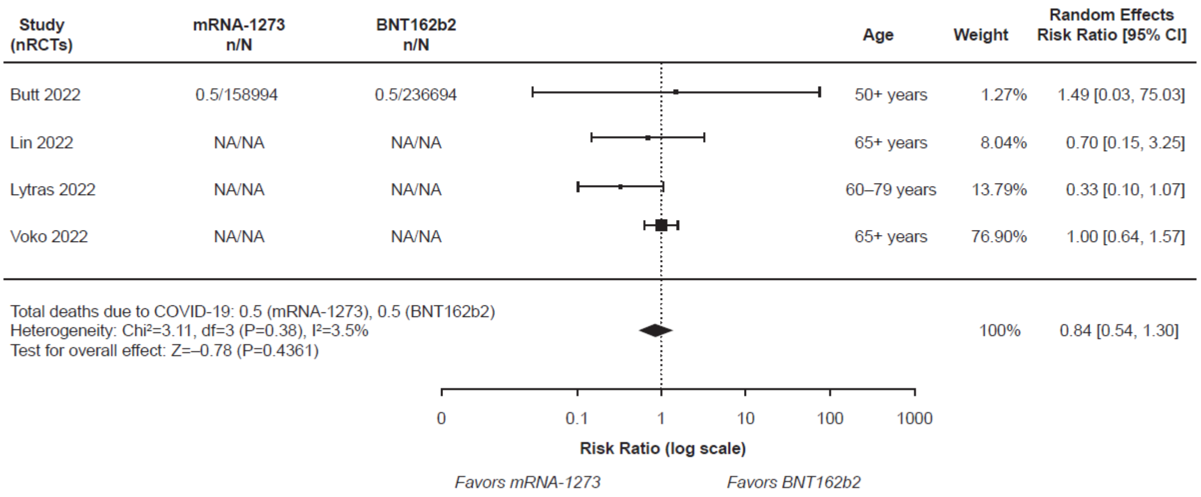

**Figure S8. Meta-analysis results on clinical effectiveness outcomes of the mRNA-1273 versus BNT162b2 COVID-19 vaccines in the subgroups excluding studies that evaluated only VE.**

**(A) *SARS-CoV-2 Infection***

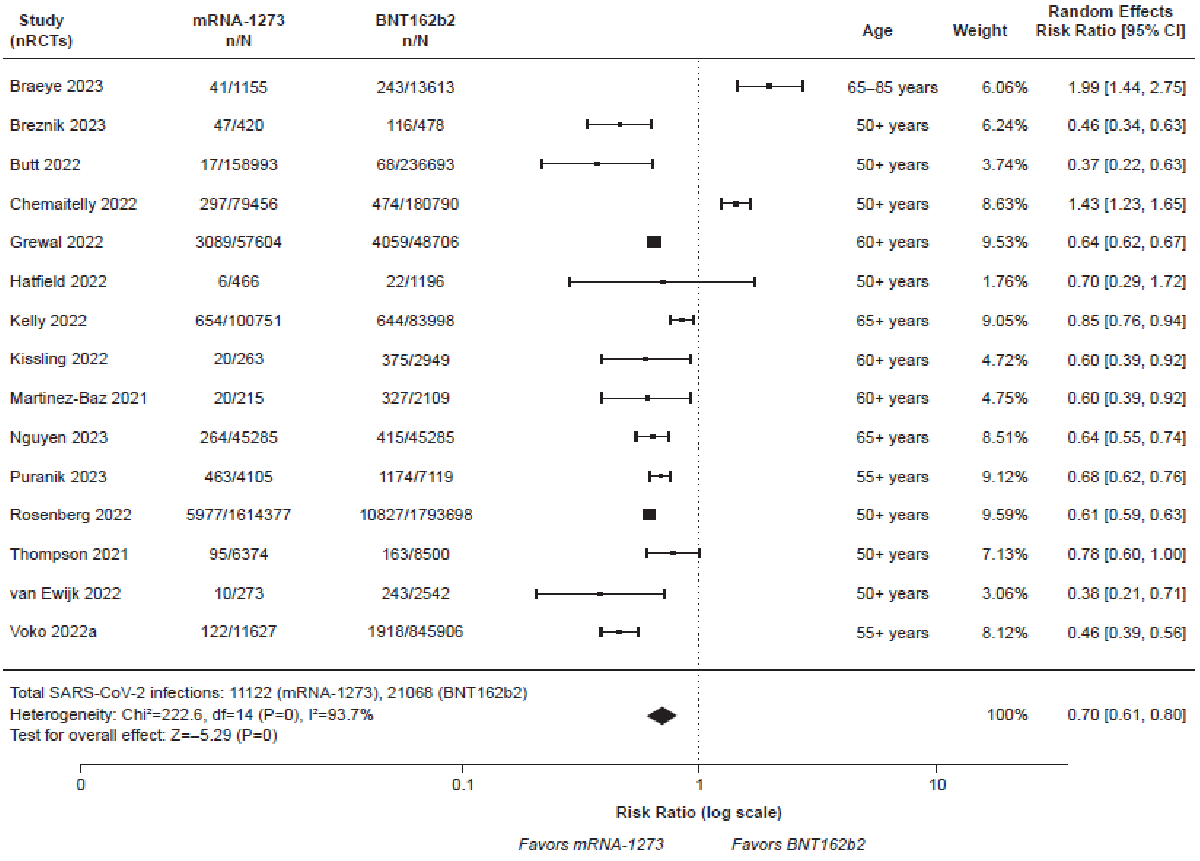

### (B) *Severe SARS-CoV-2 Infection*

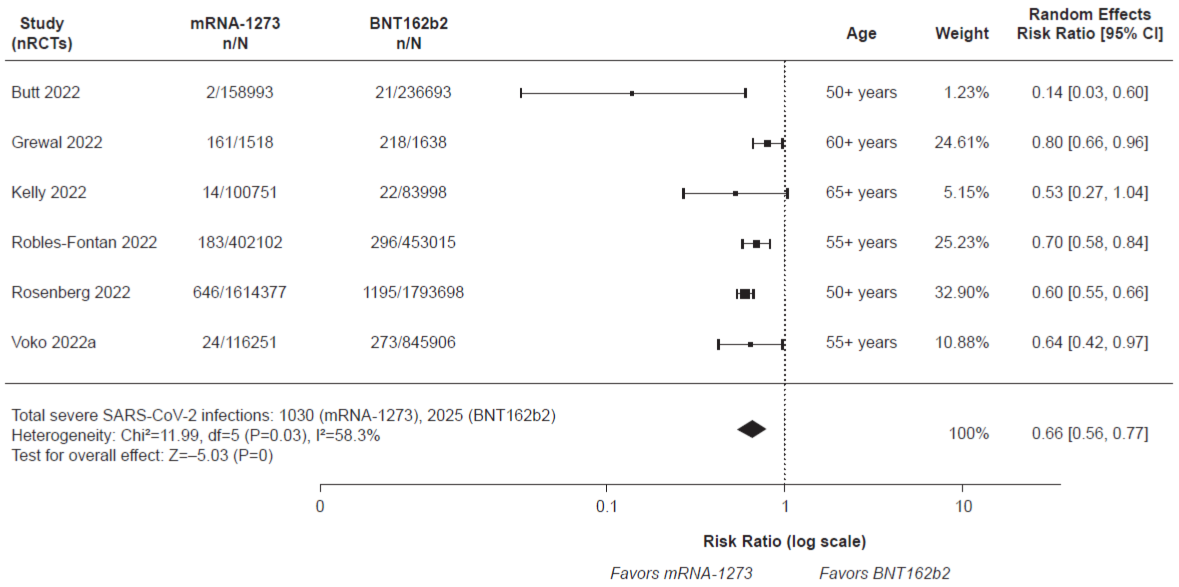


### (C) *Hospitalization*

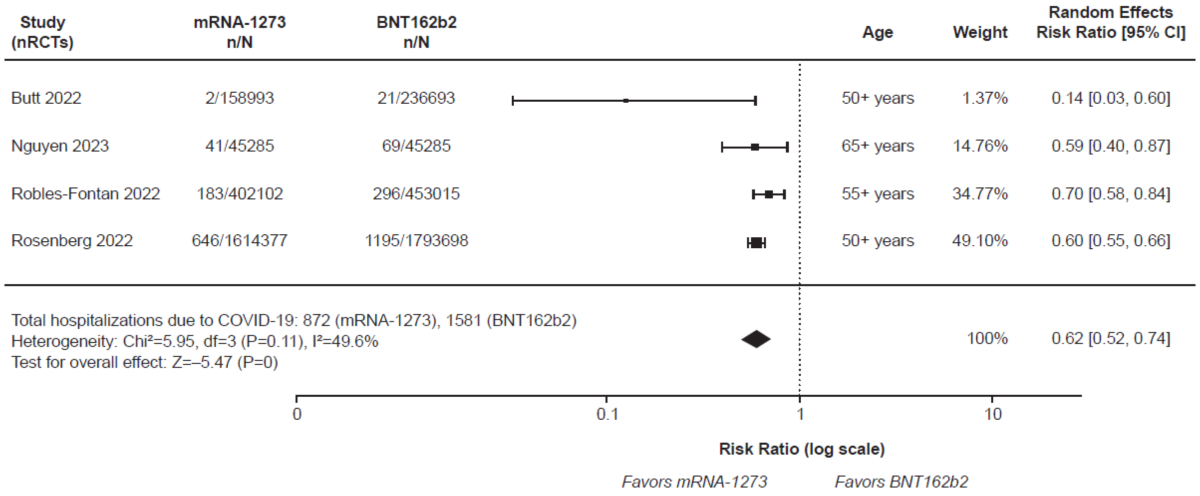


### (D) *Death*

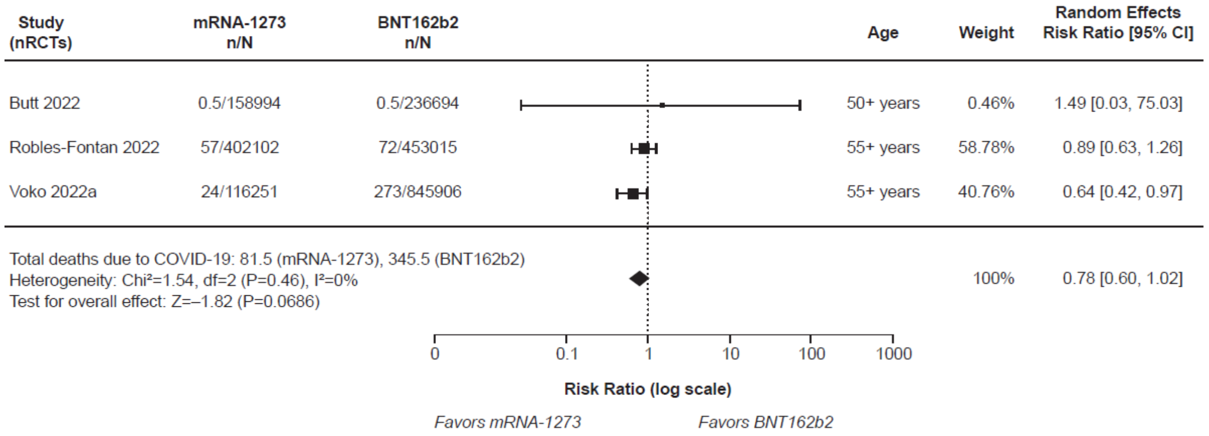

**Figure S9. Funnel plots and Egger's test to assess publication bias.**

**(A)** ***SARS-CoV-2 Infection***

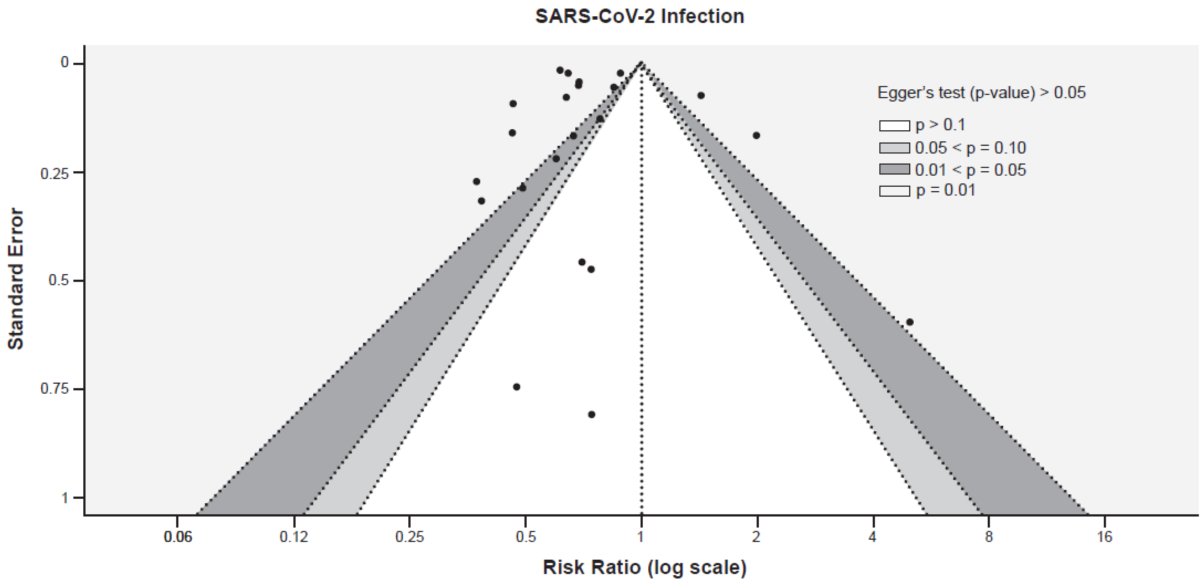


**(B)** ***Laboratory-Confirmed Symptomatic SARS-CoV-2 Infection***

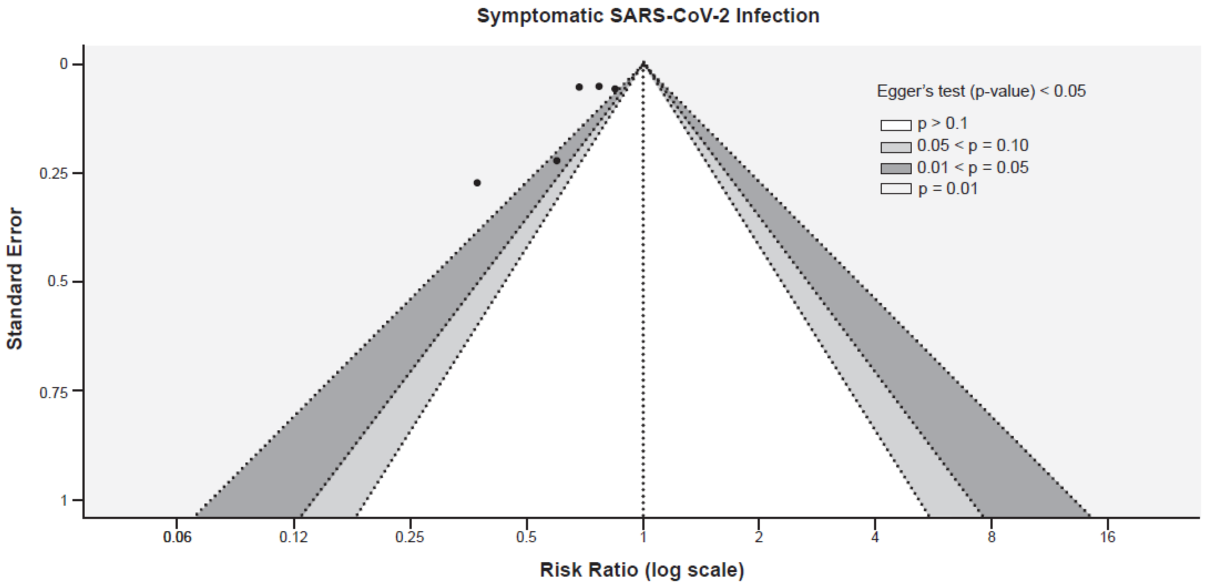

**(C)** ***Severe SARS-CoV-2 Infection***

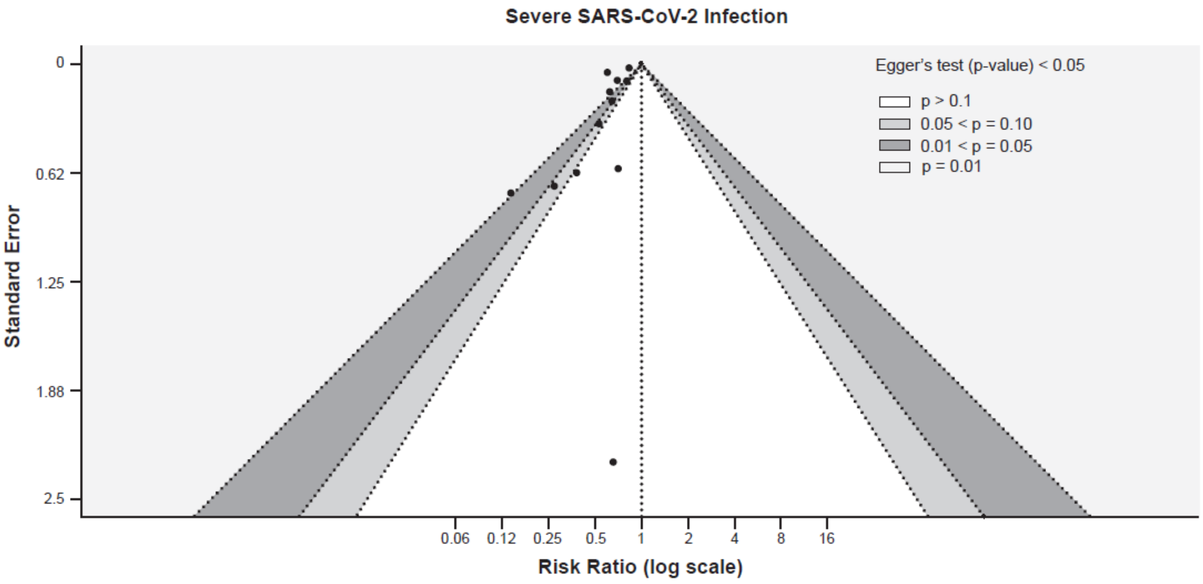


**(D)** ***Hospitalization***

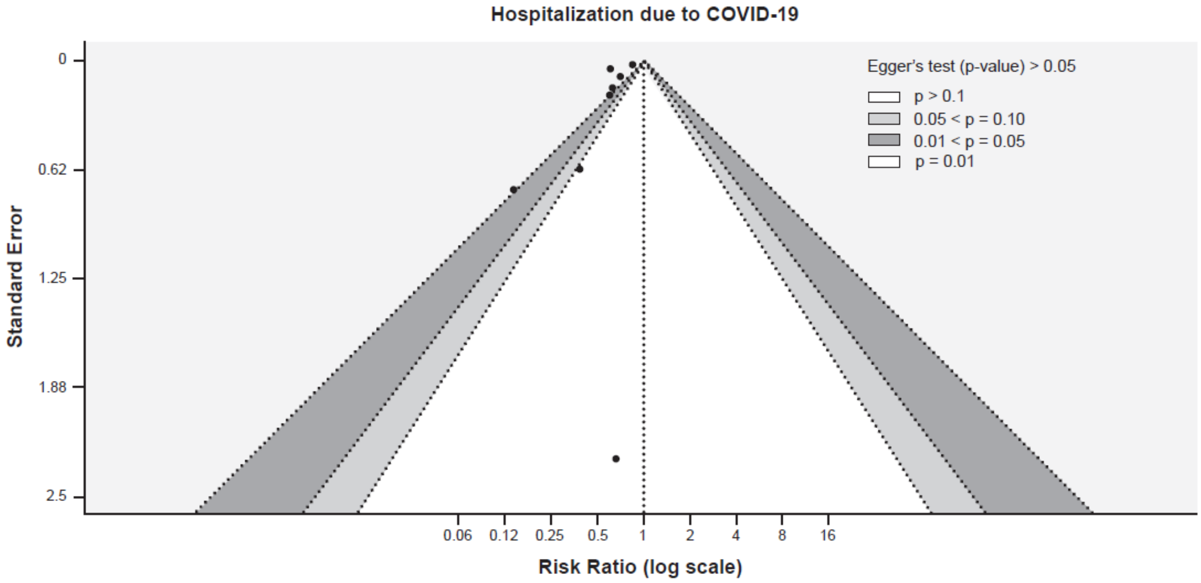

**(E) *Death***

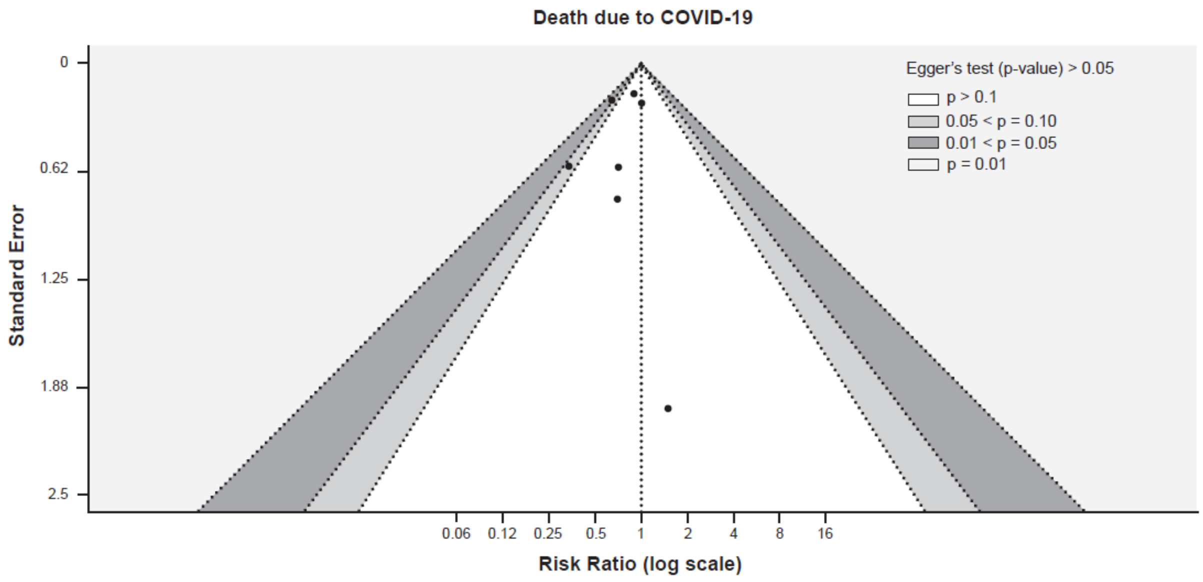
Death due to COVID-19
Egger's test (p-value) > 0.05
p > 0.1
0.05 < p = 0.10
0.01 < p = 0.05
p = 0.01
Standard Error
0
0.62
1.25
1.88
2.5
0.06
0.12
0.25
0.5
1
2
4
8
16
Risk Ratio (log scale)
